# Atrial Fibrillation Drivers: Redefining the Electrophysiological Substrate

**DOI:** 10.1101/2020.12.14.20248171

**Authors:** Markus Rottmann, Anna Pfenniger, Shin Yoo, David Johnson, Gail Elizabeth Geist, Suman Mandava, Amy Burrell, Bradley P. Knight, Rod Passman, Rishi Arora

## Abstract

**Background:** We performed high-density mapping of persistent atrial fibrillation (AF) in animals and patients (1) to test that AF is due to ≥1 reentries, and (2) to characterize activation delay and reentries pre/ post pulmonary vein isolation (PVI). We determined electrophysiological characteristics that may predispose to the induction, maintenance, and reduction of AF.

**Methods and Results:** This study includes 48 dogs and nine patients. 43 AF- and five sinus/ paced rhythm dogs (3-14 weeks rapid atrial pacing) were studied at open chest surgery with 117 epicardial electrograms (EGMs) (2.5mm dist.) in 6 bi-atrial regions. Rotational activity automatically detected with a new algorithm tracking the earliest and latest activation in all regions (5±2 per region) were stable over 424±505ms [120– 4940ms]. Reentry stability was highest in the right atrial appendage (RAA) (405±219ms) and the posterior left atrium (PLA) (267±115ms) and anchored between >=3 zones of activation delay (15±5ms, median 13ms) defined as >10ms per 2.5mm. Cycle length (CL) and degree of focal fibrosis were highest in the PLA and left atrial free wall (LAFW) with 94±7ms, 96±5ms, and 49±14%, 47±19%. Fiber crossing density correlated with the stability of rotational activity (R=0.6, P<0.05). Activation delay was 2x higher in AF compared to sinus rhythm/paced rhythm (interval 200-500ms). Activation delay zones > 10ms were at the same locations, but increased 4x during AF vs. SR and were located at fiber crossings, fibrosis/ fat zones. Stability of rotational activity correlated with Organization Index (OI), Fraction Index (FI), Shannon’s Entropy (ShEn), and CL (R>0.5, p< 0.0001). PVI in five hearts increased CL [2-14%] and reduced stability of rotational activity in nearly all regions remote to the pulmonary veins (PVs). Also in the clinical evaluation in nine patients using the HD-catheter (16 electrodes, 3mm dist.) activation delay at the reentrant trajectory was 2x higher at edges with maximal delay (20.5±8.1ms, median 19.6ms) vs (9.3±8.8ms, median 9.2ms) and 1.4 x higher during AF (13.0±18.7ms, median 7.2ms) compared to SR/ CS-pacing (18.0±11.6ms, median 17.7ms).

**Conclusion:** Rotational activities in all bi-atrial regions anchored between small frequency-dependent activation delay zones in AF. PVI led to beneficial remodeling in bi-atrial regions remote to the PVs. These data may identify a new paradigm for persistent AF.

**Subject Terms:** Arrhythmias, Atrial Fibrillation, Cardiac Electrophysiology, High-Density Mapping, Catheter Ablation, Pulmonary Vein Isolation, Fibrosis

**Clinical Perspective:** *What Is New?:*

- Rotational activity trajectories based on high-resolution mapping follow propagation line patterns.
- Rotational activities anchor frequently between small frequency-dependent slow conduction zones in all bi-atrial regions.
- Slow conduction zones are fiber crossings zones and develop into fibrosis and fat regions over time.
- PVI reduces slow conduction zones and AF drivers in regions remote to the PVs in both atria.

*What Are the Clinical Implications?:*

- The new method for the robust detection of rotational activity based on the earliest and latest activation may be useful for an improved AF treatment.
- Stability of rotational activity may be predicted with the correlated substrate characteristic fiber crossings density, with slow conduction zones, and with established electrogram measures in the different atrial regions.
- PVI leads to beneficial remodeling in all regions remote to the PVs in the left atrium and right atrium.

## INTRODUCTION

The multiple wavelet reentry hypothesis as a mechanism for atrial fibrillation (AF) was proposed by Moe and Abildskov in 1959.^1^ In 1985, Allessie experimentally confirmed this hypothesis showing that multiple wavefronts throughout the atrium maintain themselves by reentry around continuously shifting areas of functional conduction block.^2^ In 1998, Haïssaguerre demonstrated in his seminal paper that pulmonary veins (PV) triggers can be critical for the initiation of AF and PVI can reduce the occurrence of AF.^3^ However, despite extensive research, the mechanisms of AF are still not clear. ^4,5^ The present work aimed to determine electrophysiological characteristics both in a human-like large animal AF model and in AF patients that may facilitate the occurrence of local arrhythmias. Many mapping studies aimed to test the hypothesis that AF is caused by either focal or reentrant drivers in AF using atrial electrograms (EGMs) based on sequential or simultaneous multisite site mapping at the endocardium and epicardium using different types and numbers of electrodes. ^6-19^ Several studies showed that reentry circuits can drive AF, that slow conduction leads to reentry in AF and that PVI reduces AF.^20^ Although the overall association between areas of slow conduction and AF drivers is evident, several issues remain unsettled (Figure 1A) including:

**Figure 1:**
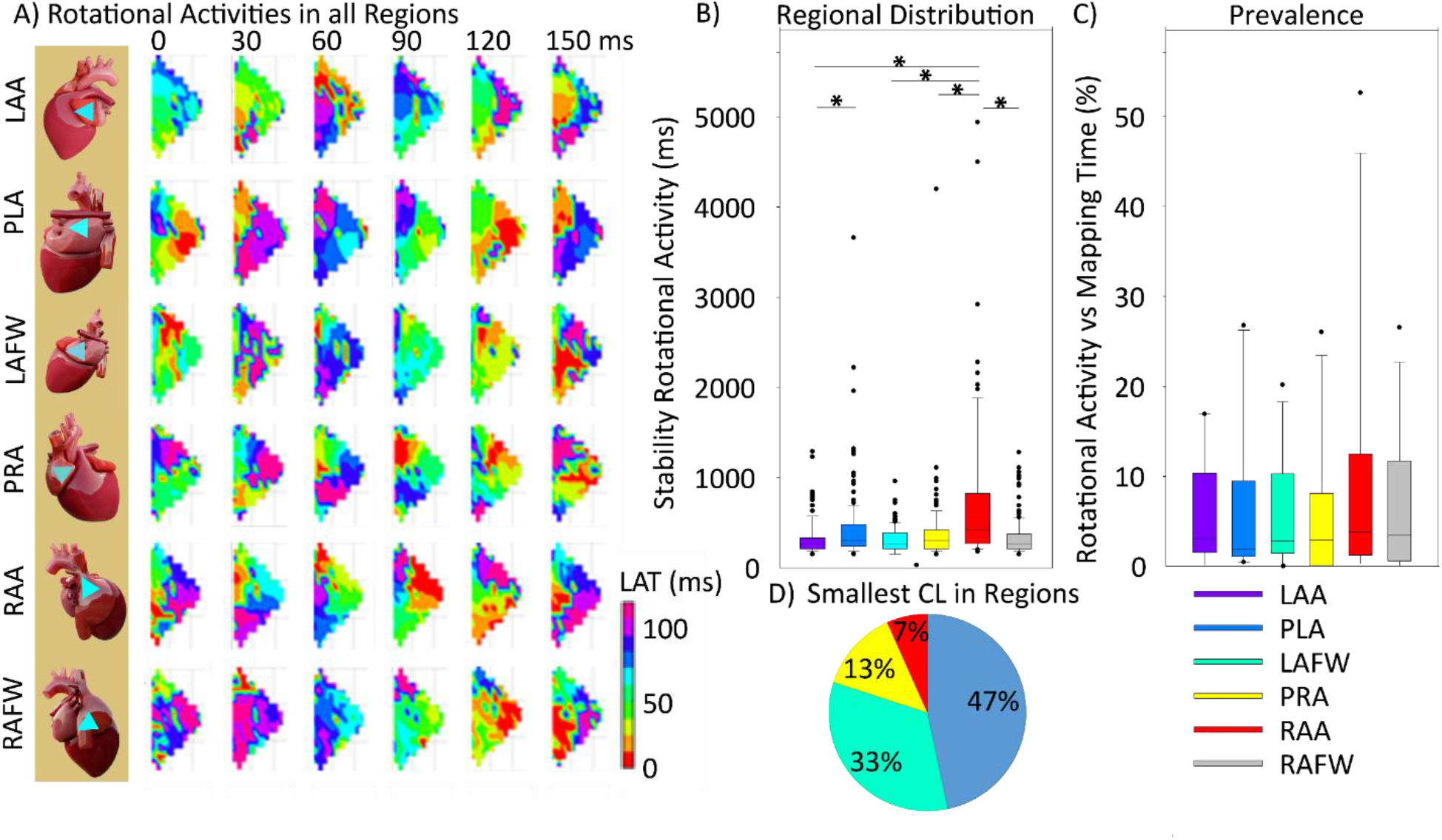
Multiple rotational activities in all regions in both atria. A, (Left panel) Location of mapping plaque (blue triangle) in the 6 bi-atrial regions. (Right panel) Multiple rotational activities in high-density local activation time (LAT) maps in all 6 atrial regions in heart #10. B, The highest stability of rotational activities was detected in the RAA and second in the PLA. C, Percentage of rotational activity time episodes vs total mapping time in all regions. The average rotational activity episodes covered 12 % of the total mapping time in the RAA. D, Regional distribution of smallest CL of all regions in percentage of all hearts. * P < 0.05 for all comparisons.

1. What are the regional differences in rotational activities and activation delay zones?
2. Is there a critical activation delay and size threshold during AF that can differentiate tissue regions producing stable reentry from those that do not?
3. What is the shape of the reentrant trajectory based on high-resolution mapping data?
4. Is the stability of rotational activity correlated with slow conduction zones, established electrogram measures, and substrate characteristics?
5. What are the effects of PVI rotational activity and slow conduction zones on regions in both atria remote to the pulmonary veins?
6. Are slow conduction zones similar in animals and patients during AF and SR/ paced rhythm?

This study aimed to answer these questions in a canine model and patients of persistent AF.

## METHODS

An expanded materials and methods can be found in the supplementary material online Supplementary Data.

### Animals and Experimental Rapid Atrial Pacing Protocol

We induced AF in 43 hound dogs by rapid atrial pacing (RAP) (600 beats/min) for 3-8 weeks. In five dogs we also performed PVI. Before all the procedures, animals were premedicated with acepromazine (0.01–0.02 mg/kg, Vedco) and induced with propofol (3–7 mg/kg, Zoetis). All experiments were performed under general anesthesia (inhaled) with isoflurane (1-3 %). Adequacy of anesthesia was assessed by toe pinch and palpebral reflex. The Animal Care and Use Committee of Northwestern University approved this experimental protocol. The protocol is in line with the Guide for the Care and Use of Laboratory Animal (US National Institutes of Health Publication). Detailed methods are given in the Data Supplement. For pacemaker insertion, the right jugular vein was accessed by direct cutdown and ligated distally. A bipolar screw-in Medtronic pacing lead was inserted through an incision in the right jugular vein. The tip of the lead was fluoroscopically placed and fixed in the RA appendage after confirming an adequate capture threshold (<0.5 mV with a pulse width of 0.4 ms). The proximal end of the pacing lead was connected to a custom-modified Medtronic programmable pulse generator that was subsequently implanted in a subcutaneous pocket in the neck. After all the incisions were closed, the dogs were allowed to recover from anesthesia and were returned to the animal quarters. After confirming an adequate threshold for atrial capture (<0.5 mV with pulse width 0.4 ms), rapid atrial pacing was performed incrementally over 1-3 days until adequate capture was confirmed at 600 bpm. The pacing was continued for 3-14 weeks until persistent AF was induced.

### High-Density Mapping During Open Chest Surgery

Atrial epicardial mapping studies were performed during open-heart surgery in AF on the beating heart. High-resolution mapping was performed with the UnEmap mapping system (University of Auckland, Auckland, New Zealand) which consists of a triangular mapping plaque and records 117 bipolar electrogram signals (1kHz sampling rate, 117 electrodes, interelectrode distance of 2.5 mm) covering a total area of 7.3 cm^2^ on the atrial epicardial surface. Bipolar electrograms were recorded from six different regions in both atria: posterior left atrium (PLA), left atrial free wall (LAFW), left atrial appendage (LAA), posterior right atrium (PRA), right atrial free wall (RAFW), and right atrial appendage (RAA) (Figure 1B, Supplemental Figure S1-S13). Data were digitally recorded and processed with the UnEmap mapping system software. Data were collected with the GE Prucka Cardiolab system (GE Healthcare), and two rectangular mapping plaques with 12 electrodes (3 x 4 electrodes, interelectrode distance 5mm). The data were transferred and stored on a personal computer for further analysis. The AF EGM measures including Dominant Frequency (DF), Organization Index (OI), Fractionation Interval (FI), Shannon’s Entropy (ShEn) were calculated with in-house MATLAB (MatLab, Mathworks, Natick MA) programs. We excluded electrodes (< 10%) with inadequate quality due to noise or poor contact. The sequential activation maps of persistent AF were constructed for 10 consecutive seconds in each atrial region in all hearts. The data were analyzed automatically for activation times using in-house MATLAB programs by detecting the steepest negative slope in the bipolar electrograms.^21^

### Electrogram Measures

#### Cycle Length (CL) -

CL was calculated with the steepest negative slope using the maximal negative slope of the bipolar electrogram.

#### Dominant Frequency (DF)

DF has been shown to correspond to rotational activity in AF.^22,23^ Dominant frequency was calculated with the highest power in the power spectrum using the fast Fourier transform and bandpass filtering with cutoff frequencies of 40 and 250 Hz.

#### Organization Index (OI) -

OI is a frequency domain parameter of the temporal organization or regularity.^24, 25^ OI was computed as the area under 1-Hz windows of the DF peak, and the next 3 harmonic peaks divided by the total area of the spectrum from 3 Hz up to the fifth harmonic peak. ^24, 25^

#### Fractionation Interval (FI) -

FI is the mean interval between deflections detected in the bipolar signal segment.^26^ Deflections were defined with the following conditions: (a) the bipolar peak-to-peak amplitude was greater than a defined noise level, (b) the time between a neighbored positive peak and the negative peak was within a 10 ms window, and (c) the detected deflection did not overlap within 50 ms with another detected deflection. The noise level was determined by the amplitude level, which avoids the detection of noise-related deflections in the iso-electric portions of the signal.

#### Shannon Entropy (ShEn) -

ShEn is a measure of the complexity of the EGMS.^27^ Amplitude values of each EGM segment (3908 or 4000 amplitudes, 1 kHz, or 977 Hz sample rate) were calculated, and these were binned into 1 of 29 bins with a width of 0.125 standard deviation (SDs). ShEn was calculated using the following formula, with *p*_*i*_ the probability of an amplitude value occurring in bin i.

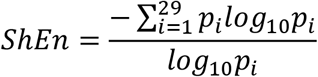

#### Detection of Slow Conduction Regions -

Conduction velocity cannot be measured from epicardial mapping alone as it may not include the entire pathway of the wavefront between 2 points; we, therefore, used the term activation delay to describe this measure. To quantify changes in epicardial activation delay objectively and reproducibly, all electrogram data including its geometric coordinates were imported into MatLab, and a proprietary code was written for automated calculation of epicardial activation delay. To correlate epicardial activation during AF, the epicardial maps were partitioned into small and fixed regions of interest (ROIs) of 2.5 mm × 2.5 mm, and the activation delay in each ROI during AF was automatically measured.

#### A New Method for the Robust Detection of Rotational Activity -

In the literature, the trajectory of reentries is calculated based on phase singularities.^20, 28, 29^ However, this method has several limitations for robust detection and can falsely detect phase singularities in the absence of rotors.^30^ In the present work, a completely new method was developed to detect the trajectory of rotational activity more robustly. Reentry was detected based on activation time maps showing 360-degree rotations using both, earliest and latest activation x,y-locations in the activation time maps over time. Multiple activation time maps were calculated in 5ms steps over 10 seconds. In each of these local activation time (LAT) maps the position of the earliest activation and the position of the latest activation was marked. If these detected locations of earliest and latest activation correlated over time with repetitive patterns showing loops, these activation patterns were considered as rotational activities. The computer program was written in a proprietary code in MatLab. The reentry stability was assessed as the duration of observed reentrant activity over time. Additional manual annotation of 360-degree rotations in local activation time maps was used for confirmation of the automatic detections of rotational activities. And the results of this new method were compared to a previously described method based on phase singularity detections based on the Hilbert phase and sinusoidal recomposition. ^31^ In addition to reentries, also focal activities were automatically detected. Focal activities and their dimensions were detected as simultaneous earliest activities within activation time maps in five msec time steps.

### Pulmonary Vein Isolation and Remapping-

In the open chest model, pulmonary vein isolation was performed on the epicardium with an isolator synergy ablation clamp (AtriCure Synergy, AtriCure, Inc., Cincinnati, OH) for bipolar radiofrequency energy lesions. The Atricure system includes a bipolar RF clamp and an RF generator. Lines of conduction block were generated by RF energy delivery (75 volts, 750 milliamps) to the atrial tissue compressed between the jaws of the clamp. Energy delivery was continued until the lesion was transmural, indicated by steady and reduced conductance between electrodes in the clamp. Post PVI, epicardial mapping was performed again in all biatrial regions with the high-resolution mapping plaque.

### Tissue Analysis

We further performed a detailed tissue section analysis in six animals. After the open chest epicardial mapping procedure, we excised the heart out of the chest and immersed in an ice-cold cardioplegia solution containing (mmol/l) NaCl 128, KCl 15, HEPES 10, MgSO_4_ 1.2, NaH_2_PO_4_ 0.6, CaCl_2_ 1.0, glucose 10, and heparin (0.0001 U/ml); pH 7.4. The solutions were equilibrated with 100% O_2_. We then cannulated the heart via the aorta and perfused with ice-cold cardioplegia solution containing protease inhibitors (Millipore Sigma, P8340) until the vessels were clear of blood, and tissue was cold. The 6 mapped bi-atrial tissue regions in the size of the triangular mapping catheter were dissected. Specimens from six dogs were fixed in 10% formalin and embedded in paraffin for further examination.

### Masson’s trichrome staining

Paraffin sections with 5 μm thickness were stained using Masson’s Trichrome stain kit (Sigma). Paraffin was then removed in Xylene for three minutes twice and then in the mixture of 100% Xylene and absolute Ethanol for three minutes twice. Sections were then rehydrated with ethanol series which include absolute Ethanol (twice), 95% Ethanol, 70% Ethanol, and 50% Ethanol. The paraffin sections were treated with Bouin’s mordant at room temperature overnight. The following day the sections were rinsed in running water to remove excess yellow. The sections were stained in Weigert’s Iron Hematoxylin solution for five minutes. Next, it was washed under running water for five minutes and briefly rinsed in distilled water. The sections were then stained in Beibrich Scarlet-Acid fuchsin for five minutes, followed by a rinse in distilled water. Subsequently, the sections were incubated in the phosphomolybdic-phosphotungstic acid solution for five minutes. The issue section was then stained in the Aniline Blue solution for five minutes. The sections were incubated in 1 % Glacial acetic acid for two minutes. The sections were then dehydrated through ethanol series, which include 70%, 90%, and absolute Ethanol (twice). Then, the sections were placed in Xylene for five minutes twice. A coverslip was finally placed using cytosol mounting media on the sections for microscope examination.

### Examination of Uniformity of Fiber Orientation and Degree of Fibrosis -

Masson’s trichrome stained tissue sections at different depths (0, 200, and 500 μm) from the epicardial surface were digitized with the NanoZoomer 2.0-HT (Hamamatsu Photonics. Hamamatsu Japan) at 5x magnification. The whole sections were divided into quadrants by drawing regions of interest for the quantitative morphometric analysis. We quantified uniformity of fiber orientation in RAP atrial tissue sections at 200 μm and 500 μm from the epicardial surface and quantified the orientation of the myocytes. The standard deviation of the detected fiber angle and number of fiber crossing points were quantified in ROI’s of 2.5mm x 2.5mm similar to the electrode sizes of the Unemap mapping plaque using the ImageJ plugin OrientationJ and fiber crossing were defined as angle > 30 degrees between neighbored ROI’s (compare Supplemental Data S45-50). ^32^ We further analyzed the degree of focal fibrosis in Masson’s trichrome stained tissue section at 200 μm levels with a semi-automatic color-based segmentation and the degree of interstitial fibrosis as there was qualitative similarity among sections at different depths using ImageJ and macro.^32^

### Clinical Mapping and Ablation Procedure -

Nine early persistent AF Patients at Northwestern Memorial Hospital were included. The patients received conscious sedation or general anesthesia at the discretion of the treating physician. This study was approved by the institutional review board at Northwestern Memorial Hospital (Northwestern University). After obtaining access, transseptal puncture across the interatrial septum was performed using an SL1 (Abbott, Chicago, IL) or Preface sheath (Biosense Webster, New Brunswick, NJ) and Bayliss RF needle (NRG® Transseptal Needle, Baylis Medical). Intravenous heparin was given with an activated clotting time (ACT) goal of > 300 s. 3D mapping was used at the discretion of the operator. Using the Advisor® HD Grid Mapping Catheter (Abbott) with 16 electrodes arranged in a 4 x 4 grid, EGMs were recorded and tagged according to the anatomic location (posterior wall, roof, lateral wall, septum) on the Ensite NavX mapping system ® (Abbott). Electrograms were sampled at 2 kHz and bipolar electrogram filtering 30 to 300 Hz with a 60Hz noise filter. We detected slow conduction at reentry sites and focal sources near the pulmonary veins in nine patients using the high-definition (HD) mapping catheter (Figure 8A). Example bipolar voltage map and LAT map of the left atrium near the pulmonary veins are given in Figure 8 G-H.

#### Statistics

Continuous variables with normal distribution are reported as mean and SD. For those with non-normal distribution, range and median values are reported. For comparisons of continuous variables, a 2-sample t-test with unequal variance was used if data were normally distributed and a Wilcoxon rank-sum test was used for non-normally distributed variables. For comparisons of multiple variables, 1- or 2-way ANOVA methods were used. Correlation between continuous variables (stability of rotational activities and electrogram measures) was evaluated using linear regression. The normality assumption was tested using the skewness and kurtosis test. A P value <0.05 was considered statistically significant. Analyses were conducted using STATA SigmaPlot 14.0 software (Systat Software, Inc., CA, USA) and MatLab.

## Results

### Baseline Characteristics in the Animals and Patients

48 dogs and nine patients were studied. A total of 43 AF and five sinus/ paced rhythm hound dogs underwent electrophysiological study. During AF, multiple rotational activities of different cycle lengths (103±13 ms, median 103ms) were present in both atria in all 18 hearts. The rotational activity was detected in all regions (32 per heart, 5±2 per region, median 4) with stability over 424±505 ms. (median 270 ms, range 120 – 4940 ms). The animal characteristics are summarized in Table 1.

**Table 1.** Animal Characteristics.

| Animal Characteristics |  |  |  |
| --- | --- | --- | --- |
| Animal | Gender | Weight | Pacing Days |
| 1 | F | 25 | 40 |
| 2 | F | 28 | 42 |
| 3 | F | 24 | 56 |
| 4 | F | 27 | 73 |
| 5 | F | 25 | 35 |
| 7 | F | 22 | 29 |
| 8 | F | 25 | 41 |
| 9 | F | 30 | 41 |
| 10 | F | 22 | 38 |
| 11 | F | 34 | 20 |
| 12 | F | 26 | 28 |
| 13 | F | 30 | 26 |
| 14 | F | 25 | 15 |
| 15 | F | 22 | 54 |
| 16 | F | 35 | 119 |
| 17 | F | 32 | 29 |
| 18 | F | 22 | 26 |
| 19 | F | 23 | 27 |
| 21 | M | 29 | 55 |
| 22 | M | 33 | 53 |
| 23 | F | 31 | 54 |
| 24 | F | 32 | 28 |
| 25 | F | 25 | 98 |
| 26 | F | 22 | 34 |
| 27 | F | 18 | 55 |
| 28 | F | 27 | 40 |
| 29 | F | 30 | 49 |
| 30 | F | 20 | 40 |
| 31 | F | 29 | 41 |
| 32 | F | 23 | 35 |
| 33 | F | 27 | 41 |
| 34 | F | 22 | 61 |
| 35 | M | 32 | 85 |
| 36 | F | 35 | 65 |
| 37 | F | 30 | 55 |
| 38 | F | 34 | 98 |
| 39 | F | 27 | 49 |
| 40 | F | 24 | 42 |
| 41 | F | 39 | 61 |
| 42 | F | 32 | 49 |
| 43 | M | 31 | 47 |
| 44 | M | 26 | 72 |
| 45 | M | 31 | 54 |
| 46 | M | 34 | 56 |
| 47 | M | 30 | 52 |
| 48 | M | 32 | 85 |

Gender: F, female, M, male.

### Regional Differences in Rotational Activity

Rotational activities were detected in all 6 bi-atrial regions. Figure 1B) is a representative example of AF showing multiple rotational activities of different CLs in all six atrial regions in the same heart (heart #10). Rotational activities, likely AF drivers, were significantly faster in the PLA (94±7 ms, median 93 ms) and LAFW (96±5 ms, median 95 ms) (compare Figure 7A). Shortest CL was detected in the PLA in 47%, in the LAFW in 33%, in the PRA in 13%, and in the RAA in 7% of all animals (Figure 1D). In contrast, the longest CL was in the RAA in 90% and the RAFW in 10% of all animals. Rotational activities were most stable in the RAA (405±219 ms, median 420 ms) and second in the posterior left atrium (PLA) (267±115 ms, median 300 ms). The highest stability of rotational activity was detected in the RAA in 80%, in the RAFW and LAFW in 10% of the animals. The second most stable reentries were detected in the LAA in 60% and the RAFW in 40% of all animals. Importantly, rotational activity was most present in the RAA in 12% (average) of the mapping time, second in the RAFW and third in the PLA of the mapping time (Figure 1C).

### Redefining Trajectory and Anchoring of AF Drivers

Figure 2A is a representative example of AF activation showing stable rotational activities in all 6 bi-atrial regions in the same animal (heart #10). The rotational activities during AF anchored frequently at slow conduction zones (number n>=3) of activation delay (14.9±5.2 ms, median 13.0 ms, range 10 – 35 ms) over a distance of 2.5 mm (Figure 2B, and Supplemental Data Figure S24-S25). In contrast-to the phase singularity analysis of reentry in previous reports, we defined the trajectory and anchoring of rotational activities based on activation and activation delay in high-resolution mapping data. Figure 2D (left panel) shows an oval shape schematic of a reentrant circuit trajectory, which is typically described in the literature. In contrast, high-density mapping shows that the trajectory of reentry based on earliest and latest activation was different with patterns of lines with normal conduction (Figure 2E) and slow conduction at the edges of these line patterns. Figure 2D (right panel) exemplarily shows the schematic of the new reentry model in form of a polygon with observed curvature angle (138°) between these propagation lines. Activation delay at the edges of the reentrant trajectory (15±5 ms, median 13 ms) was four times slower compared to the straight and fast conduction at propagation lines with activation delay (4.1±2.0 ms, median 4.0 ms) (Figure 2C). These detected slow and highly curved conduction regions measured > 2.5 mm x 2.5 mm and were stable over time. Both trajectories of earliest and latest activation within activation time maps over time had the same overlapping slow conduction regions (Figure 2 B, left and right panel). As described in the literature, slow conduction zones are located at abrupt changes in the fiber direction, compare schematic in Figure 2 F.) Using this newly developed method based on the earliest and latest activation, the accuracy of automatically detecting reentries increased by about 30% compared to previously described methods, which used Hilbert phase maps.

**Figure 2:**
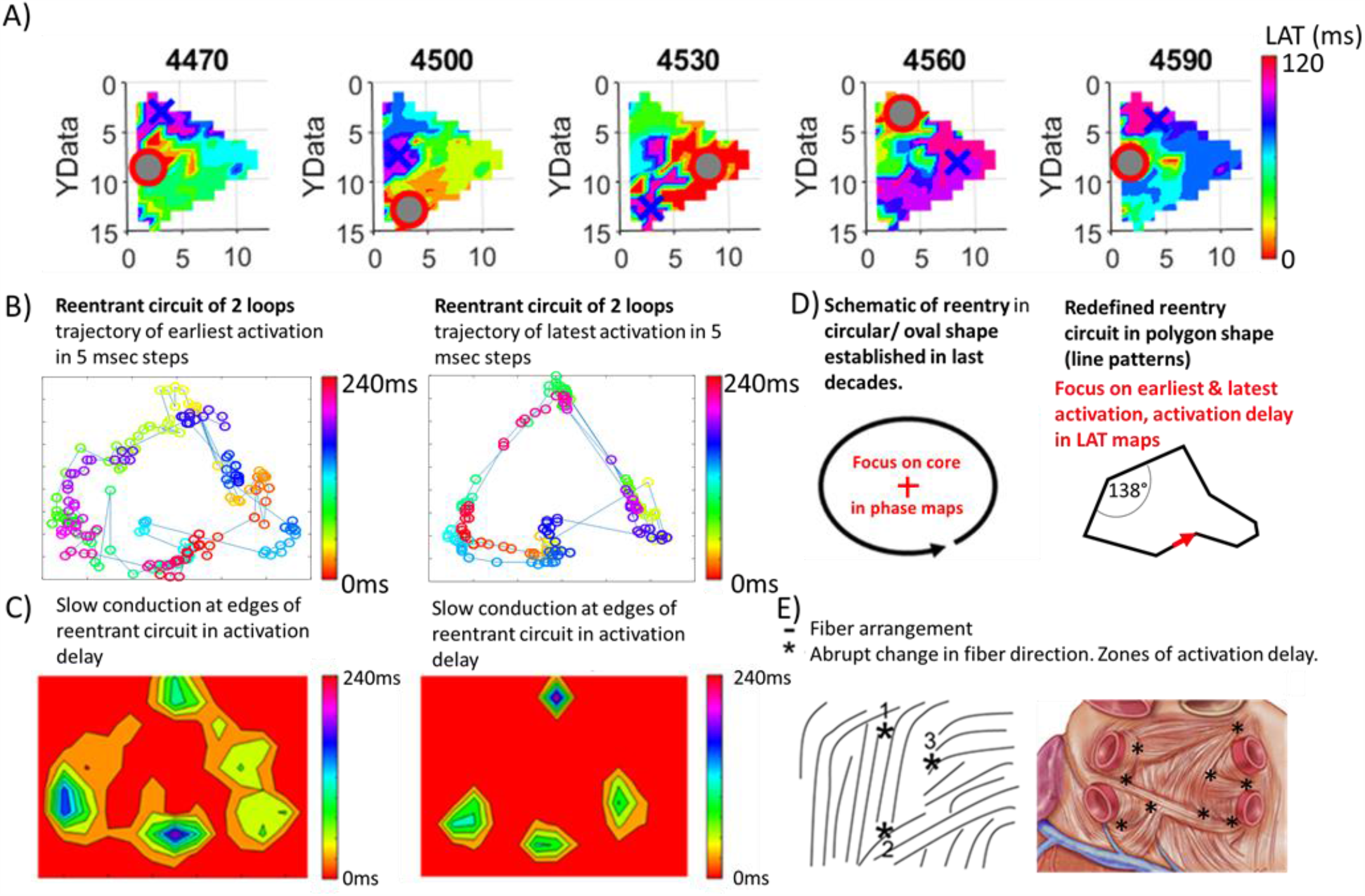
Multiple interacting rotational activities in all regions in both atria. A, Local activation time maps showing the stable reentrant circuit in the RAA. The earliest activation is marked in red (symbol: o). The latest activation is marked in blue (symbol: x). Data from a representative example of AF attributable to activation from the stable reentrant site in the RAA (from heart #1). B, Reentrant trajectory of 2 loops of earliest and latest activation and C, slow conduction zones at the edges of the trajectory of earliest and latest activation. Reentrant circuit trajectory along with patterns of lines (up to 6 mm in the panel with curvature angle between the lines and >4 times slower conduction at edges in propagation (highest curvature angles). Conduction patterns potentially because of patterns in fiber directions. D, Reentrant circuit trajectory of a circular reentry (established schematic model in the last decades). E, Redefined reentrant circuit trajectory along with patterns of lines and with curvature angle between the lines at >4 times slower conduction at edges in propagation. F, Conduction pattern lines potentially because of patterns in fiber directions. (Left panel) Schematic of fiber arrangement and abrupt changes in the fiber direction (cp. Spach et al. 1982).^48^ (Right panel) Fiber orientation in the PLA with multiple fiber crossings (Figure adapted).

### Episodes in the Development of Stable Reentry and Slow Conduction Characteristics

The activation patterns necessary for the induction of a stable reentry based on high-resolution mapping remained unclear. Supplemental Data Figure S27 (all times) and Figure 3A) (parts) show a representative example of the development stages of rotational activity over time consisting of varying activation pattern episodes (heart #7,region RAA): The first episode is a repetitive planar wave activation (0-570ms). Then a time episode of repetitive focal wave activation (600–1830ms) continues, which importantly induces a first instable reentrant loop (1860-1890ms). Importantly this reentrant loop resulted in chaotic activations over 210ms with multiple small slow conduction zones (Figure 3A, time 1920-2010ms), which then induced stable rotational activity (2070-4470ms). The activation time maps, trajectory, and activation delay of earliest and latest activation in 2 reentrant loops of this stable reentry are shown in Figure 3B. We further assessed to characterize these slow conduction regions which are responsible for the anchoring of reentrant drivers. Figure 3C) exemplarily shows the activation delay over time in eight following reentrant loops (heart #7 in the RAA). Importantly activation delays in reentrant loops were similar over time in all bi-atrial regions (Figure 3D).

**Figure 3:**
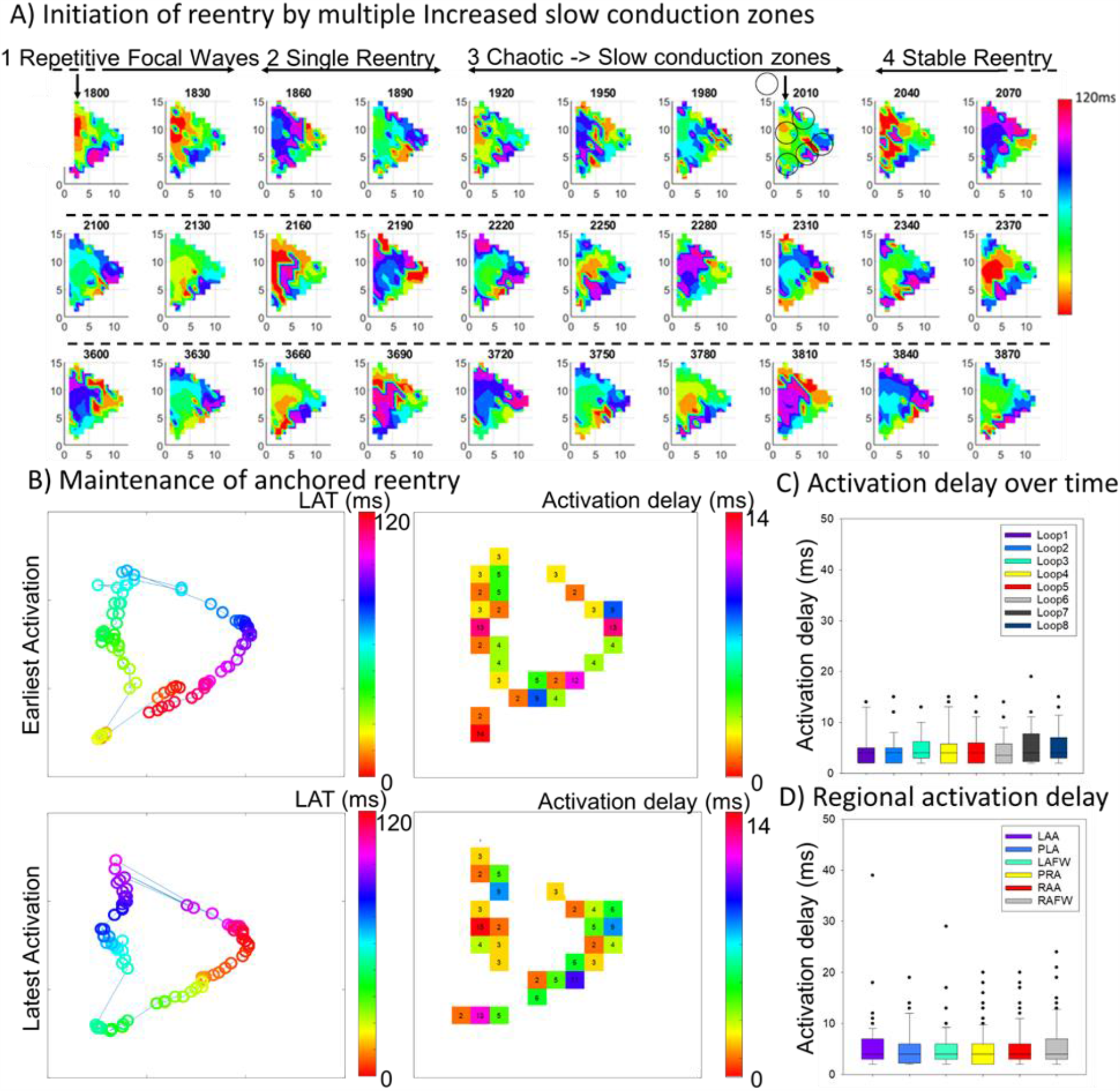
Initiation and maintenance of rotational activity between small slow conduction zones. **A**, A representative example of the development stages of rotational activity over time consisting of varying activation pattern episodes in Heart #7 in the region RAA: 1) Focal wave activation (1800 – 1830ms (further LAT maps of repetitive focal wave activation (600 – 1830ms) are given in the compare Supplemental Data)). 2) First instable reentrant loop (1860, 1890 ms). 3) Chaotic activations (1920-2010 ms). 4) Stable rotational activity (2040 - 3870ms). **B**, Activation time maps, trajectory, and activation delay of earliest activation time in 2 reentrant loops. **C**, Activation delay over time in 8 following reentrant loops. **D**, Activation delay in the 6 different regions in reentrant loops of all animals.

### Termination of Stable Reentry and Overlapping Slow Conduction Regions during Reentry and Planar Wave Activity

Episodes of stable reentries were frequently observed over time. However, the reason for the abrupt ending of reentries in high-resolution mapping remained unclear. We, therefore, analyzed the CL variations during and before the termination of stable reentry. We found that in stable reentries spontaneous increases in CL occurred before the termination of the rotations. Figure 4A) shows an example of a stable reentry (2 seconds). After the increase in CL from 120ms to 150ms, the reentry terminated then resulted in aplanar wave activation. Activation patterns in AF are complex. In this study, episodes of different activation patterns including rotational, focal activity, complex activation pattern, and normal nearly planar wave activations varied over time in the same region. Figure 4B shows a representative example of the RAA (heart #10) of varying episodes of rotational activity, and episodes of more organized nearly planar wave activation (Figure 4B) in the same region. Importantly, the zones of slow conduction during the reentrant activity were at the same locations also present during planar wave activation (marked in black in Figure 4B) and during sinus rhythm (marked in black in Figure 4C). Figure 4B) exemplarily shows a selected bipolar EGM from the rotational activation episode (Figure 4B, upper right panel) and the more organized activation episode (Figure 4B, lower right panel). Activation delay increased by the factor 3-4 during AF compared to sinus rhythm or planar wave activation. Regional cycle length variation shown as the standard deviation of CL was higher in the right atrium compared to the left atrium (Figure 4D). Conduction delay zones were a cycle length dependent phenomenon. Activation delay decreased significantly with increasing CL, during AF and SR/paced rhythm with pacing intervals (200ms to 500ms) (Figure 4E).

**Figure 4:**
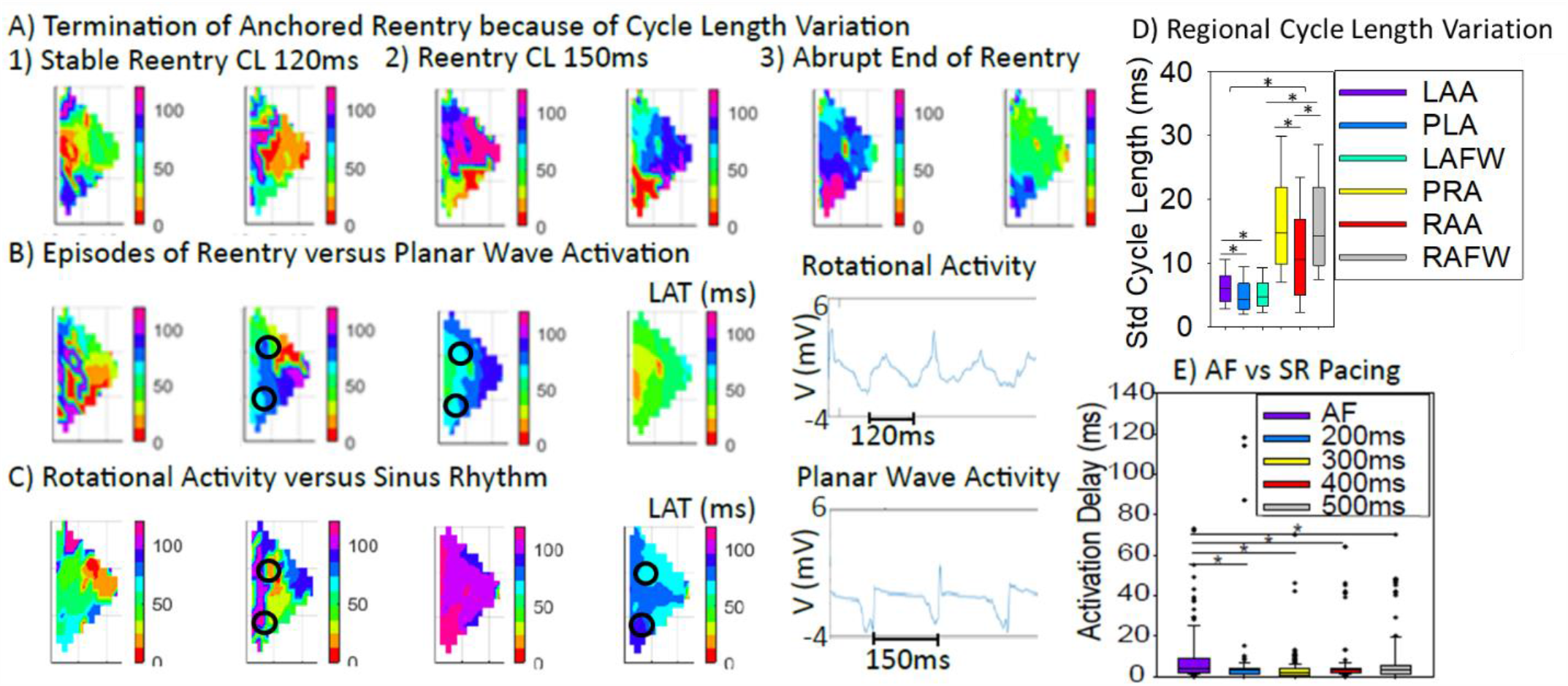
Termination of rotational activity because of CL prolongation and similar slow conduction zone locations in AF and SR. A representative example of cycle length variation and the termination of stable reentry. Rotational activity with increasing CL over time in the RAA in heart #10. (1) CL of 120 ms changed to (2) CL of 150 ms after 1.5 seconds rotational activity. After CL prolongation (3) rotational activity abruptly ended. B, A representative example of rotational activity (CL=90 ms) in LAT maps (30 ms steps) in the RAA in Heart #10. (Left panel) The episode of rotational activity. (Right panel) More organized nearly planar wave episode (CL=120 ms) in the same region (RAA) and animal. Slow conduction at the same location during rotational activity and planar wave activation. C, Overlapping slow conduction zones during AF (top panel) and sinus rhythm in the PLA. Activation delay increased by the factor 3-4 during AF compared to sinus rhythm or planar wave activation at slow conduction zones. CL increased from 120ms to 150ms (compare signal recordings). D, regional cycle length variations (standard deviation of CL) with the highest CL variations in the right atrium. E, Decrease of activation delay (2x) when comparing AF with sinus rhythm/ pacing intervals (200ms to 500ms). The pacing was performed from the LAA.

### Thin Line Activation, Line of Block, Figure-of-Eight Activation

The activation patterns focal activity, epicardial breakthrough, and line of block were reported in previous studies in high-density epicardial AF mapping. ^33^ In this study, we further found a new activation pattern ‘thin line activation’, (Figure 5A). We defined ‘thin line activation’ as simultaneous earliest activation (red in activation time maps) along thin line regions on the epicardial surface. Importantly these thin line activations often closely juxtaposed to activation patterns such as focal activity and lines of block (Figure 5B). Figure 5B shows focal activity as simultaneous activation between lines of block, the locations,which were consistent over time episodes in LAT maps. Also, focal activities between the lines of block were frequently observed at the same locations (Figure 5 B-C). Figure 5C demonstrates a figure-of-eight activation along thin activation lines in the PRA (heart #5) frequently observed at the same location with the dimension of the figure-of-eight reentrant trajectory of 7.5mm x 15mm. Importantly most figure-of-eight-activations with small circuit dimensions were located in the PLA. Further, the detected focal source dimensions measured with the length and width of the simultaneous earliest activations in 5ms isochrones and the number of focal sources per AF cycle are given in Figure 5D. The detected lengths and widths of the simultaneous activations were (4.9±5.0 mm, and 4.9±6.2 mm), but the range of dimensions between 1-30 mm also led to thin line activations (Figure 5 B-C).

**Figure 5:**
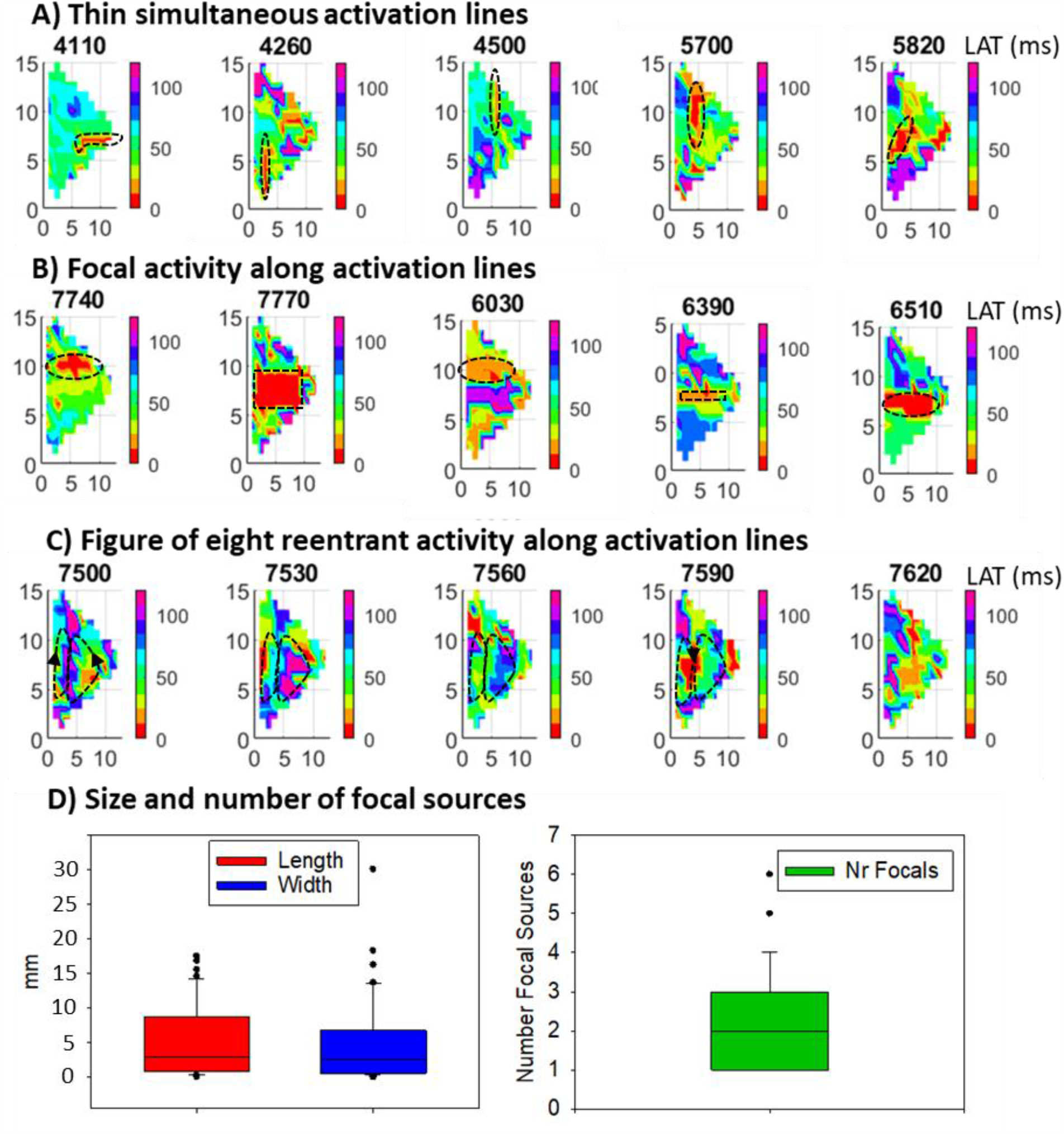
Different activation patterns along with activation lines. **A**, A representative example (heart #5) of thin simultaneous earliest activation (red in activation time maps, marked in black dashed line) along lines (size up to 2.5 x 25 mm) on the epicardial surface. B, Focal activity as simultaneous activation between activation lines, which were similar located (block dashed regions) over time episodes in LAT maps. Focal activities between line of blocks (up to 25mm) were frequently observed at the same locations potentially because of simultaneous activation in fiber lines. C, Figure of eight activation along activation lines in the PRA in heart #5 frequently observed at the same location (marked with black dashed lines). D) Dimension (length and width) of focal sources detected as simultaneous earliest activation in 5ms and number of focal sources per AF cycle.

### Correlation of Fiber Crossings with Stability of Reentry

We further analyzed the underlying substrate characteristics in all bi-atrial regions in six animals (Figure 6). Figure 6A shows the exemplary tissue sections in all regions in heart #7. The highest degree of foal fibrosis/ fat was detected in the PLA (49±14%, median 42%) and in the LAFW (47±19%, median 50%) (Figure 6B). There was no significant regional difference in fiber anisotropy detected as the standard deviation of fiber orientation (measured in degree) (Figure 6C). But the fiber crossings density was significantly higher in the PLA, LAA, and in the right atrium in the RAA (Figure 6). However, there was no significant correlation between fiber orientation and stability of rotational activity. In contrast, the stability of rotational activity correlated with the number of fiber crossings per tissue section (R=0.6, P<0.05) (Figure 6E). Furthermore, the slow conduction regions were located at fibrotic/ fat tissue regions in histological tissue sections (>=3mm) and low voltage regions (<1mV) (Supplemental Data Figure S23). Slow conduction may be the product of increased fiber crossings as well as more fibrosis/fat. Interestingly nearly no dense fibrosis/ fat zones were detected in the appendages, but about 1.5 fibrosis/ fat regions per tissue section were detected in the remaining bi-atrial regions (Figure 6F). Importantly, more than 90% of the detected fibrosis and fat regions were located near fiber crossings in the atrial regions (Figure 6G).

**Figure 6:**
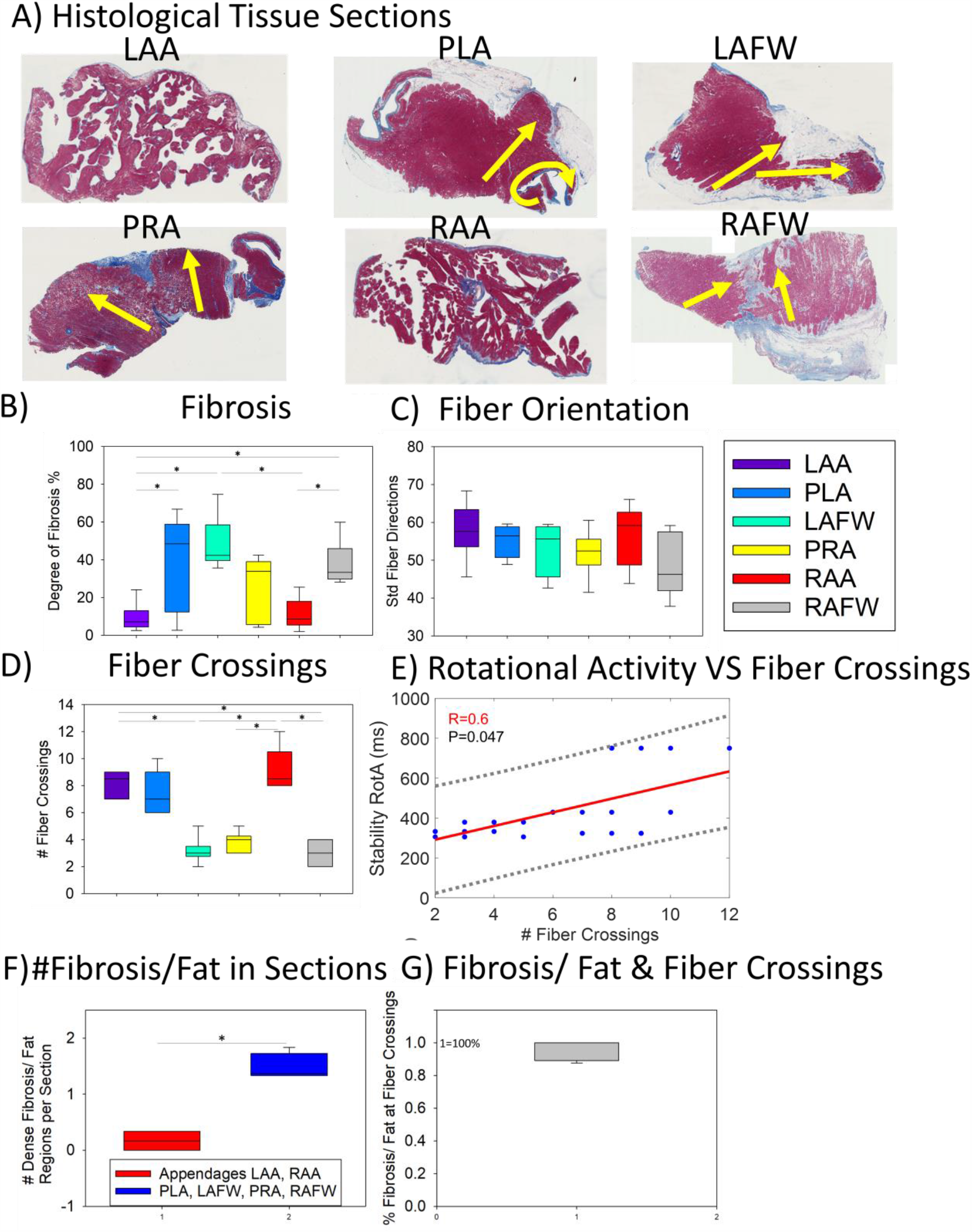
Substrate characteristics and correlation of rotational activity with fiber crossings density. A, Masson’s trichrome stained tissue section. in the 6 bi-atrial regions: left atrial appendage (LAA), posterior left atrium (PLA), left atrial free wall (LAFW), posterior right atrium (PRA), right atrial appendage (RAA), right atrial free wall (RAFW). Fiber orientations are marked in yellow. Dense focal fibrosis developed at fiber crossings (intersections of fiber lines). B, Degree of focal fibrosis in all bi-atrial regions. The highest focal fibrosis was detected in the PLA and LAFW. C, Fiber isotropy index in all bi-atrial regions. The highest anisotropy was detected in the PLA, LAA, and in the right atrium in the RAA (P-value non-significant). D, The highest number of fiber crossings were detected in the RAA, LAA, and PLA. E, Correlation of fiber crossings with the stability of rotational activity. F, Number of detected dense fibrosis/ fat regions in each tissue section. Nearly no dense fibrosis/ fat regions were detected in the appendages, but about 1.5 in the other atrial regions. G, More than 90% of the detected fibrosis and fat regions were located near fiber crossings in the atrial regions. P < 0.05 for all comparisons.

### Correlation of Rotational Activity Stability with Electrogram Measures

We further correlated different established electrogram measures with the detected stability of rotational activity in this study. Importantly, stability of rotational activity correlated with electrogram measures OI (R=0.68), FI (R=0.61), ShEn (R=0.58) and CL (R=0.56), but not with DF (R=0.28), (p < 0.0001, for all comparisons) (Supplemental Data Figure S52). The stability of rotational activity correlated best with the electrogram measure OI.

### PVI Reduced AF Drivers in Remote Regions to the PVs

Pulmonary vein isolation is thought to be efficacious in patients with paroxysmal and persistent atrial fibrillation. However, the underlying mechanisms by which PVI leads to beneficial atrial remodeling are not clear. We hypothesized that PVI affects rotational activity and slow conduction zones in regions close to and remote to the PVs. We further hypothesized that a partial mechanism may be CL prolonguation, which improves conduction (i.e. decreases slow conduction zones) and therefore decreases the stability of anchored rotational activity. To assess the beneficial effects of PVI on AF, we performed epicardial pulmonary vein isolation in five hearts. PVI led to a significant increase in cycle length (range: 2.3%-14.1%) (Figure 7A) and decreased stability of rotational activities in nearly all atrial regions remote from the site of ablation (Figure 7B). Slow conduction zones were similarly located pre and post PVI (Supplemental Data Figure S53). Importantly the percentage change of slow conduction zones pre vs post-PVI was highest in the right atrial appendage (50% reduced slow conduction zones) (Figure 7C) and correlated with the stability of rotational activity at baseline (Figure 7D). PVI decreased rotational driver stability throughout the atria, by affecting the extent of conduction slowing at ‘slow conduction’ zones also in remote regions to the PVs. The extend of slow conduction further correlated with the fiber crossing density in the histological tissue analysis (Figure 7E).

### Patient Characteristics and Evaluation of AF Drivers and Slow Conduction in Patients

We further included nine patients (age 62.8±9.2, male 75%) in early persistent AF in the clinical study. The majority of the patients had bi-atrial dilatation (75%) with a mean left atrium volume of 67.1±45.9 body surface area. The mean AF cycle length was 146±3 ms (median 134.2). The clinical patient characteristics of the study population are summarized in Table 2. We evaluated slow conduction at reentry sites and at focal sources near the pulmonary veins in 9 patients using the high-definition (HD) mapping catheter with 16 equidistant electrodes (HD Grid Mapping Catheter Sensor Enabled, Abbott Technologies, Minneapolis, MN) (Figure 8A). Similar to the findings observed in the rapid atrial pacing model in animals, rotational activity trajectories had maximal activation delay zones at the edges of the reentrant circuits. The activation time map of 1 AF cycle in the PLA (Figure 8B) shows a reentrant circuit with the 2 to 6 fold higher activation delay at one edge of the reentrant trajectory compared to the other reentry trajectory segment parts. Figure 8C) exemplarily shows an activation time map during CS-pacing. We detected 2.3±1.6 (median 2) reentries and 2.1±1.1 (median 2) focal sources per AF cycle in patients in the left atrium (Figure 8D). Activation delay and maximal activation delay at focal sources were 20.2±19.1 ms (median 15.7 ms) and 26.8±17.4 ms (median 24.4 ms). Activation delay and maximal activation delay at reentries were 9.3±8.8 ms (median 9.2 ms) and 20.2±8.1 ms (median 19.6 ms) along 2.5 mm (Figure 8E). The activation delay was 2x higher in AF compared to SR/ CS-pacing patient maps (Figure 8F). Figure 8G presents an exemplarely bipolar voltage map and the corresponding activation time map in the Ensite NavX mapping system ® (Abbott) (Figure 8H).

**Table 2.**
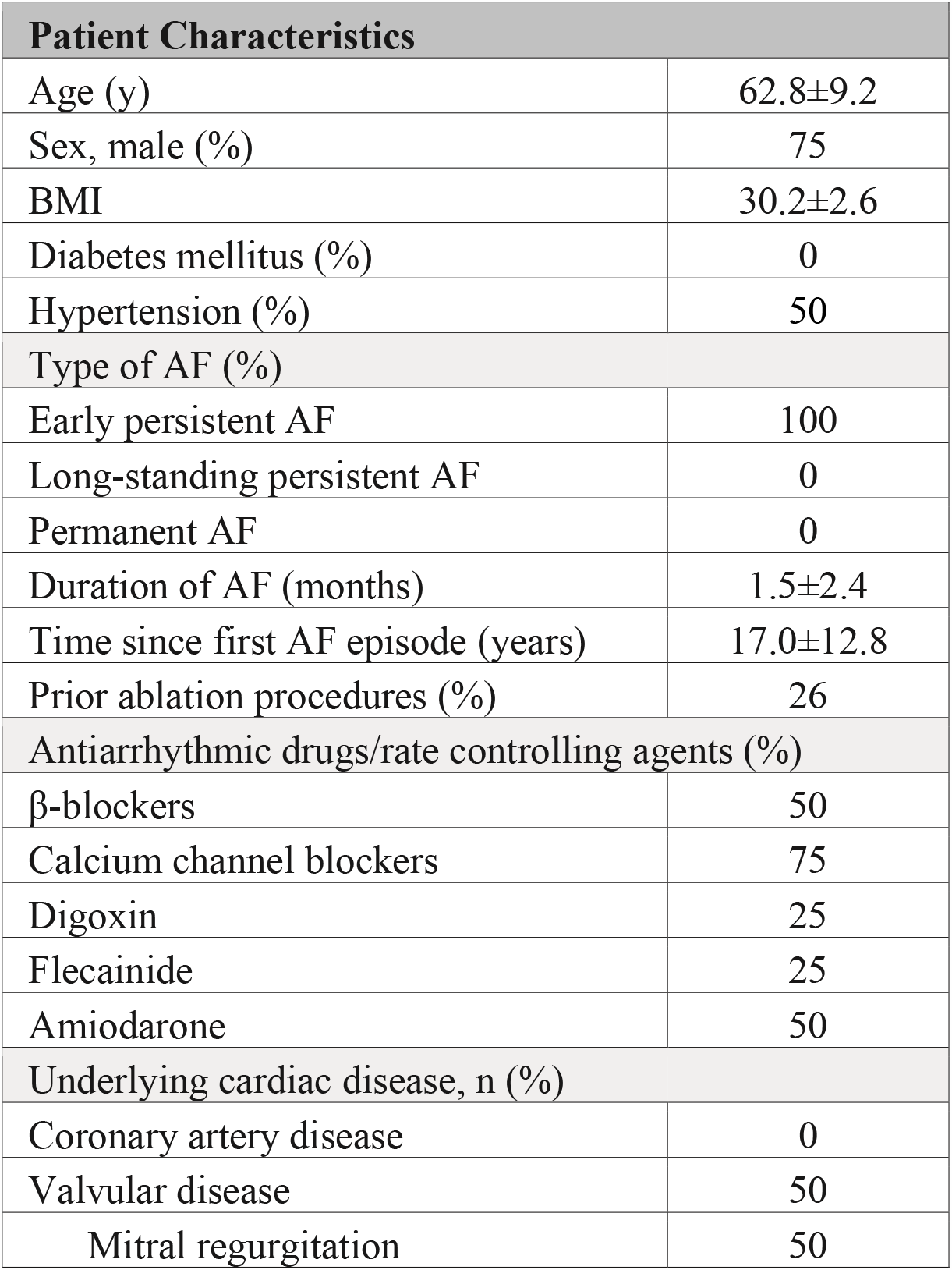
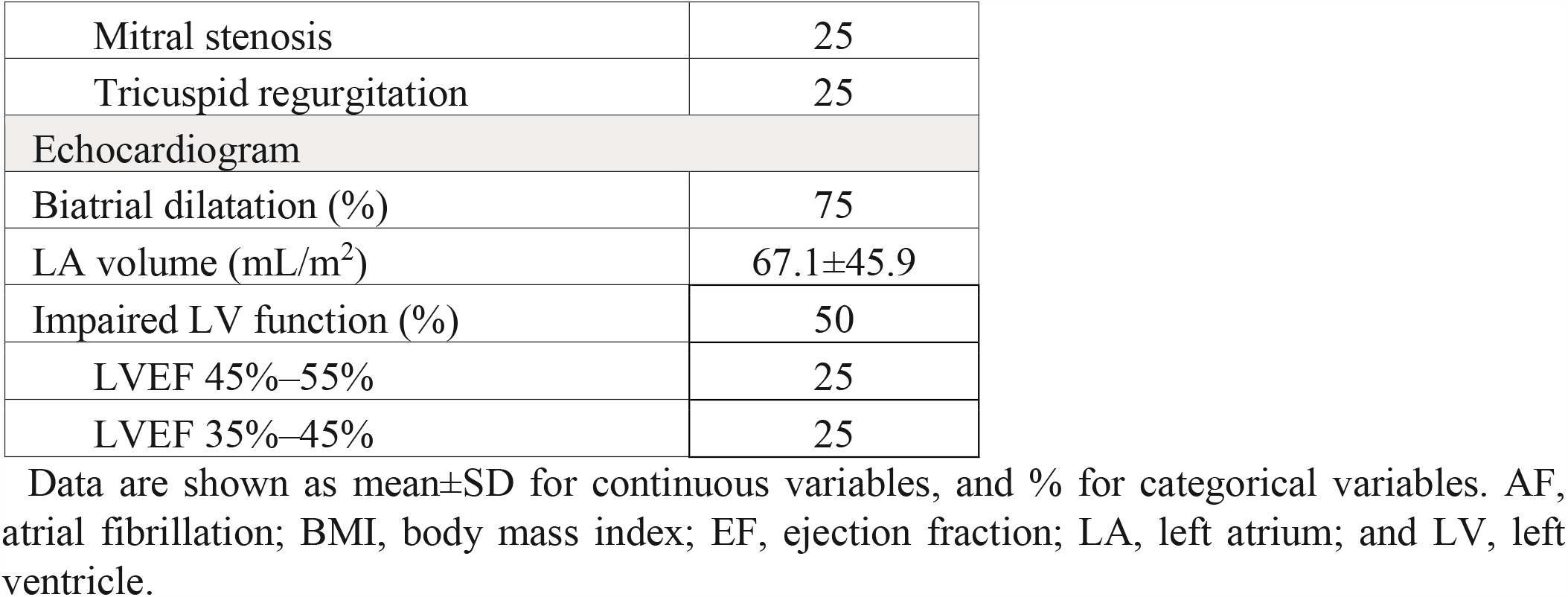
Patient Characteristics.

| Patient Characteristics |  |
| --- | --- |
| Age (y) | 62.8±9.2 |
| Sex, male (%) | 75 |
| BMI | 30.2±2.6 |
| Diabetes mellitus (%) | 0 |
| Hypertension (%) | 50 |
| Type of AF (%) |  |
| Early persistent AF | 100 |
| Long-standing persistent AF | 0 |
| Permanent AF | 0 |
| Duration of AF (months) | 1.5±2.4 |
| Time since first AF episode (years) | 17.0±12.8 |
| Prior ablation procedures (%) | 26 |
| Antiarrhythmic drugs/rate controlling agents (%) |  |
| β-blockers | 50 |
| Calcium channel blockers | 75 |
| Digoxin | 25 |
| Flecainide | 25 |
| Amiodarone | 50 |
| Underlying cardiac disease, n (%) |  |
| Coronary artery disease | 0 |
| Valvular disease | 50 |
| Mitral regurgitation | 50 |
| Mitral stenosis | 25 |
| Tricuspid regurgitation | 25 |
| Echocardiogram |  |
| Biatrial dilatation (%) | 75 |
| LA volume (mL/m<sup>2</sup>) | 67.1±45.9 |
| Impaired LV function (%) | 50 |
| LVEF 45%–55% | 25 |
| LVEF 35%–45% | 25 |

Data are shown as mean±SD for continuous variables, and % for categorical variables. AF, atrial fibrillation; BMI, body mass index; EF, ejection fraction; LA, left atrium; and LV, left ventricle.

## Discussion

### Are Reentries Frequently Anchored between Slow Conduction Zones in All Bi-Atrial Regions?

A detailed analysis of rotational activities in both atria, including the appendages based on high-resolution contact mapping, has not been systematically performed in prior studies. In the current study, our results demonstrate that potential AF drivers were likely the result of activation wavefronts emanating from interacting rotational and focal activity in all 6 analyzed bi-atrial regions. The rotational activity, automatically detected with a novel algorithm based on the earliest and latest activation, was most stable in the appendages and PLA and anchored between small slow conduction zones with activation delay > 10ms. In this work, we also observed changing beat-to-beat wavefronts and varying spatiotemporal behavior of driver activities. Reentrant activation was not sustained (median, 2.6 rotations lasting 449±89 ms), meandered, and recurred repetitively in the same region between (n>=3) small slow conduction zones.

Several previous studies showed PV-triggers and that AF triggers outside the PVs were located in the vena cavae, the crista terminalis, the coronary sinus, the ligament of Marshall, the inter-atrial septum, and the appendages.^34-42^ The role of the PVs in the development and maintenance of AF was published by Haïssaguerre et al.. Narayan et al, detected focal and rotor activity with spatial stability using low-resolution endocardial basket catheter and phase singularity AF mapping.^16^ Recently, Rudy et al, identified in a noninvasive mapping study in paroxysmal and persistent AF patients multiple (2–5) concurrent wavelets (92%), with both simultaneous focal activation from areas near the pulmonary veins (69%) and non-pulmonary veins (62%), but reentry which sustained >1 rotation was rarely seen. ^14^ In contrast, Haissaguerre et al showed AF drivers in a noninvasive mapping study with 80.5% reentries and 19.5% focal breakthroughs with incessantly changing beat-to-beat wavefronts and spatiotemporal behavior.^43^ de Groot et al. showed in high-density sequential epicardial mapping in patients with AF epicardial breakthrough sites some intermittent focal activity (0.8%).^33^ Recently, Kalman et al., showed, using sequential, epicardial LA and RA mapping in patients with persistent AF, intermittent foci (≤2 beats), and intermittent reentry (≥2 rotations), but neither sustained focal nor sustained reentrant activation was detected. ^18^ However, in this study, we found the most stable rotational activities pre and post PVI in the RAA and also in the LAA in which AF drivers were more stable compared to most other regions. Based on the findings in this work, electrical isolation of the appendages in addition to PVI may further improve the success rate of freedom from AF compared to PVI only. A previous clinical study showed that the left atrial appendage may be an underrecognized trigger site of atrial fibrillation.^44^ Furthermore, Natale and colleagues showed that empirical electrical isolation of the left atrial appendage improved long-term freedom from atrial arrhythmias.^45^In a comparison of our study to previous reports, the finding of rotational driver activation during AF is consistent, in whole or in part, with the findings of others. ^7, 8, 10, 11, 18, 33^ However, importantly, our data further characterized AF drivers with slow conduction regions in bi-atrial regions.

### Novel Structural And Functional ‘Polygon Reentry Model’ of Anchored Reentry between Fiber Crossings -

Almost 100 years ago, Garrey and Lewis first hypothesized that re-entry could sustain and drive AF. ^46^ Allessie et al. published in 1973, that reentry may be functional and not require an anatomically defined pathway.^47^ In this work, we demonstrated that the reentrant circuit trajectories were more complex than circles or ovals like those previously described and followed line patterns with a curvature angle between lines, which appeared to be at last partially anchored to myofiber crossings or regions of fibrofatty infiltrations. The velocity of stable rotational activities along the described line patterns with curvature propagation angle was 4 x slower at the edges of the trajectory compared to the propagation lines. These slow conduction zones appeared to be located largely over crossing fibers, fibrosis, or fat tissue zones in the histological tissue section analysis. This finding indicates that regions of fibrofatty infiltrations may at least partially develop at fiber crossings, with the densest fibrofatty infiltrations in the PLA near the pulmonary veins and in the LAFW. Furthermore, the stability of rotational activity correlated with the number of fiber crossings. Several previous studies suggest that fiber orientation distribution is an important factor in the development of AF. However, in this study, we detected only a trend between fiber orientation distributions measured with the standard deviation of fiber orientations and stability of rotational activity. In contrast to previous studies, we analyzed for the first time also the number of fiber crossings per tissue section in different biatrial regions. Importantly, the number of fiber crossings were more specific and correlated with the stability of rotational activity.

Previous studies demonstrated that the main contributors to conduction and slow conduction in the heat are: a) fiber orientation, b) Na channel characteristics, c) gap junction characteristics, and d) the presence of fibrosis. Spach et al, demonstrated that conduction delay occurs at zones that reveal sudden changes in the fiber direction.^48^ These sites also have an abrupt increase in axial resistivity in the direction of propagation ensued, which may result in a decrease of the safety factor for propagation. Hocini et al., 2002 further demonstrated that zones of activation delay of up to 120 ms over distances as small as 3 mm in canine pulmonary veins correlated with abrupt changes in fascicle orientation. ^49^ The architecture of muscular sleeves in the PVs may facilitate reentry and arrhythmias associated with the ectopic activity. In this RAP animal model, activation delays, within 2.5 mm x 2.5 mm was > 4 times larger compared to normal activation and activation frequently curved around slow conduction zones, which predisposed to reentry. In patients, reentries and focal sources anchored at zones of activation delay up to 90 ms within 2.5 mm using the HD grid catheter near the PVs. Also in the clinical data, activation delay at the reentrant trajectory was about 2x higher at edges of the trajectory and 1.4 higher during AF compared to SR/ CS-pacing similar to the animal data in this work. Furthermore, these findings are in line with observations from Hanse and colleagues, who showed that stable reentries anchored at regions of increased fiber angle (epicardial 37 degrees) and interstitial fibrosis (12.8 ± 1%) and made repetitive a U-turn at micro-anatomic tracks (2 mm).^50^ However based on optical mapping restrictions their human data study of AF drivers was limited to the lateral right atrium. Our findings of slow conduction sizes and activation delays are similar to observations from Spach et al. 1982, Hocini et al., 2002 and Hanse et al. 2015 ^48-50^ Furthermore, we found in this work, slow conduction regions and anchored reentries around these slow conduction regions in all 6 analyzed bi-atrial regions.

The reentry models described in the literature in the last decades like circus movement, leading circle, spiral wave, and the new proposed model of reentry along line patterns in the form of a polygon are given in Figure 9A. In the circus movement reentry model, the anatomic reentry is around a fixed obstacle and allows for an excitable gap. In the leading circle reentry model, the conduction velocity (CV) and refractory period (RP) determine the circuit size. Also, the center of the circuit is maintained refractory by centripetal wavelets. The spiral wave model describes a curved wavefront with a core the so-called phase singularity (PS) which rotates around an excitable but unexcited core. In contrast to the previous models, the new model describes both structural and functional characteristics with slow conduction zones at the edges of propagation line patterns. Previous models focused on the tip of the rotational activity analyzing the core with phase singularity points in phase maps. ^20^ In contrast, the focus in the new model is the earliest and latest activation over time and slow conduction determined with activation delay within activation time maps. The main findings of this study are given in summary in Figure 9B. Small conduction zones (>3mm) may be at least partially myofiber crossings or regions of fibrofatty infiltrations in all bi-atrial regions. At these fiber crossings zones, focal fibrosis and fat tissue were detected and may develop over time at these zones. These fiber crossing zones were frequency-dependent zones of activation delay and rotational activity frequently anchored between these small slow conduction zones. These zones were further located at the same locations in different rhythms or activation patterns (AF and sinus rhythm, rotational activity, and planar wave activation). Importantly, activation delay increased at these small conduction zones during rotational activity and decreased during planar wave activation or sinus rhythm. PVI increased CL and reduced the activation delay and stability of rotational activity in nearly all regions remote to the PVs. These activation delay zones at rotational activities were present in the animal model and patients. Importantly, these findings result in a proposed new model of reentry. In contrast to the previous models, the new model describes both structural and functional characteristics with slow conduction zones at the edges of propagation line patterns. In contrast to previous reports, this study provides a detailed mapping of all subregions in both atria, and a new slow conduction model of reentry. Furthermore, this work demonstrates a clear dependence of slow conduction on cycle length (both structural and functional), new human data,using HD grid, that shows parallels between animal and clinical activation maps/reentry/focal activity and the effect of PVI on rotational activity/conduction characteristics ‘at a distance’ to the PVs.

**Figure 7:**
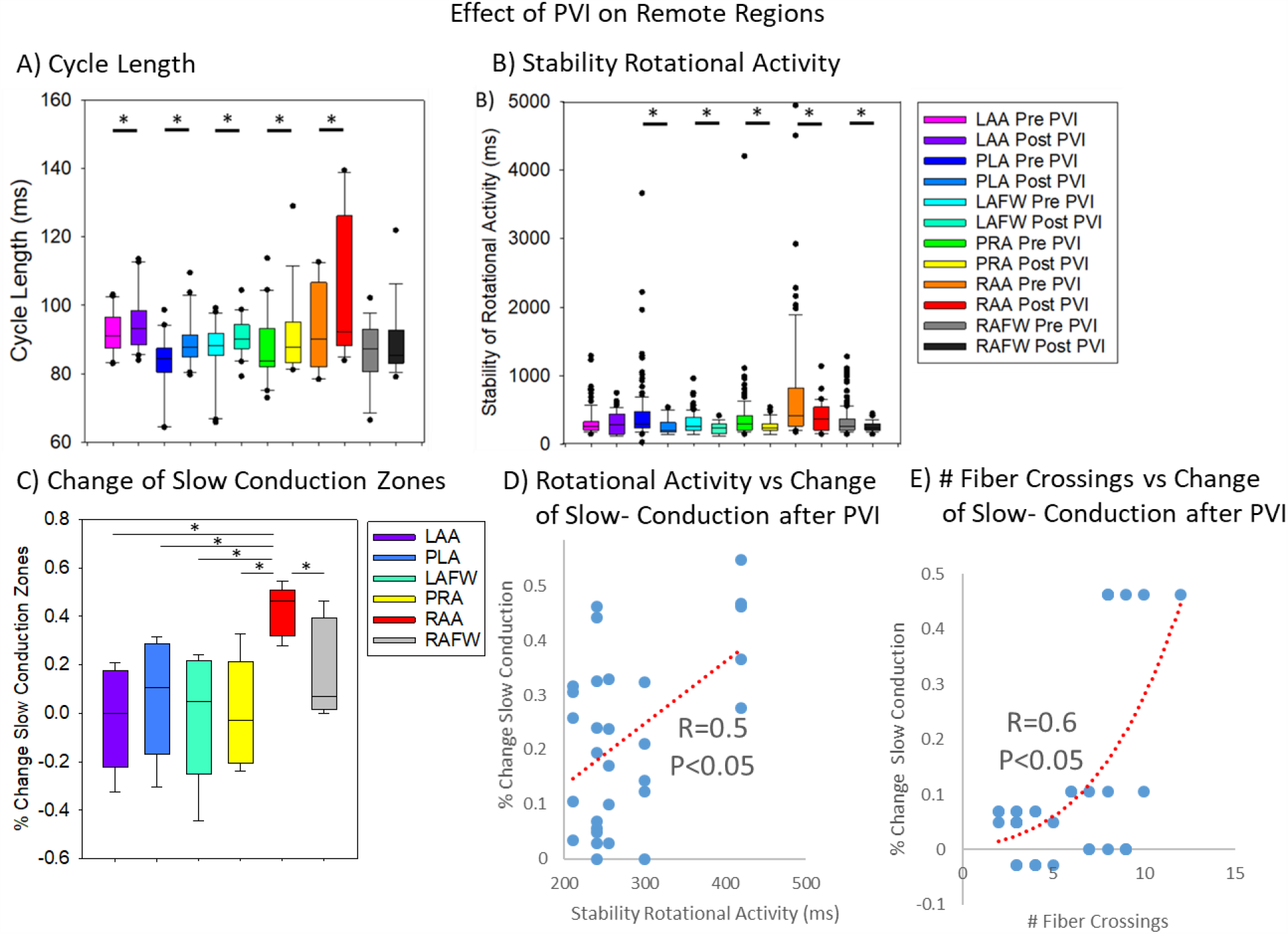
Effects of PVI on Regions remote to the PVs. A, PVI led to a significant increase in CL in nearly all regions remote to the PVs. B, Stability of rotational activity decreased significantly (baseline vs after PVI) in nearly all atrial regions. C, Change of slow conduction zones (>10ms activation delay) pre vs post PVI in %. D, The percentage reduction change (range 0 - 60%) of slow conduction zones pre vs post PVI correlated with the stability of rotational activity at baseline (R=0.5, P=0.005). E, Number of fiber crossings per tissue sections vs change of slow conduction after PVI.

**Figure 8:**
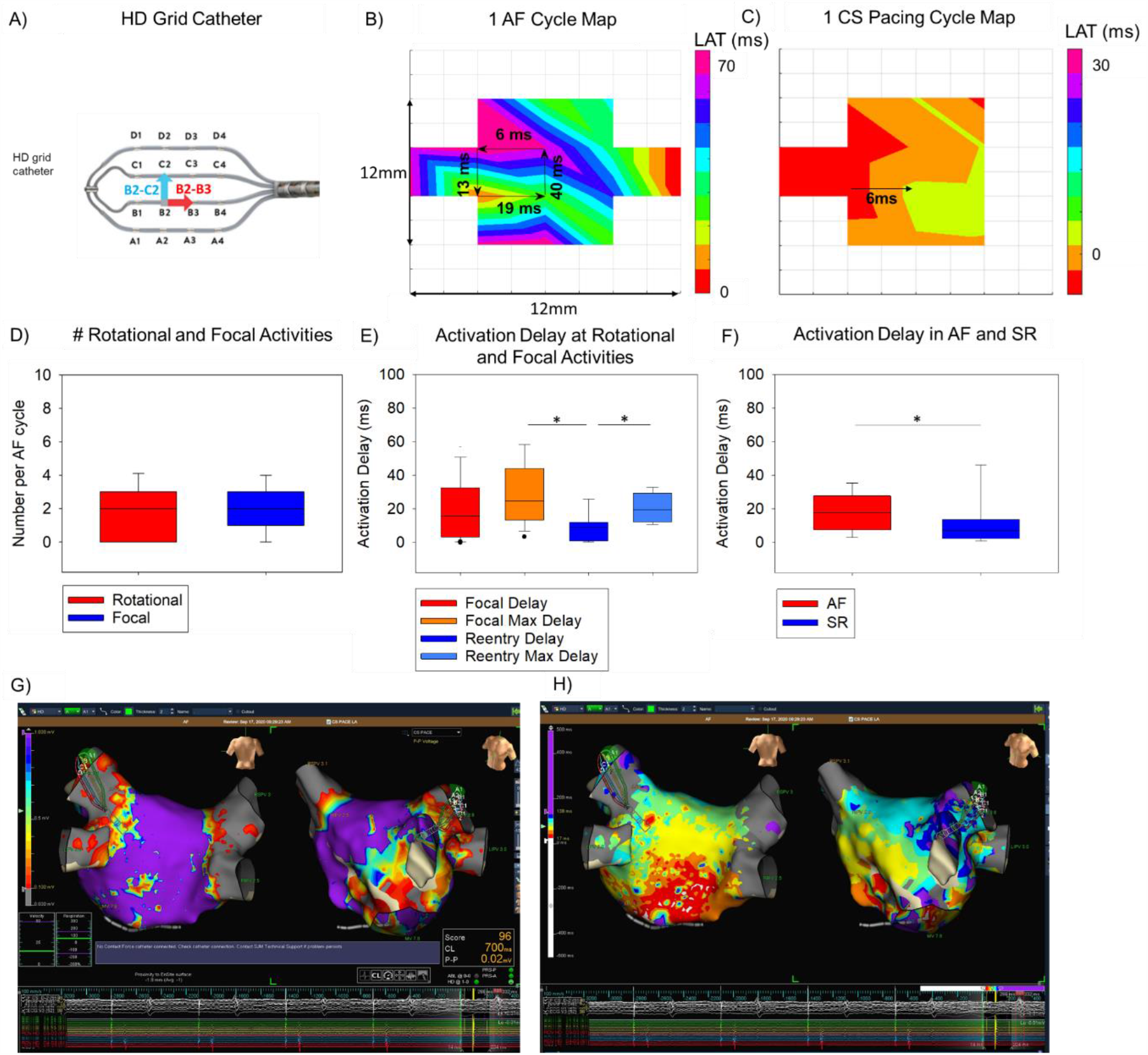
Clinical Evaluation of AF Drivers and Slow Conduction Zones in the LA in Persistent AF Patients. A, Clinical left atrial evaluation of AF drivers and slow conduction zones in persistent AF patients using the clinical A) HD-grid catheter with 16 electrodes (interelectrode distance of 2.5 mm). 24 bipolar signals recordings between the 16 electrodes of the HD grid catheter. Vertical bipolar recording marked in blue (y-direction). Horizontal bipolar electrodes marked in red (x-direction). B, Activation time map of 1 AF cycle showing a reentrant circuit with increased activation delay (40ms) at one trajectory segments (2-6 fold higher compared to the other 3 trajectory segment parts). C, Activation time map of 1 CS-pacing measurement with the HD-grid catheter showing about 2x smaller activation delays compared to AF. D, Analysis of detected reentries, focal waves per AF cycle. E, Analysis of activation delay and maximal activation delay at focal sources 20.2±19.1 ms (median 15.7 ms) and 26.8±17.4 ms (median 24.4 ms) and activation delay and maximal activation delay at reentries 9.3±8.8 ms (median 9.2 ms) and 20.2±8.1 ms (median 19.6 ms) and. F, Activation delay in AF and SR showing 2x higher activation delay in AF compared to CS-pacing/SR. G, Bipolar voltage map and corresponding H, activation time map in Ensite NavX mapping system ® (Abbott).

**Figure 9.**
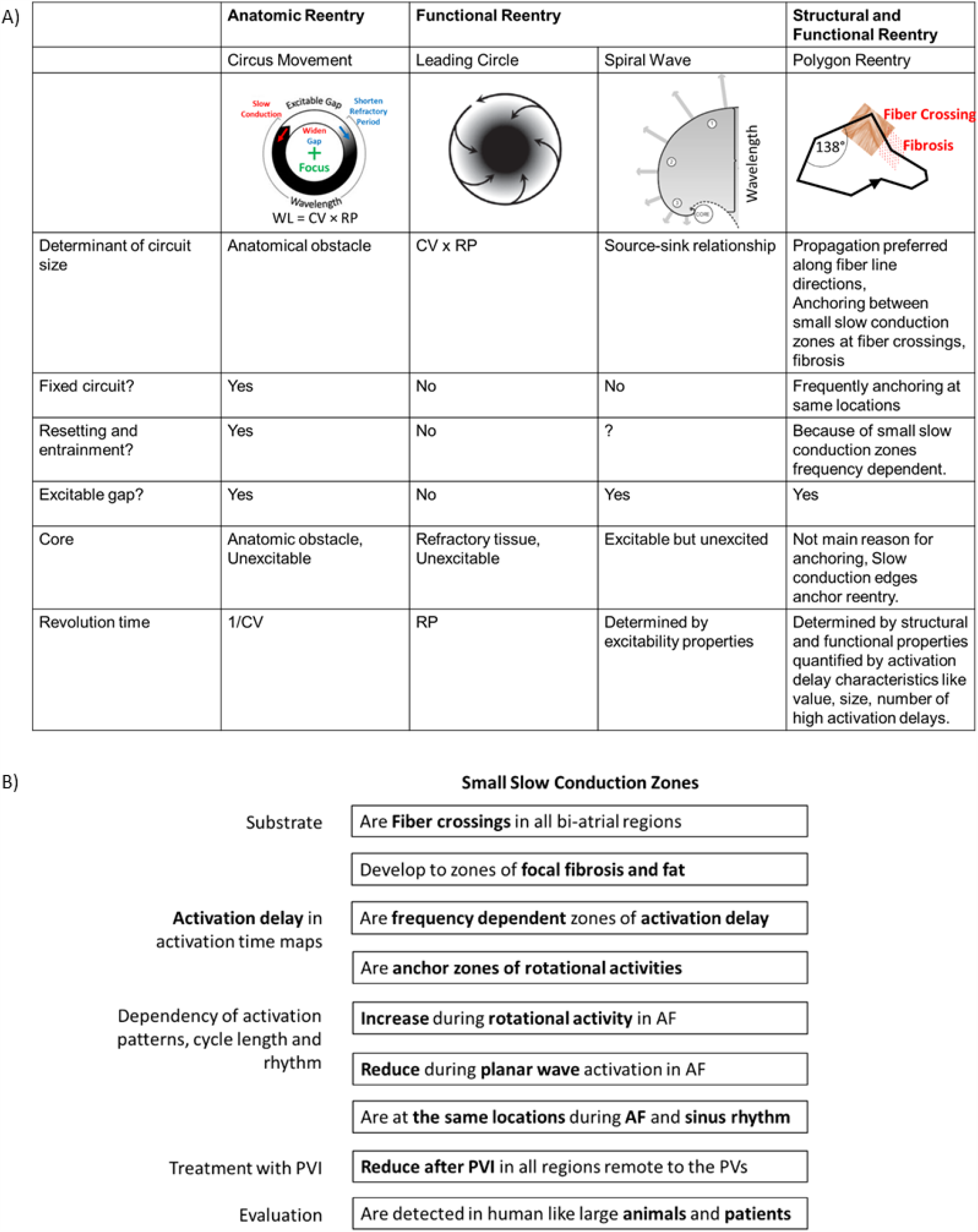
Summary figure of novel reentry model and key findings. A, Reentry models circus movement, leading circle, spiral wave, and new models of reentry along line (polygon) patterns. In the circus movement, the anatomic reentry is around a fixed obstacle and allows for an excitable gap. In the leading circle reentry model, the conduction velocity (CV) and refractory period (RP) determine the circuit size. Also, the center of the circuit is maintained refractory by centripetal wavelets. The spiral wave model describes a curved wavefront with a core the so-called phase singularity (PS) which rotates around an excitable but unexcited core. In contrast to the previous models, the new model describes both structural and functional characteristics with slow conduction zones at the edges of propagation line patterns. Previous models focused on the tip of the rotational activity analyzing the core with phase singularity points in phase maps. The focus in the new model is in contrast to the previous models the earliest and latest activation within activation time maps. B, Summary figure of this study. Small slow conduction zones (>3mm) were zones of fiber crossings in all bi-atrial regions. At these fiber crossings zones, focal fibrosis and fat tissue developed. These fiber crossing zones were frequency-dependent zones of activation delay and rotational activity anchored between these zones. These zones were located at the same locations in different rhythms or activation patterns (AF and sinus rhythm, rotational activity, and planar wave activation). Activation delay increased at these zones during rotational activity and decreased during planar wave activation or sinus rhythm. PVI increased CL and reduced the activation delay of these zones in all regions remote to the PVs. These activation delay zones at rotational activities were present in the animal model and patients. These findings result in a proposed new model for reentry. In contrast to the previous models, the new model describes both structural and functional characteristics with slow conduction zones at the edges of propagation line patterns. Previous models focused on the tip of the rotational activity analyzing the core with phase singularity points in phase maps. The focus in the new model is in contrast to the previous models the earliest and latest activation within activation time maps.

### Implications

Our data describe a new paradigm for the mechanism that induces and maintains persistent AF. Understanding the mechanism of reentry between small slow conduction regions presents opportunities for new potential approaches to the treatment of persistent AF, especially for ablation. In previous reports, the trajectory of reentries was calculated based on phase singularities.^20, 28, 29^ But, this method is limited in robust reentrant detection and can falsely detect phase singularities in the absence of rotors.^30^ In contrast, this work presents a completely new method for detecting the trajectory of rotational activity more robustly (30% more accurate) based on earliest, latest activation, and activation delay. This new method may be integrated into future clinical systems to provide physicians a robust detection of AF drivers in addition to clinical relevant slow conduction regions. Current success rates in persistent AF have been far from optimal, and actual treatment approaches remain largely empirical because we have not understood well enough the mechanism(s) maintaining persistent AF. The study of the mechanism(s) maintaining AF has been limited by low resolution, and the difficulty in mapping this complex arrhythmia. The data from this work may provide opportunities for a targeted, rather than an empirical approach to treat AF.

### Study Limitations

The findings cannot be assumed to apply to paroxysmal AF because our mapping study was only performed in hearts with persistent AF. In this study, we focused on rotational activities. Further studies of focal sources and breakthrough activities and subepicardial reentrant circuits need to be performed. To prove that drivers maintain AF, first putative drivers need to be identified, and then it needs to be demonstrated that AF reduced or stopped. However, we performed PVI only in 5 hearts. Further studies with the performance of ablation at additional trigger sites and slow conduction regions need to be performed. Although we detected similar characteristics of AF drivers and slow conduction zones in patients and animals, the values based on animal studies may not apply to the values in humans in all atrial regions. Also, the vulnerable substrate for AF in humans develops over many years, leading to increased structural complexities, compared with months in a dog model.

## CONCLUSION

Rotational activities in all bi-atrial regions anchored between small frequency-dependent activation delay zones in AF. PVI led to beneficial remodeling in bi-atrial regions remote to the PVs. These data may identify a new paradigm for persistent AF.

## Data Availability

Detailed data are available in the attached Data Supplemental.

## Patient Characteristics

Data are shown as mean±SD for continuous variables, and % for categorical variables. AF, atrial fibrillation; BMI, body mass index; EF, ejection fraction; LA, left atrium; and LV, left ventricle.

## ABBREVIATION

ACT: Activated clotting time
AF: Atrial fibrillation
ANOVA: Analysis of variance
CL: Cycle length
CV: Conduction velocity
DF: Dominant Frequency
EGM: Electrogram
FI: Fractionation Interval
LA: Left atrium
LAA: Left atrial appendage
LAFW: Left atrial free wall
LAT: Local activation time
OI: Organization Index
Pi: Probability of an amplitude value occurring in bin i
PLA: Posterior left atrium
PRA: Posterior right atrium
RA: Right atrium
RAA: Right atrial appendage
RAP: Rapid atrial pacing
RAFW: Right atrial free wall
RotA: Rotational activity
RP: Refractory period
PV: Pulmonary vein
PVI: Pulmonary vein isolation
PS: Phase singularity
SD: Standard deviation
SEM: Standard error of the mean
ShEn: Shannon’s entropy

## Literature

1. Moe GK and Abildskov JA. Atrial Fibrillation as a Self-Sustaining Arrhythmia Independent of Focal Discharge. Circulation. 1957;16:917–917.

2. Allessie MA, Lammers WJ, Bonke IM and Hollen J. Intra-atrial reentry as a mechanism for atrial flutter induced by acetylcholine and rapid pacing in the dog. Circulation. 1984;70:123–35.

3. Haissaguerre M, Jais P, Shah DC, Takahashi A, Hocini M, Quiniou G, Garrigue S, Le Mouroux A, Le Metayer P and Clementy J. Spontaneous initiation of atrial fibrillation by ectopic beats originating in the pulmonary veins. N Engl J Med. 1998;339:659–66.

4. Calkins H, Kuck KH, Cappato R, Brugada J, Camm AJ, Chen SA, Crijns HJG, Damiano RJ, Davies DW, DiMarco J, Edgerton J, Ellenbogen K, Ezekowitz MD, Haines DE, Haissaguerre M, Hindricks G, Iesaka Y, Jackman W, Jalife J, Jais P, Kalman J, Keane D, Kim YH, Kirchhof P, Klein G, Kottkamp H, Kumagai K, Lindsay BD, Mansour M, Marchlinski FE, McCarthy PM, Mont JL, Morady F, Nademanee K, Nakagawa H, Natale A, Nattel S, Packer DL, Pappone C, Prystowsky E, Raviele A, Reddy V, Ruskin JN, Shemin RJ, Tsao HM and Wilber D. 2012 HRS/EHRA/ECAS Expert Consensus Statement on Catheter and Surgical Ablation of Atrial Fibrillation: Recommendations for Patient Selection, Procedural Techniques, Patient Management and Follow-up, Definitions, Endpoints, and Research Trial Design A report of the Heart Rhythm Society (HRS) Task Force on Catheter and Surgical Ablation of Atrial Fibrillation. Developed in partnership with the European Heart Rhythm Association (EHRA), a registered branch of the European Society of Cardiology (ESC) and the European Cardiac Arrhythmia Society (ECAS); and in collaboration with the American College of Cardiology (ACC), American Heart Association (AHA), the Asia Pacific Heart Rhythm Society (APHRS), and the Society of Thoracic Surgeons (STS). Endorsed by the governing bodies of the American College of Cardiology Foundation, the American Heart Association, the European Cardiac Arrhythmia Society, the European Heart Rhythm Association, the Society of Thoracic Surgeons, the Asia Pacific Heart Rhythm Society, and the Heart Rhythm Society. Heart Rhythm. 2012;9:632-+.

5. Benharash P, Buch E, Frank P, Share M, Tung R, Shivkumar K and Mandapati R. Quantitative Analysis of Localized Sources Identified by Focal Impulse and Rotor Modulation Mapping in Atrial Fibrillation. Circ-Arrhythmia Elec. 2015;8:554–561.

6. Sueda T, Nagata H, Shikata H, Orihashi K, Morita S, Sueshiro M, Okada K and Matsuura Y. Simple left atrial procedure for chronic atrial fibrillation associated with mitral valve disease. Ann Thorac Surg. 1996;62:1796–1800.

7. Holm M, Johansson R, Brandt J, Luhrs C and Olsson SB. Epicardial right atrial free wall mapping in chronic atrial fibrillation - Documentation of repetitive activation with a focal spread - A hitherto unrecognised phenomenon in man. Eur Heart J. 1997;18:290–310.

8. Harada A, Konishi T, Fukata M, Higuchi K, Sugimoto T and Sasaki K. Intraoperative map guided operation for atrial fibrillation due to mitral valve disease. Ann Thorac Surg. 2000;69:446–450.

9. Wu TJ, Doshi RN, Huang HLA, Blanche C, Kass RM, Trento A, Cheng W, Karagueuzian HS, Peter CT and Chen PS. Simultaneous biatrial computerized mapping during permanent atrial fibrillation in patients with organic heart disease. J Cardiovasc Electr. 2002;13:571–577.

10. Yamauchi S, Ogasawara H, Saji Y, Bessho R, Miyagi Y and Fujii M. Efficacy of Intraoperative mapping to optimize the surgical ablation of atrial fibrillation in cardiac surgery. Ann Thorac Surg. 2002;74:450–457.

11. Nitta T, Ishii Y, Miyagi Y, Ohmori H, Sakamoto S and Tanaka S. Concurrent multiple left atrial focal activations with fibrillatory conduction and right atrial focal or reentrant activation as the mechanism in atrial fibrillation. J Thorac Cardiov Sur. 2004;127:770–778.

12. Sahadevan J, Ryu K, Peltz L, Khrestian CM, Stewart RW, Markowitz AH and Waldo AL. Epicardial mapping of chronic atrial fibrillation in patients - Preliminary observations. Circulation. 2004;110:3293–3299.

13. Sanders P, Berenfeld O, Hocini MZ, Jais P, Vaidyanathan R, Hsu LF, Garrigue S, Takahashi Y, Rotter M, Sacher F, Scavee C, Ploutz-Snyder R, Jalife J and Haissaguerre M. Spectral analysis identifies sites of high-frequency activity maintaining atrial fibrillation in humans. Circulation. 2005;112:789–797.

14. Cuculich PS, Wang Y, Lindsay BD, Faddis MN, Schuessler RB, Damiano RJ, Li L and Rudy Y. Noninvasive Characterization of Epicardial Activation in Humans With Diverse Atrial Fibrillation Patterns. Circulation. 2010;122:1364-+.

15. de Groot NMS, Houben RPM, Smeets JL, Boersma E, Schotten U, Schalij MJ, Crijns H and Allessie MA. Electropathological Substrate of Longstanding Persistent Atrial Fibrillation in Patients With Structural Heart Disease Epicardial Breakthrough. Circulation. 2010;122:1674–1682.

16. Narayan SM, Krummen DE, Clopton P, Shivkumar K and Miller JM. Direct or Coincidental Elimination of Stable Rotors or Focal Sources May Explain Successful Atrial Fibrillation Ablation On-Treatment Analysis of the CONFIRM Trial (Conventional Ablation for AF With or Without Focal Impulse and Rotor Modulation). J Am Coll Cardiol. 2013;62:138–147.

17. Haissaguerre M, Hocini M, Denis A, Shah AJ, Komatsu Y, Yamashita S, Daly M, Amraoui S, Zellerhoff S, Picat MQ, Quotb A, Jesel L, Lim H, Ploux S, Bordachar P, Attuel G, Meillet V, Ritter P, Derval N, Sacher F, Bernus O, Cochet H, Jais P and Dubois R. Driver Domains in Persistent Atrial Fibrillation. Circulation. 2014;130:530–538.

18. Lee G, Kumar S, Teh A, Madry A, Spence S, Larobina M, Goldblatt J, Brown R, Atkinson V, Moten S, Morton JB, Sanders P, Kistler PM and Kalman JM. Epicardial wave mapping in human long-lasting persistent atrial fibrillation: transient rotational circuits, complex wavefronts, and disorganized activity. Eur Heart J. 2014;35:86–97.

19. Swarup V, Baykaner T, Rostamian A, Daubert JP, Hummel J, Krummen DE, Trikha R, Miller JM, Tomassoni GF and Narayan SM. Stability of Rotors and Focal Sources for Human Atrial Fibrillation: Focal Impulse and Rotor Mapping (FIRM) of AF Sources and Fibrillatory Conduction. J Cardiovasc Electr. 2014;25:1284–1292.

20. Pandit SV and Jalife J. Rotors and the Dynamics of Cardiac Fibrillation. Circ Res. 2013;112:849–862.

21. de Bakker JM. Electrogram recording and analyzing techniques to optimize selection of target sites for ablation of cardiac arrhythmias. Pacing Clin Electrophysiol. 2019;42:1503–1516.

22. Berenfeld O. Quantifying activation frequency in atrial fibrillation to establish underlying mechanisms and ablation guidance. Heart Rhythm. 2007;4:1225–1234.

23. Jarman JW, Wong T, Francis DP, Davies DW, Kanagaratnam P, Markides V and Peters NS. Spatiotemporal Behaviour of High Dominant Frequency During Paroxysmal and Persistent Atrial Fibrillation in the Human Left Atrium. Circulation. 2009;120:S639–S639.

24. Takahashi Y, Sanders P, Jais P, Hocini M, Dubois R, Rotter M, Rostock T, Nalliah CJ, Sacher F, Clementy J and Haissaguerre M. Organization of frequency spectra of atrial fibrillation: Relevance to radiofrequency catheter ablation. J Cardiovasc Electr. 2006;17:382–388.

25. Everett TH, Akar JG, Kok LC, Moorman JR and Haines DE. Use of global atrial fibrillation organization to optimize the success of burst pace termination. J Am Coll Cardiol. 2002;40:1831–1840.

26. Lo LW, Lin YJ, Tsao HM, Chang SL, Hu YF, Tsai WC, Tuan DC, Chang CJ, Lee PC, Tai CT, Tang WH, Suenari K, Huang SY, Higa S and Chen SA. Characteristics of Complex Fractionated Electrograms in Nonpulmonary Vein Ectopy Initiating Atrial Fibrillation/Atrial Tachycardia. J Cardiovasc Electr. 2009;20:1305–1312.

27. Ng J, Borodyanskiy AI, Chang ET, Villuendas R, Dibs S, Kadish AH and Goldberger JJ. Measuring the Complexity of Atrial Fibrillation Electrograms. J Cardiovasc Electr. 2010;21:649–655.

28. Valderrabano M, Chen PS and Lin SF. Spatial distribution of phase singularities in ventricular fibrillation. Circulation. 2003;108:354–359.

29. Gray RA, Pertsov AM and Jalife J. Spatial and temporal organization during cardiac fibrillation. Nature. 1998;392:75–8.

30. Aronis KN, Berger RD and Ashikaga H. Rotors: How Do We Know When They Are Real? Circ Arrhythm Electrophysiol. 2017;10.

31. Kuklik P, Zeemering S, Maesen B, Maessen J, Crijns HJ, Verheule S, Ganesan AN and Schotten U. Reconstruction of instantaneous phase of unipolar atrial contact electrogram using a concept of sinusoidal recomposition and Hilbert transform. IEEE Trans Biomed Eng. 2015;62:296–302.

32. Kennedy DJ, Vetteth S, Periyasamy SM, Kanj M, Fedorova L, Khouri S, Kahaleh MB, Xie Z, Malhotra D, Kolodkin NI, Lakatta EG, Fedorova OV, Bagrov AY and Shapiro JI. Central role for the cardiotonic steroid marinobufagenin in the pathogenesis of experimental uremic cardiomyopathy. Hypertension. 2006;47:488–95.

33. de Groot NM, Houben RP, Smeets JL, Boersma E, Schotten U, Schalij MJ, Crijns H and Allessie MA. Electropathological substrate of longstanding persistent atrial fibrillation in patients with structural heart disease: epicardial breakthrough. Circulation. 2010;122:1674–82.

34. Mansour M, Ruskin J and Keane D. Initiation of atrial fibrillation by ectopic beats originating from the ostium of the inferior vena cava. J Cardiovasc Electr. 2002;13:1292–1295.

35. Lin WS, Tai CT, Hsieh MH, Tsai CF, Lin YK, Tsao HM, Huang JL, Yu WC, Yang SP, Ding YA, Chang MS and Chen SA. Catheter ablation of paroxysmal atrial fibrillation initiated by non-pulmonary vein ectopy. Circulation. 2003;107:3176–3183.

36. Shah D, Haissaguerre M, Jais P and Hocini M. Nonpulmonary vein foci: Do they exist? Pace. 2003;26:1631–1635.

37. Lee SH, Tai CT, Hsieh MH, Tsao HM, Lin YJ, Chang SL, Huang JL, Lee KT, Chen YJ, Cheng JJ and Chen SA. Predictors of non-pulmonary vein ectopic beats initiating paroxysmal atrial fibrillation - Implication for catheter ablation. J Am Coll Cardiol. 2005;46:1054–1059.

38. Yamada T, Murakami Y, Okada T and Murohara T. Focal atrial fibrillation associated with multiple breakout sites at the crista terminalis. Pace. 2006;29:207–210.

39. Pastor A, Nunez A, Magalhaes A, Awamleh P and Garcia-Cosio F. The superior vena cava as a site of ectopic foci in atrial fibrillation. Rev Esp Cardiol. 2007;60:68–71.

40. Hwang C and Chen PS. Ligament of Marshall: Why it is important for atrial fibrillation ablation. Heart Rhythm. 2009;6:S35–S40.

41. Yamaguchi T, Tsuchiya T, Miyamoto K, Nagamoto Y and Takahashi N. Characterization of non-pulmonary vein foci with an EnSite array in patients with paroxysmal atrial fibrillation. Europace. 2010;12:1698–1706.

42. Elayi CS, Di Biase L, Bai R, Burkhardt JD, Mohanty P, Santangeli P, Sanchez J, Hongo R, Gallinghouse GJ, Horton R, Bailey S, Beheiry S and Natale A. Administration of Isoproterenol and Adenosine to Guide Supplemental Ablation After Pulmonary Vein Antrum Isolation. J Cardiovasc Electr. 2013;24:1199–1206.

43. Haissaguerre M, Hocini M, Denis A, Shah AJ, Komatsu Y, Yamashita S, Daly M, Amraoui S, Zellerhoff S, Picat MQ, Quotb A, Jesel L, Lim H, Ploux S, Bordachar P, Attuel G, Meillet V, Ritter P, Derval N, Sacher F, Bernus O, Cochet H, Jais P and Dubois R. Driver domains in persistent atrial fibrillation. Circulation. 2014;130:530–8.

44. Di Biase L, Burkhardt JD, Mohanty P, Sanchez J, Mohanty S, Horton R, Gallinghouse GJ, Bailey SM, Zagrodzky JD, Santangeli P, Hao S, Hongo R, Beheiry S, Themistoclakis S, Bonso A, Rossillo A, Corrado A, Raviele A, Al-Ahmad A, Wang P, Cummings JE, Schweikert RA, Pelargonio G, Dello Russo A, Casella M, Santarelli P, Lewis WR and Natale A. Left atrial appendage: an underrecognized trigger site of atrial fibrillation. Circulation. 2010;122:109–18.

45. Di Biase L, Burkhardt JD, Mohanty P, Mohanty S, Sanchez JE, Trivedi C, Gunes M, Gokoglan Y, Gianni C, Horton RP, Themistoclakis S, Gallinghouse GJ, Bailey S, Zagrodzky JD, Hongo RH, Beheiry S, Santangeli P, Casella M, Dello Russo A, Al-Ahmad A, Hranitzky P, Lakkireddy D, Tondo C and Natale A. Left Atrial Appendage Isolation in Patients With Longstanding Persistent AF Undergoing Catheter Ablation: BELIEF Trial. J Am Coll Cardiol. 2016;68:1929–1940.

46. Lewis T, Drury AN and Iliescu CC. Further observations upon the state of rapid re-excitation of the auricles. Heart-J Stud Circ. 1921;8:311–339.

47. Allessie MA, Bonke FIM and Schopman FJ. Circus Movement in Rabbit Atrial Muscle as a Mechanism of Tachycardia. Circ Res. 1973;33:54–62.

48. Spach MS, Miller WT, Dolber PC, Kootsey JM, Sommer JR and Mosher CE. The Functional-Role of Structural Complexities in the Propagation of Depolarization in the Atrium of the Dog - Cardiac Conduction Disturbances Due to Discontinuities of Effective Axial Resistivity. Circ Res. 1982;50:175–191.

49. Hocini M, Ho SY, Kawara T, Linnenbank AC, Potse M, Shah D, Jais P, Janse MJ, Haissaguerre M and de Bakker JMT. Electrical conduction in canine pulmonary veins - Electrophysiological and anatomic correlation. Circulation. 2002;105:2442–2448.

50. Hansen BJ, Zhao J, Csepe TA, Moore BT, Li N, Jayne LA, Kalyanasundaram A, Lim P, Bratasz A, Powell KA, Simonetti OP, Higgins RS, Kilic A, Mohler PJ, Janssen PM, Weiss R, Hummel JD and Fedorov VV. Atrial fibrillation driven by micro-anatomic intramural re-entry revealed by simultaneous sub-epicardial and sub-endocardial optical mapping in explanted human hearts. Eur Heart J. 2015;36:2390–401.

