## Supplementary material for "Atrial Fibrillation Drivers: Redefining the Electrophysiological Substrate": Data Supplemental

**SUPPLEMENTAL MATERIAL**

Data Supplement consists of the following:

- Figures S1 to S13:

  Rotational Activities in Hearts #1- #13

- Figures S14 to S21:

  Rotational Activities Trajectories in Hearts

- Figure S22:

  Anchored rotational activity at borderzone between low and normal voltage .

- Figure S23:

  Slow conduction during reentrant and non- reentrant rhythm.

- Figure S24 to S S34:

  Reentrant trajectory and activation delay.

- Figure S35:

  Activation delay along reentrant trajectory in 8 following reentrant loops.

- Figure S36:

  Bipolar voltage maps during rotational activity over time.

- Figure S37:

  Activation delay along reentrant trajectory in all regions.

- Figure S38:

  Activation delay along reentrant trajectory in all regions.

Percentage of rotational activity compared to mapping time.

- Figure S45-S50:

  Fiber angle and fiber crossing detection in ROI's in all 6 bi-atrial regions.

- Figure S51:

  Activation delay in AF, and sinus rhythm/ paced rhythm showing similar located slow conduction zones.

- Figure S52:

  Correlation of electrogram measures OI, FI, ShEn, CL, DF with stability of rotational activity.

- Figure S53:

  Slow conduction zones similar located pre and post PVI.

Supplemental Figure

#### Supplemental Figures

Supplemental Figure S1

### Heart #1

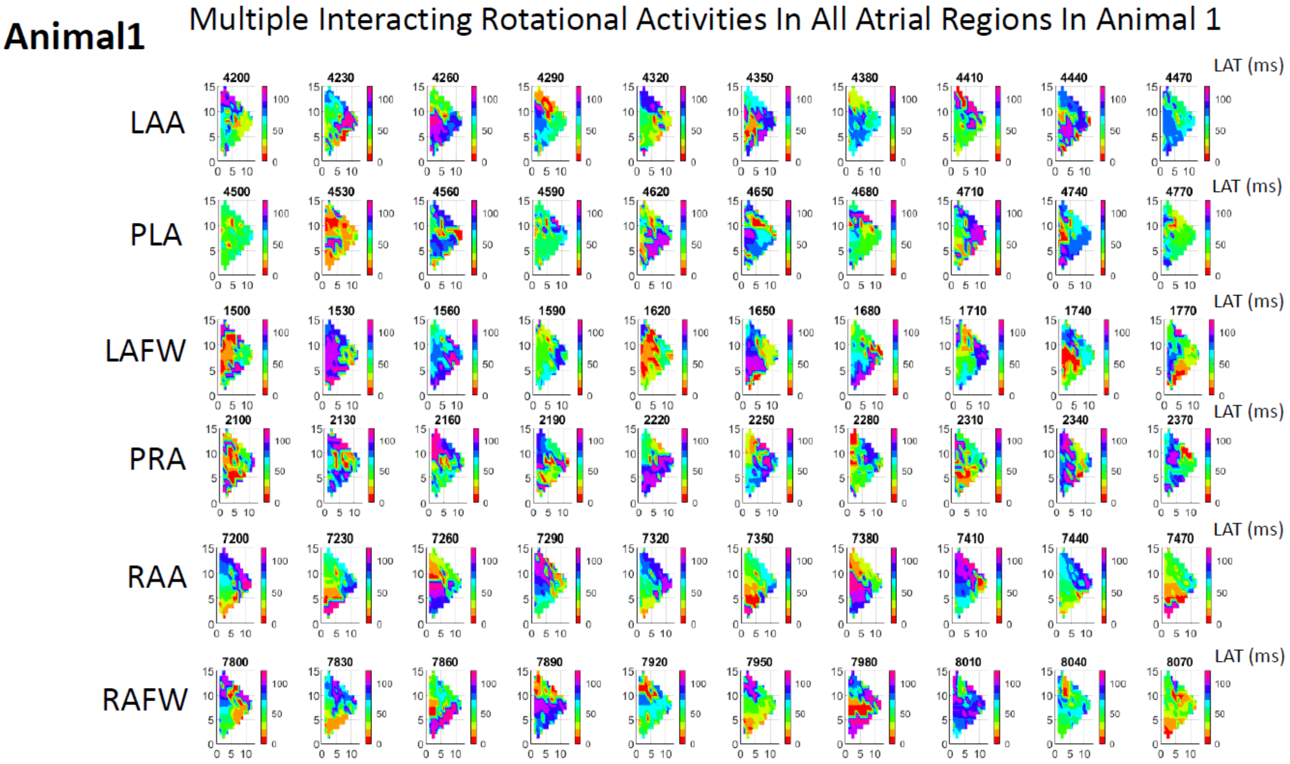


Heart #1: Multiple interacting rotational activities in all regions.

### Heart #2

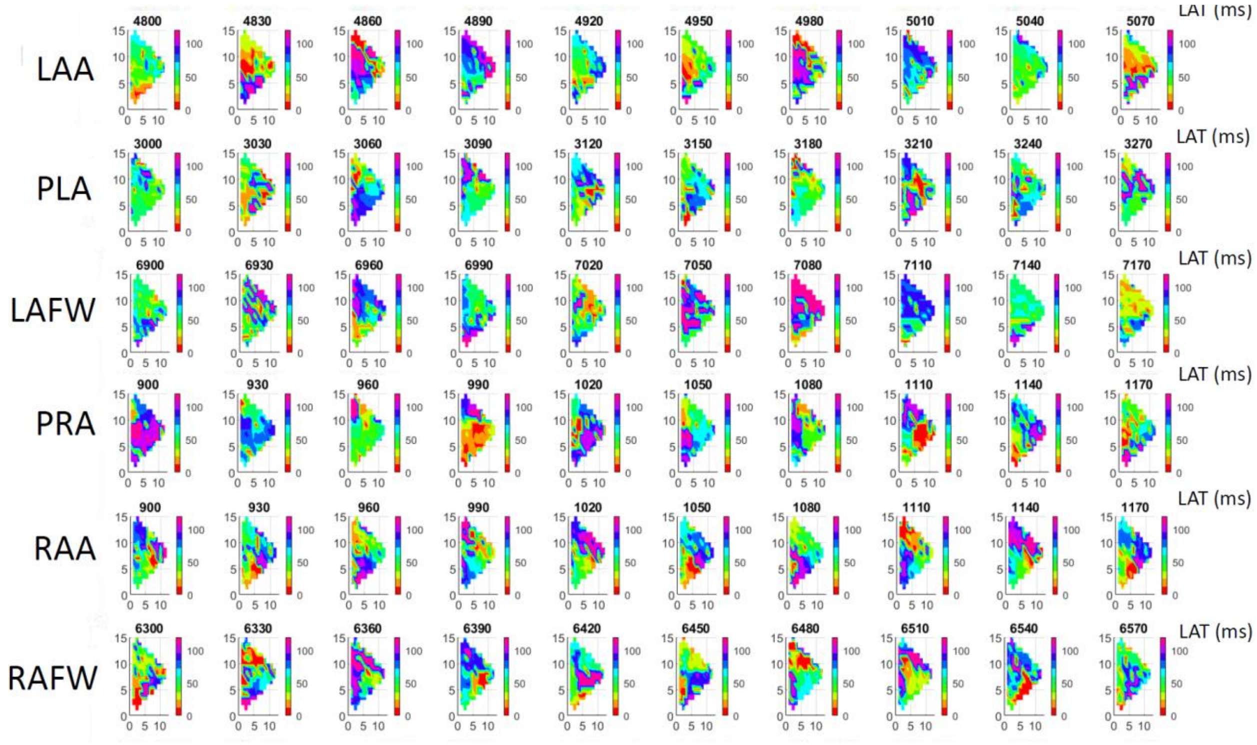


Heart #2: Multiple interacting rotational activities in all regions.

Supplemental Figure S4

### Heart #3

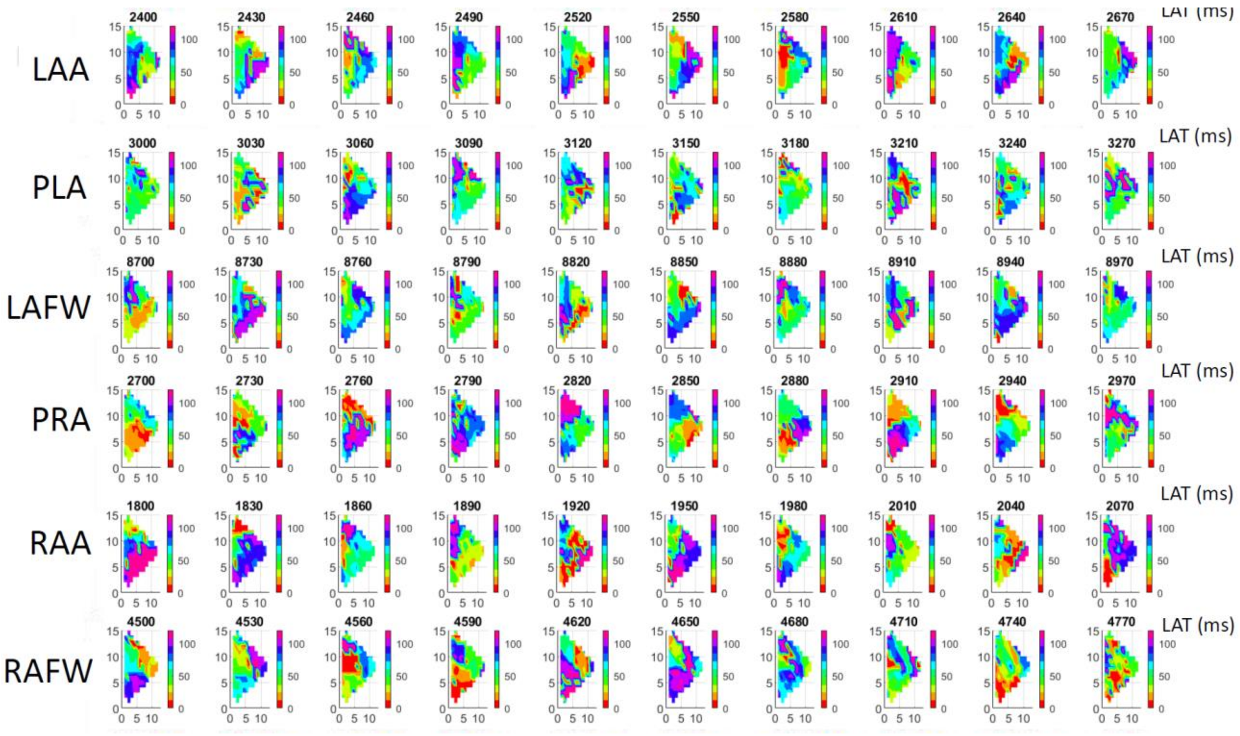


Heart #3: Multiple interacting rotational activities in all regions.

Supplemental Figure S5

### Heart #4

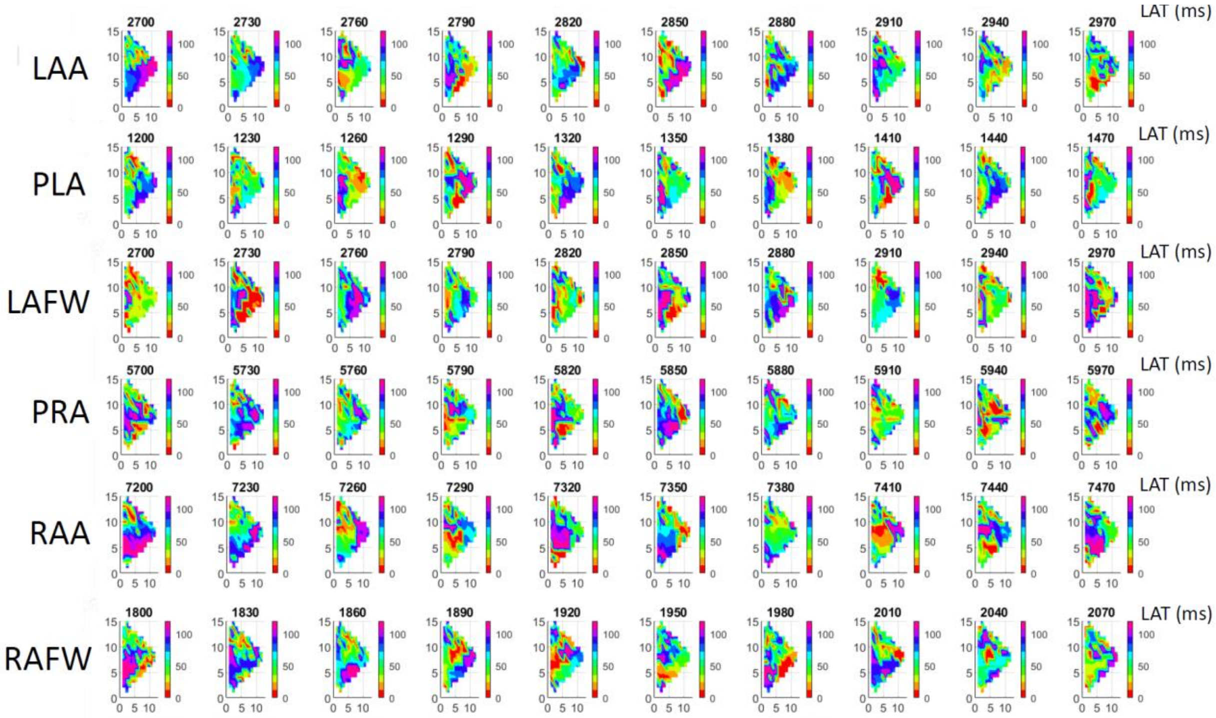


Heart #4: Multiple interacting rotational activities in all regions.

Supplemental Figure S6

### Heart #5

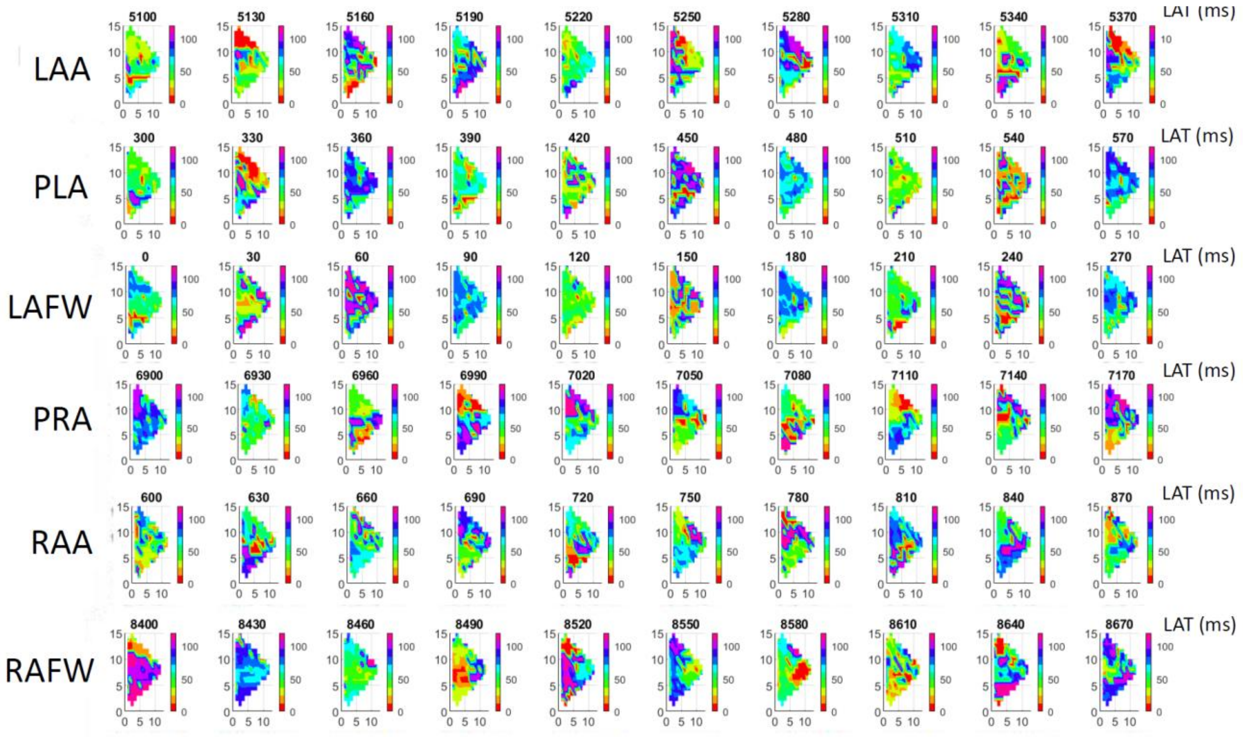


Heart #5: Multiple interacting rotational activities in all regions.

Supplemental Figure S7

### Heart #6

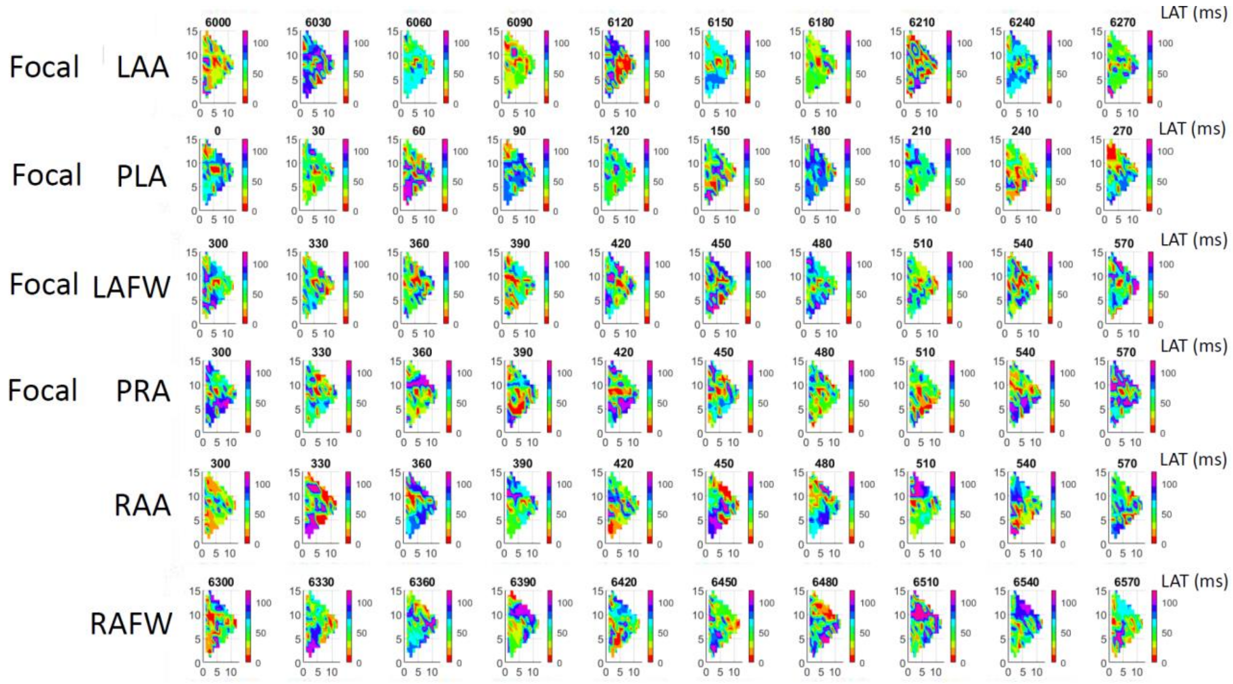


Heart #6: Multiple interacting rotational and focal activities in all regions.

Supplemental Figure S8

### Heart #7

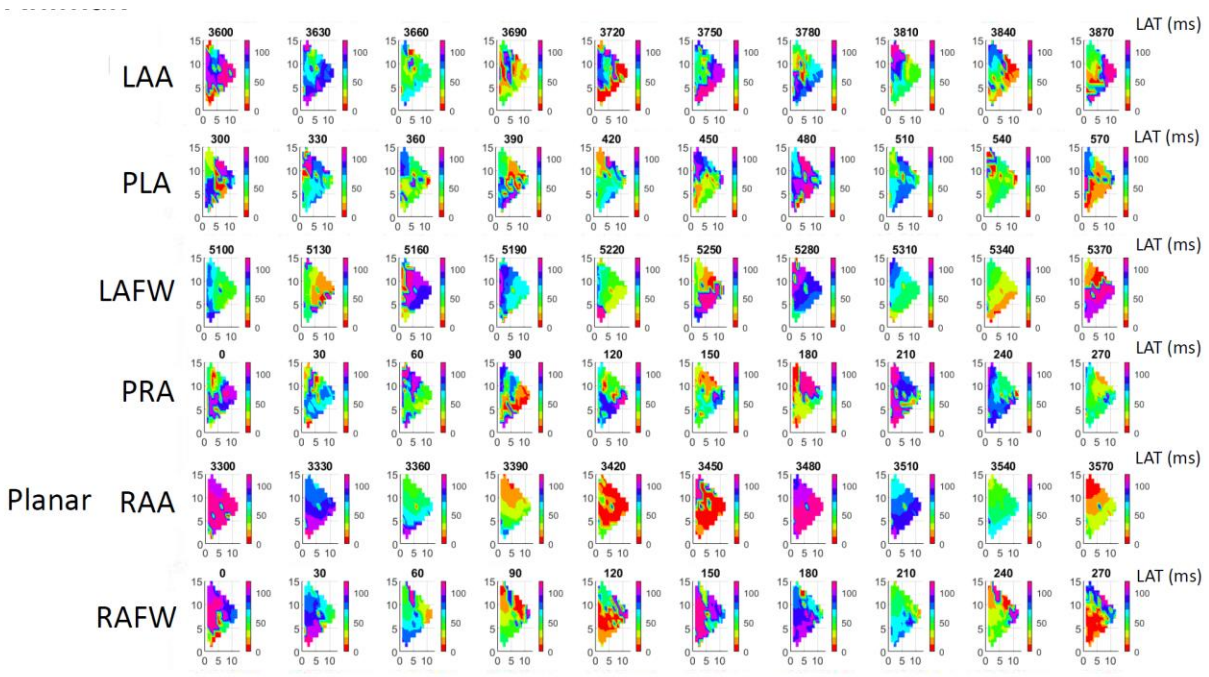


Heart #7: Multiple interacting rotational and focal activities in all regions.

Supplemental Figure S9

### Heart #8

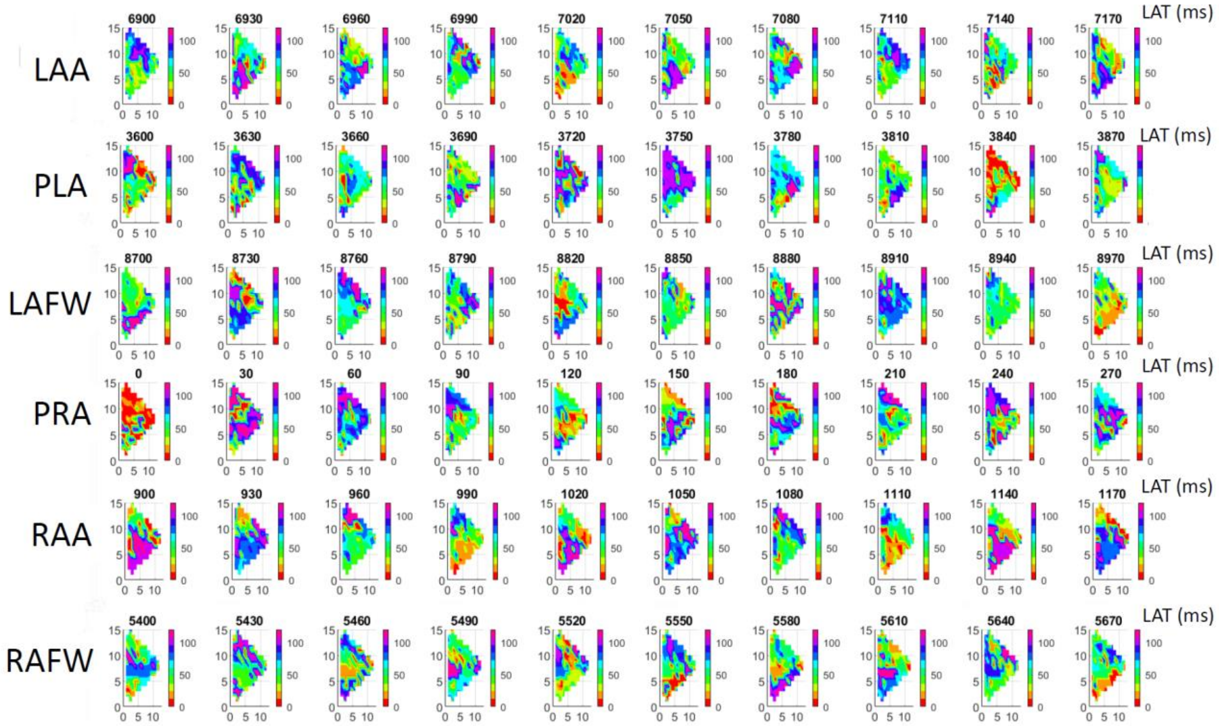


Heart #8: Multiple interacting rotational activities in all regions.

Supplemental Figure S10

### Heart #9

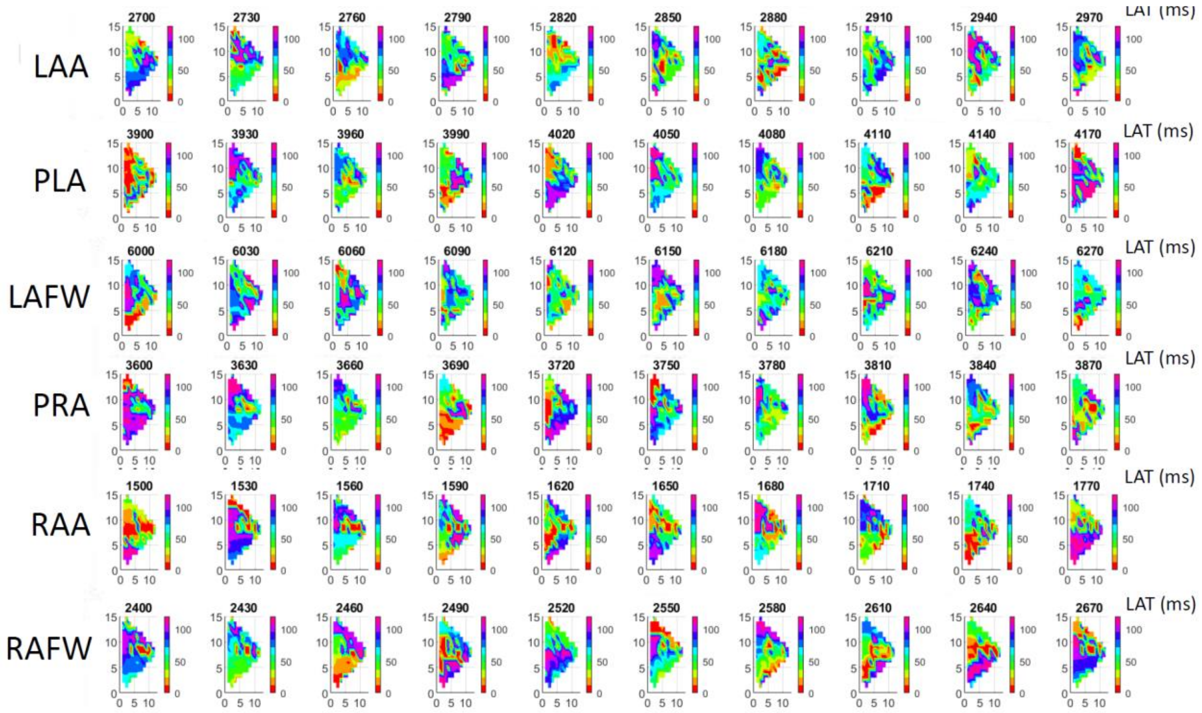


Heart #9: Multiple interacting rotational activities in all regions.

Supplemental Figure S11

### Heart #10

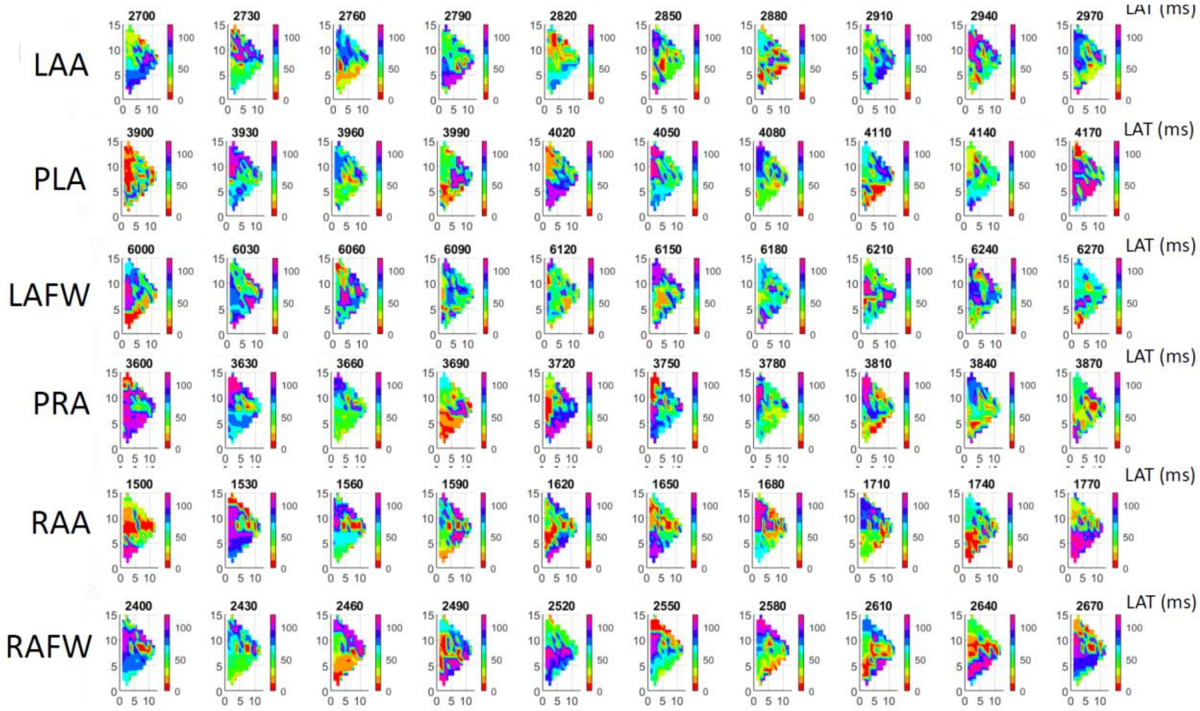


Heart #10: Multiple interacting rotational activities in all regions.

Supplemental Figure S12

### Heart #11

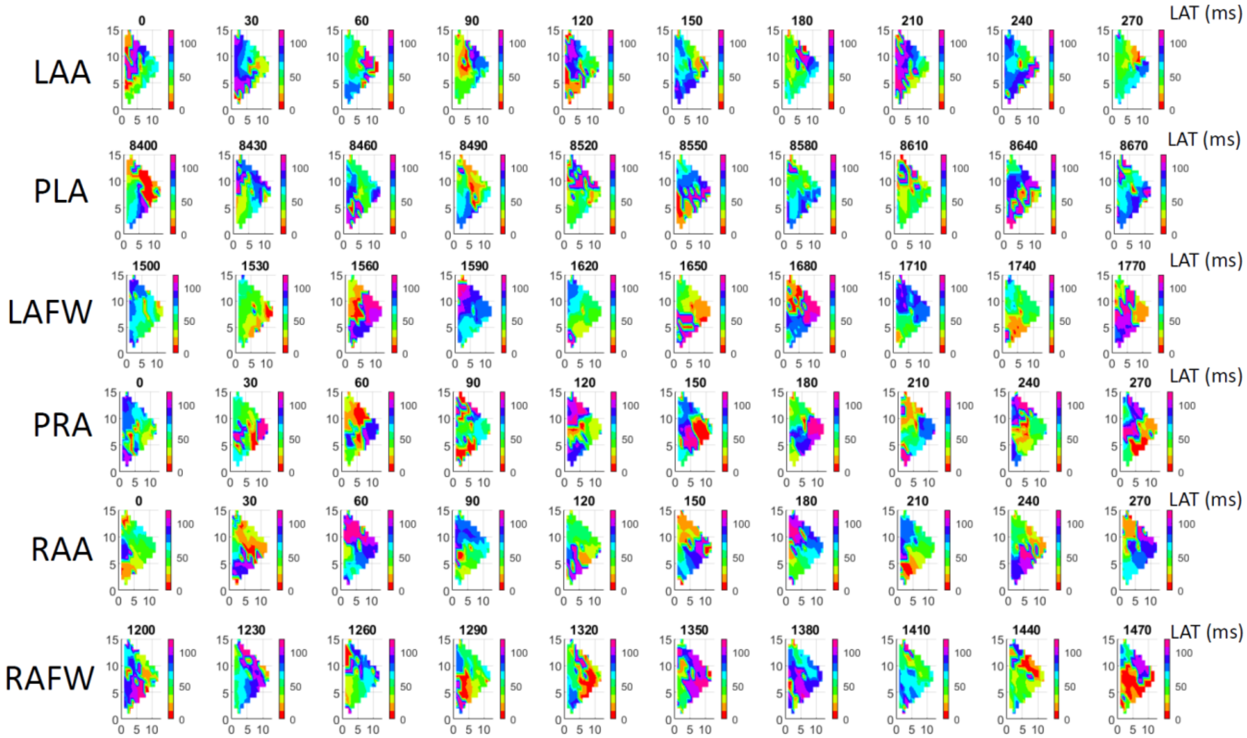


Heart #11: Multiple interacting rotational activities in all regions.

Supplemental Figure S13

### Heart #12

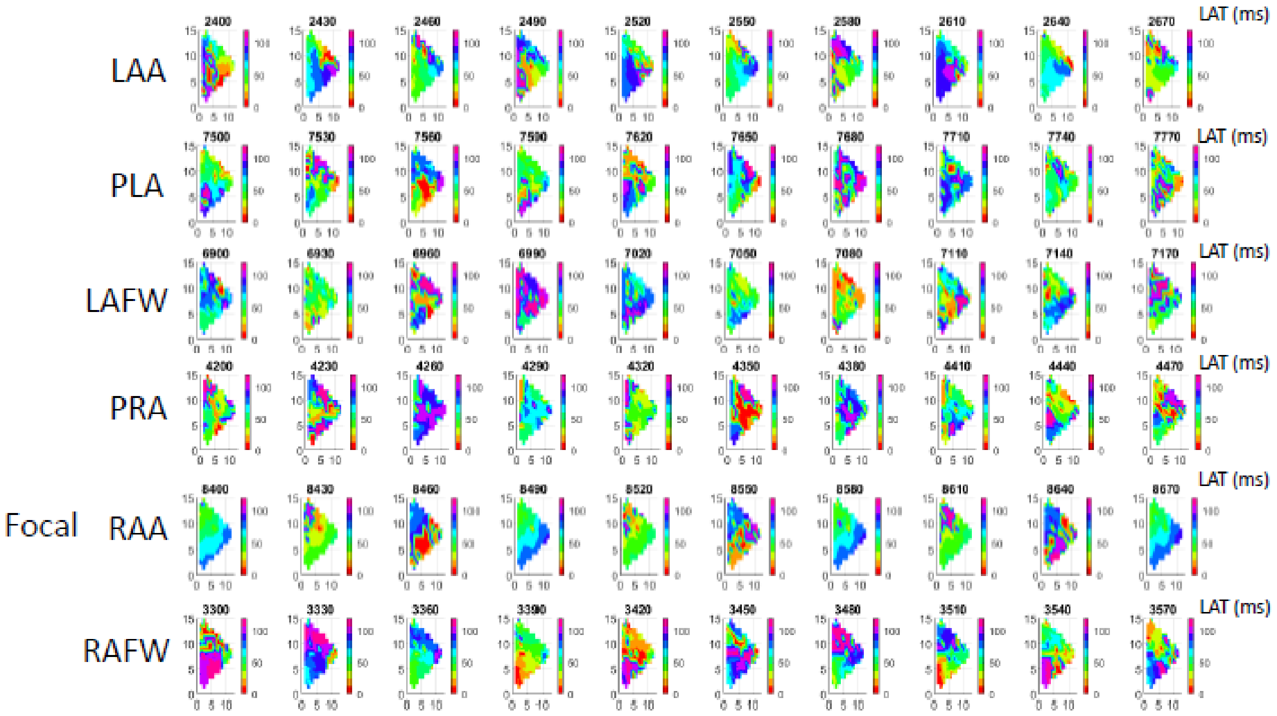


Heart #12: Multiple interacting rotational activities in all regions.

Supplemental Figure S14

### Heart #13

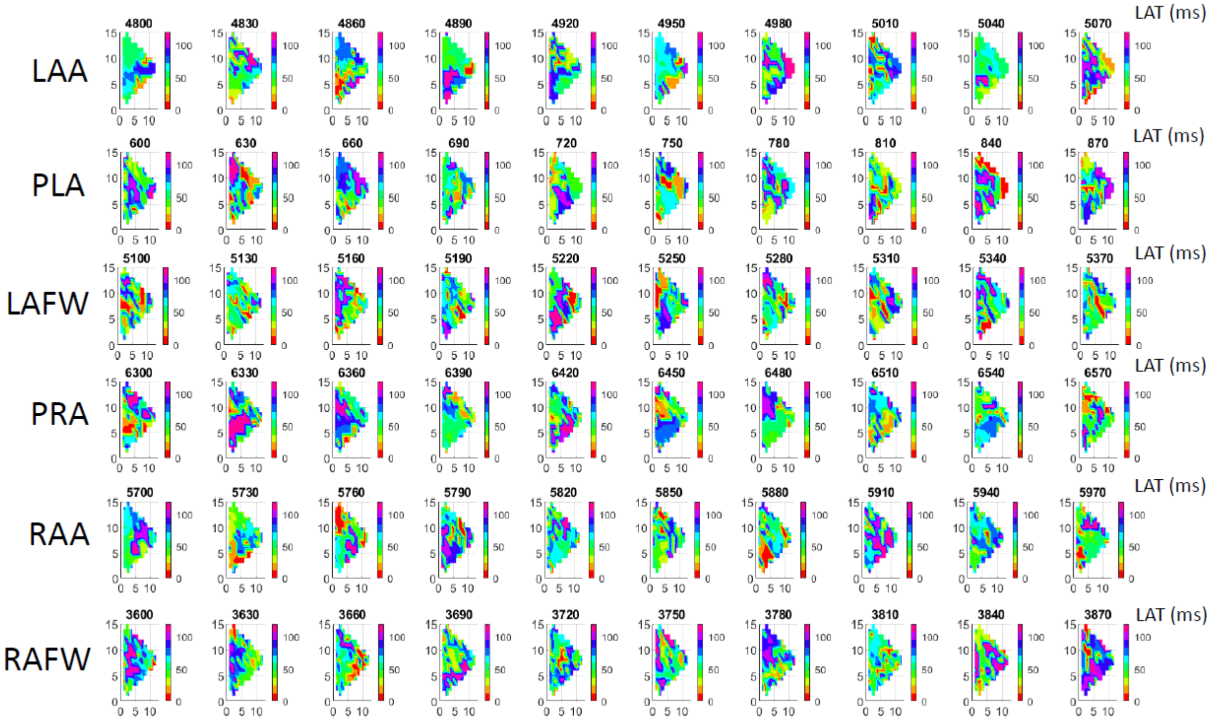


Heart #13: Multiple interacting rotational activities in all regions.

Supplemental Figure S15

### Heart #1

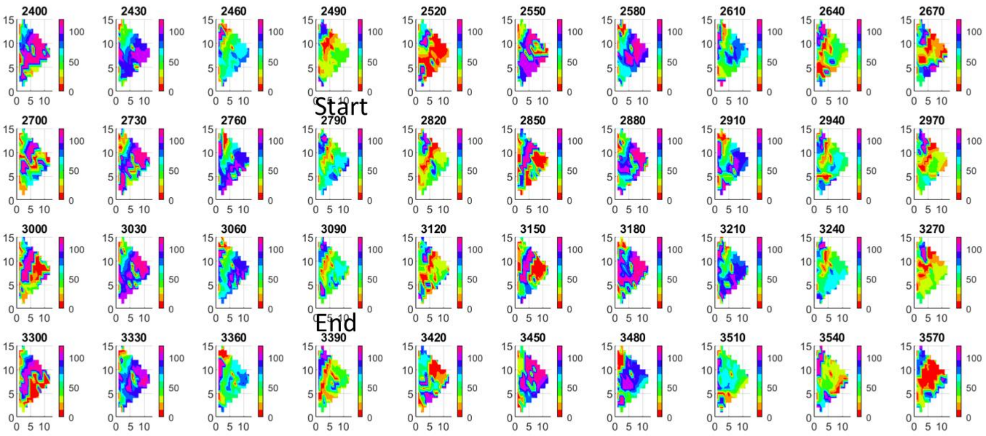


Heart #1: Figure Eight Trajectory in Heart #1 in the PRA

Supplemental Figure S16

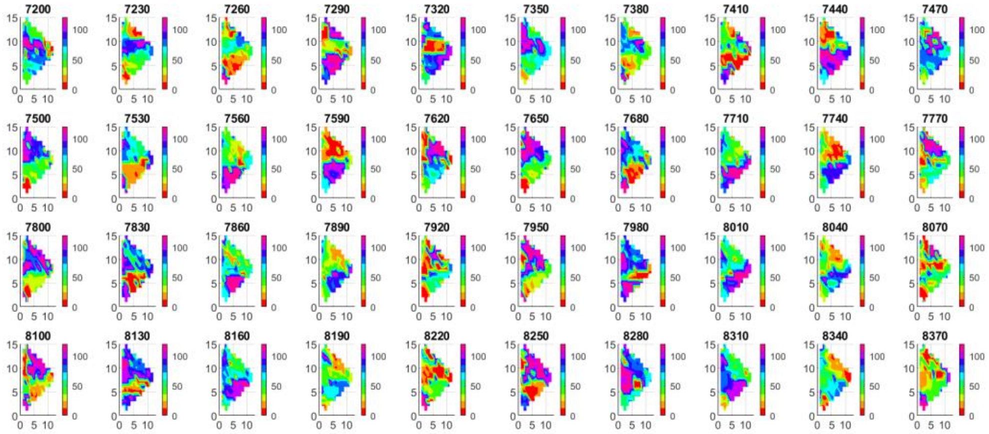


Heart #1: Figure of Eight Trajectory in Heart #1 in the RAFW

Supplemental Figure S17

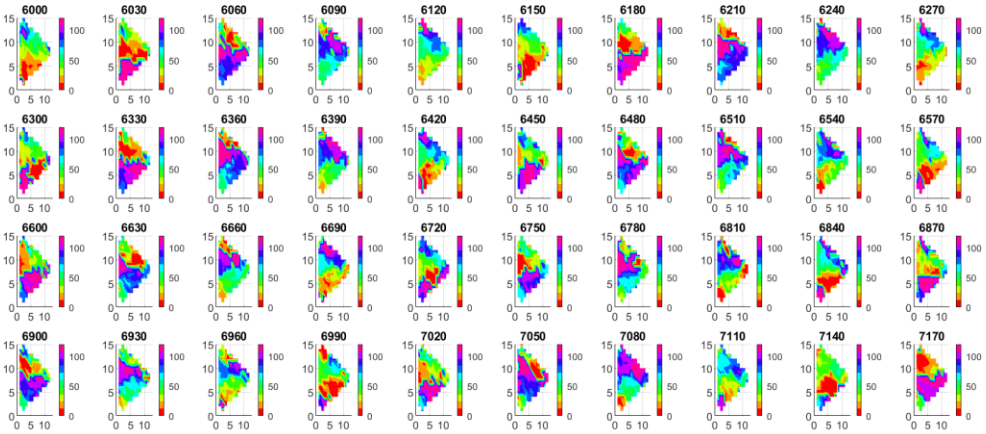

Heart #1: Figure of Eight Trajectory in Heart #1 in the RAFW: Reentrant loop larger than mapping field

Supplemental Figure S18

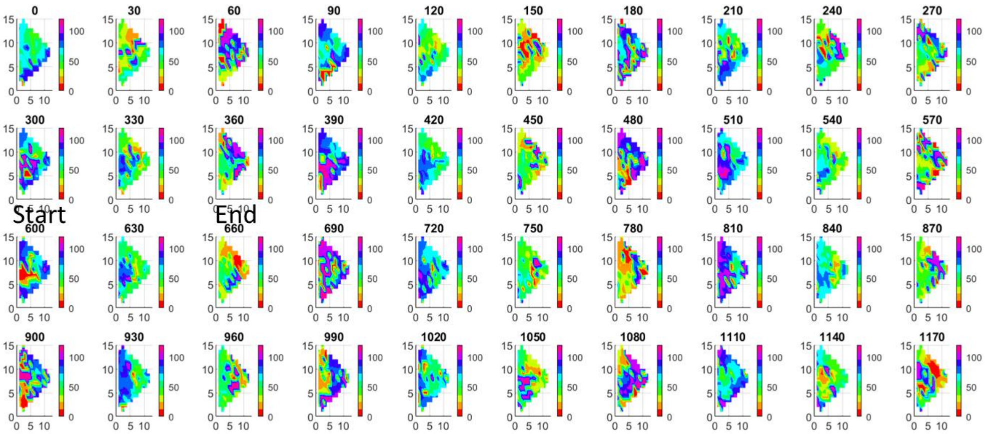


Heart #2: Small Reentrant Loop in Heart #2 in the LAA

Supplemental Figure S19

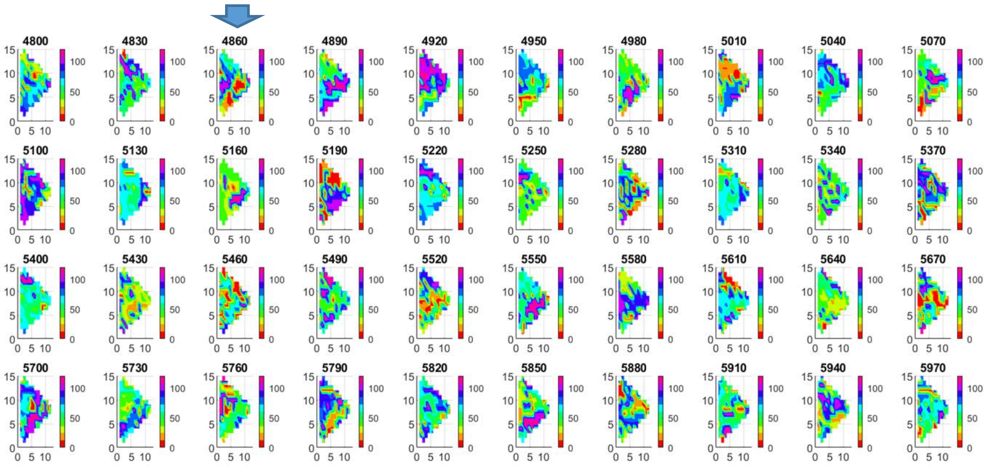


Heart #2: Block Line in Heart #2 in the MIDPLA

Supplemental Figure S20

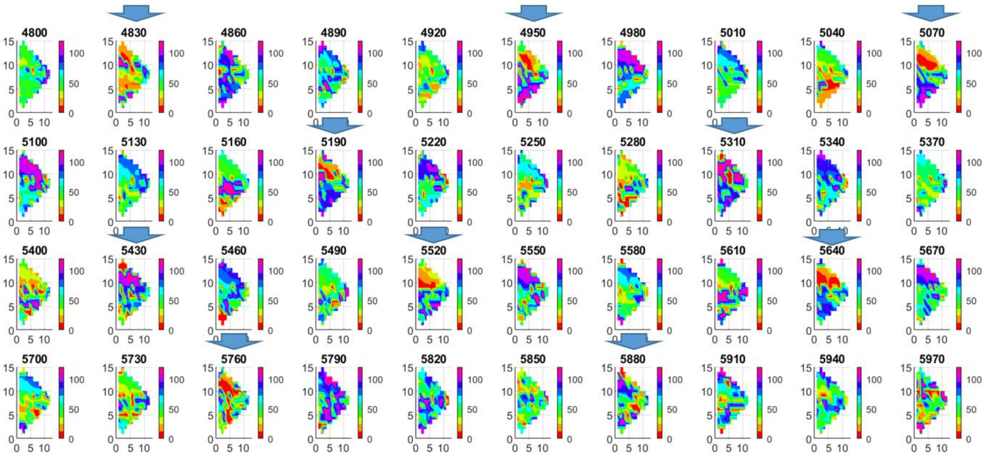


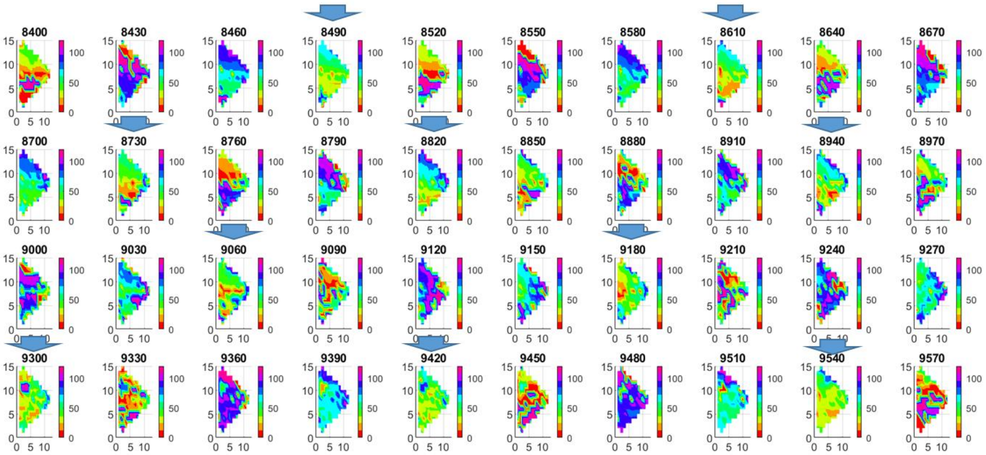


Heart #5: Repetitive Focal Activities in Heart #5 in the PLA

Supplemental Figure S21

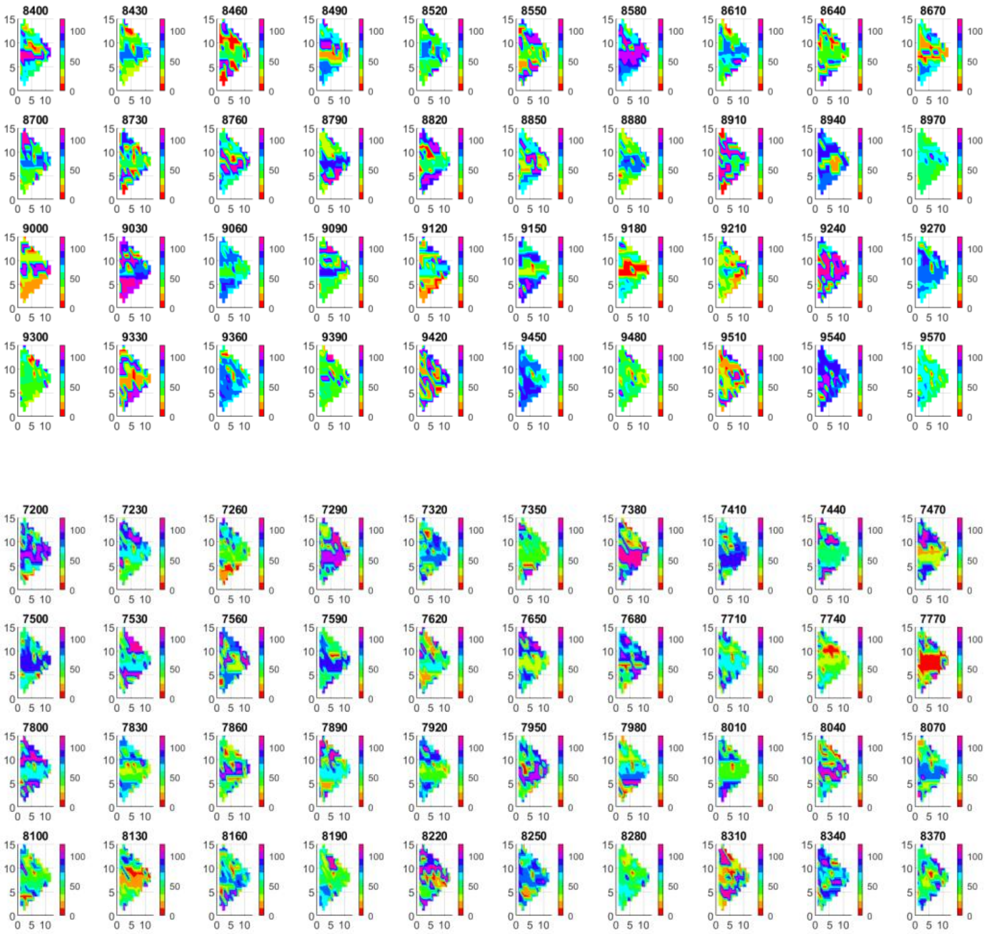


Heart #5: Instantaneous activation along thin lines in Heart #5 in the LAFW

Supplemental Figure S22

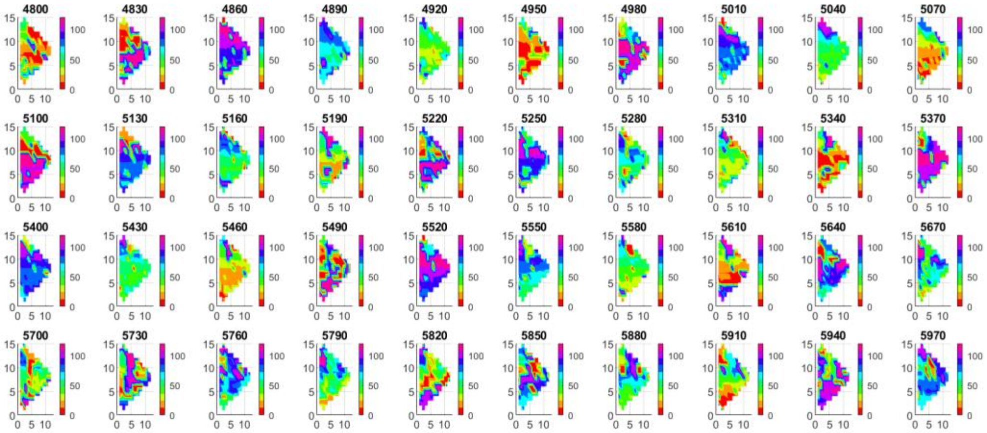

Heart #5: Figure of eight activation along thin activation lines (5700ms – 5850ms) in Heart #5 in the PRA

Supplemental Figure S23

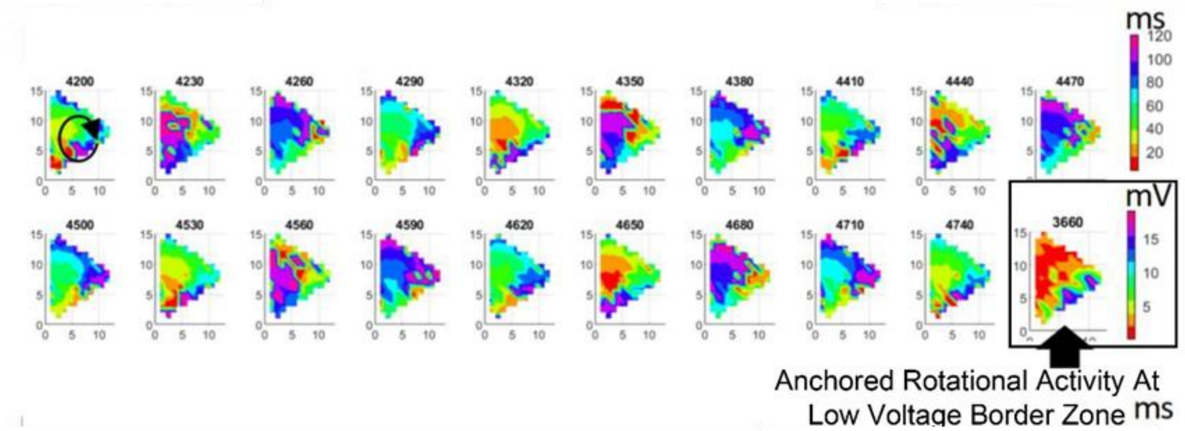


Heart #1: Rotational activity anchored at low voltage border zone.

Supplemental Figure S24

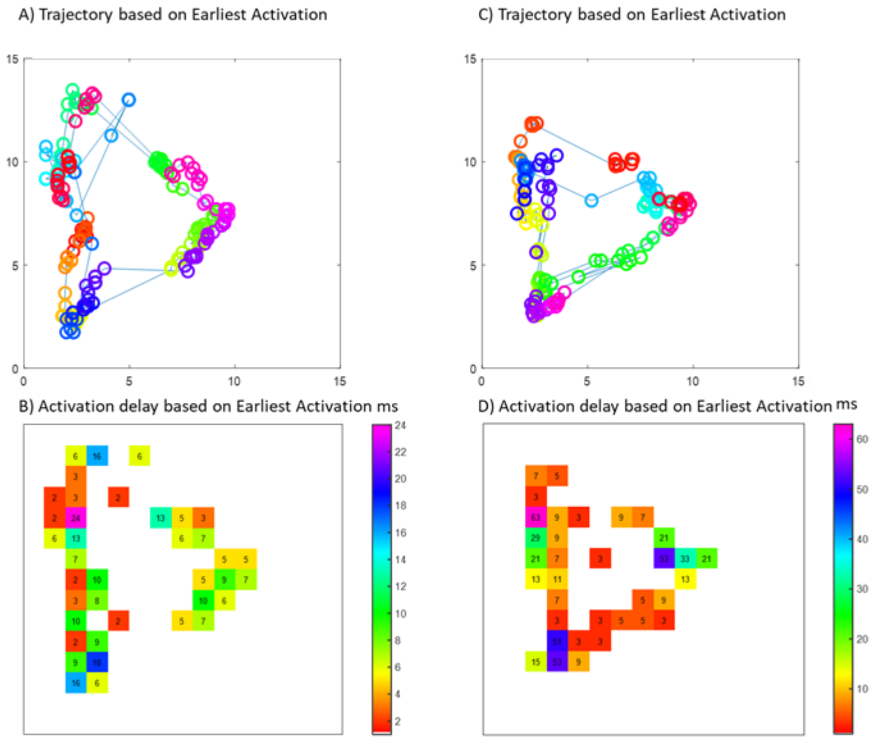


Heart #10: Reentrant trajectory and activation delay in 2 loops

Supplemental Figure S25

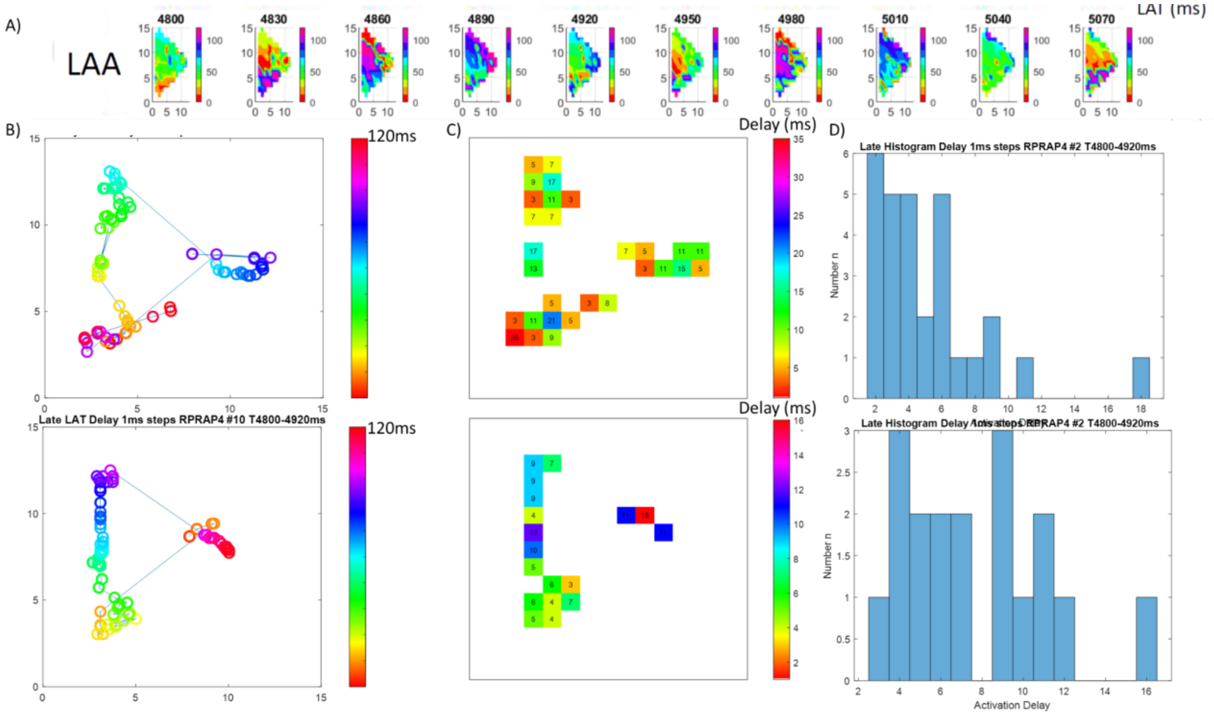


Heart #2, Region LAA: A) Activation time map. B) Reentrant trajectory and activation delay in ROIs (2.5 mm x 2.5 mm) in 2 loops. C) Histogram analysis of activation delay.

Supplemental Figure S26

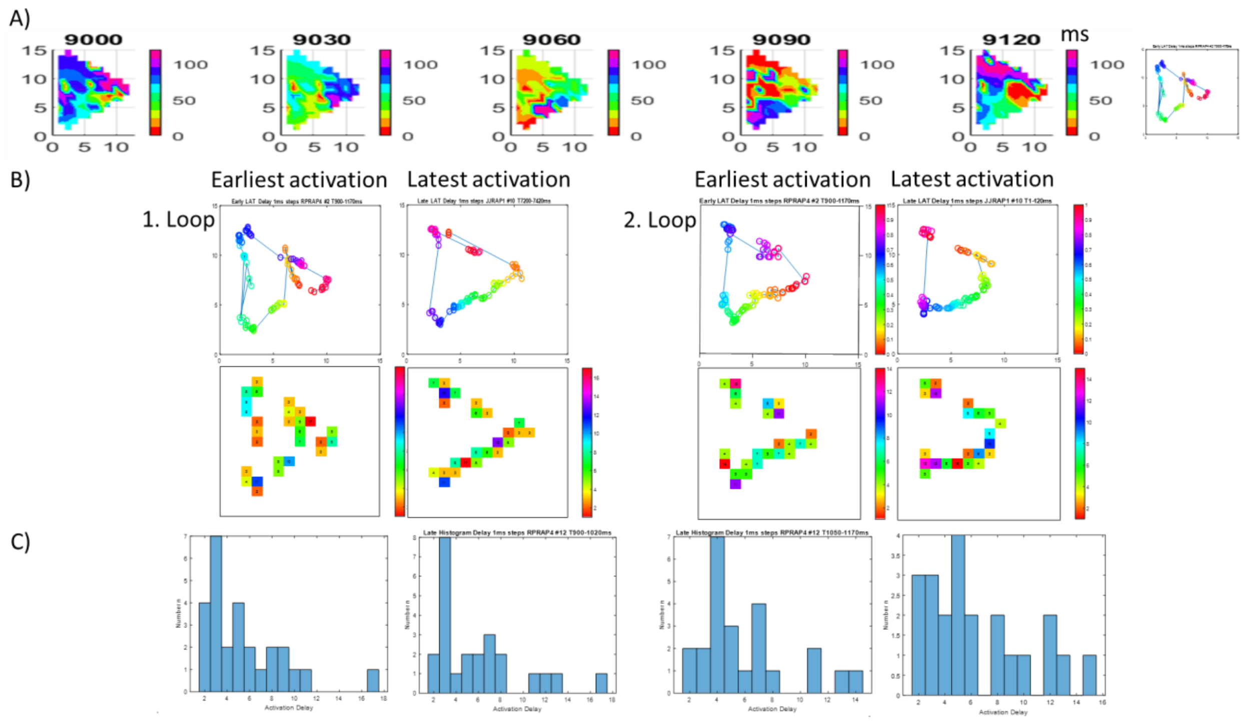


Heart #2, Region RAA: A) Activation time map. B) Reentrant trajectory and activation delay in ROIs (2.5 mm x 2.5 mm) in 2 loops. C) Histogram analysis of activation delay.

### Supplemental Figure S27

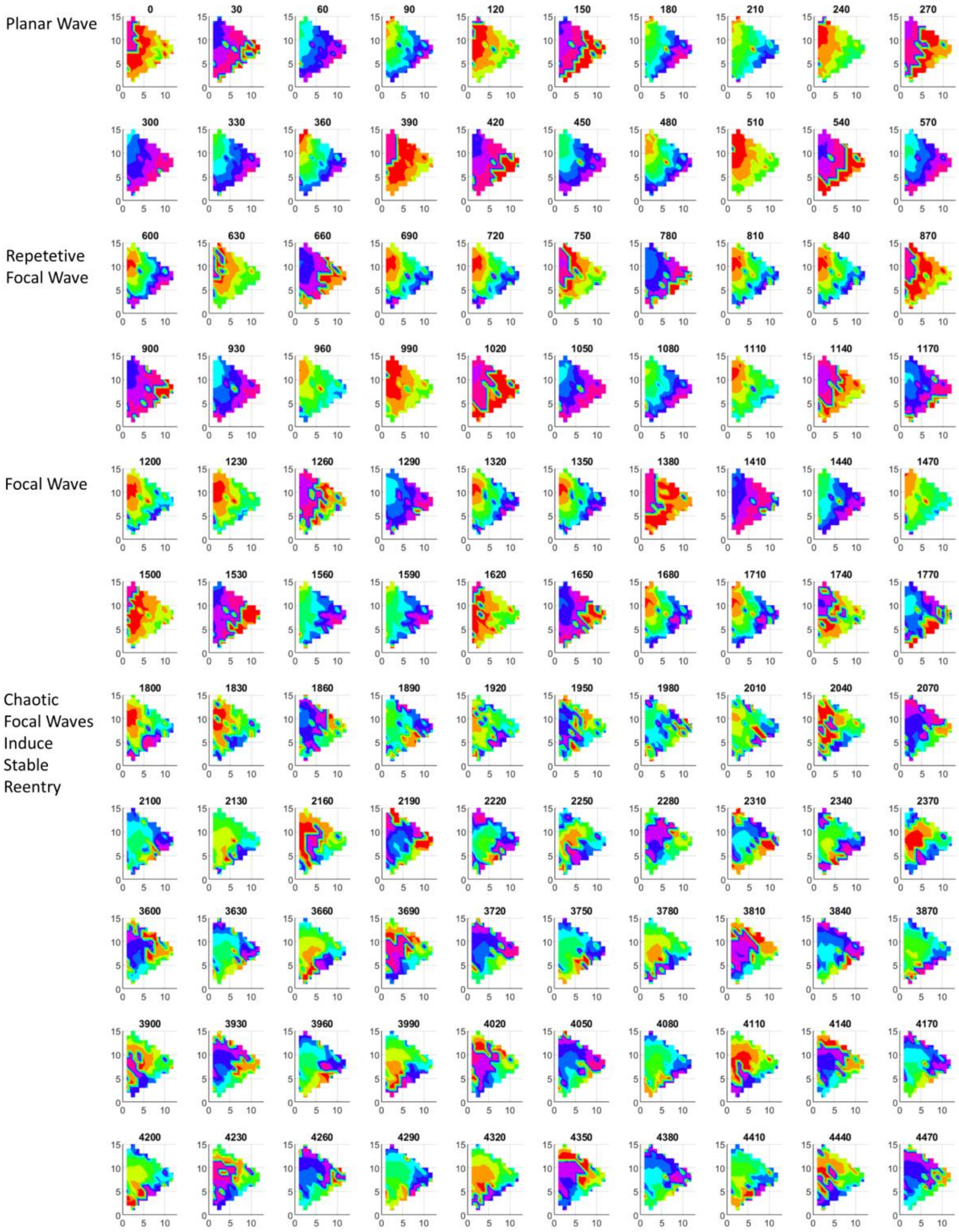

Supplemental Figure S28

Heart #7, Region RAA: A) Development of Rotational Activity beginning with repetitive Planar Wave, Focal Wave Activation. Then 1 first instable reentrant loop which results in multiple chaotic wavelets and then in stable reentrant.

Supplemental Figure S29

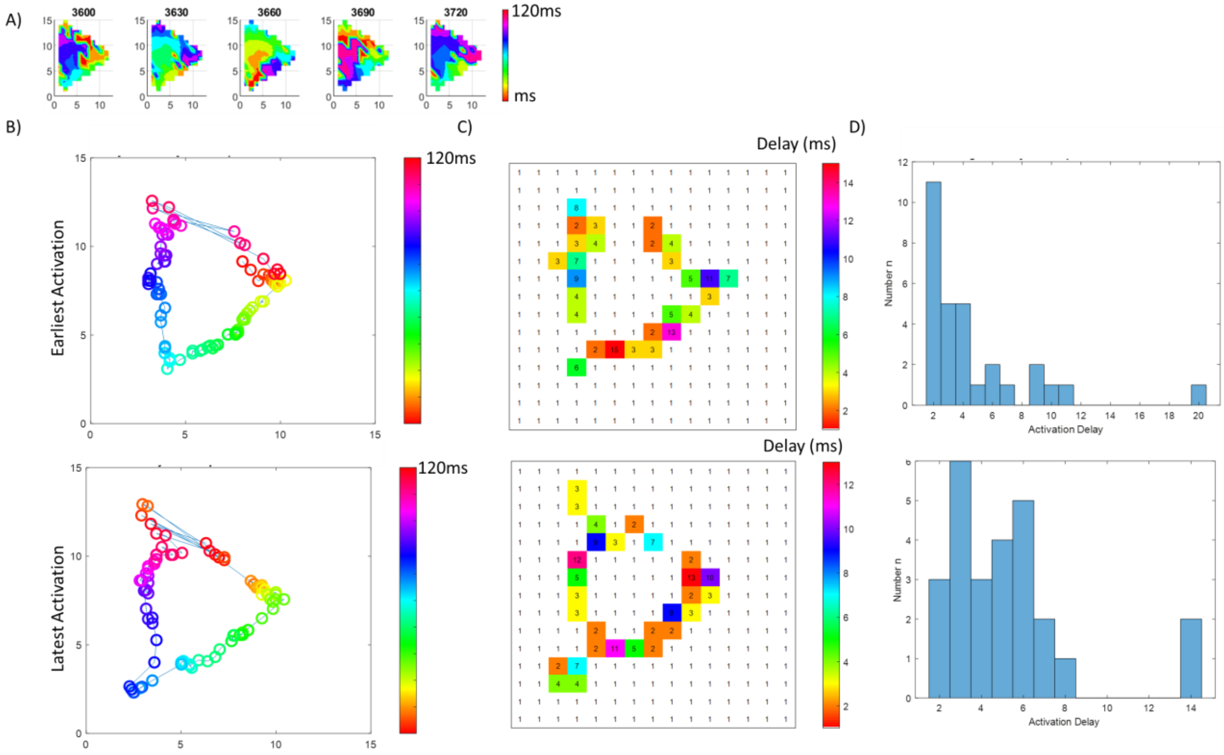


Heart #7, Region RAA: A) Stable Rotational Activity in LAT map. B) Trajectory of first loop based on earliest activation (upper figure panel), latest activation (lower figure panel) within series of activation time maps in 1 msec steps. C) Activation delay along reentrant trajectory. D) Histogram of activation delay.

Supplemental Figure S30

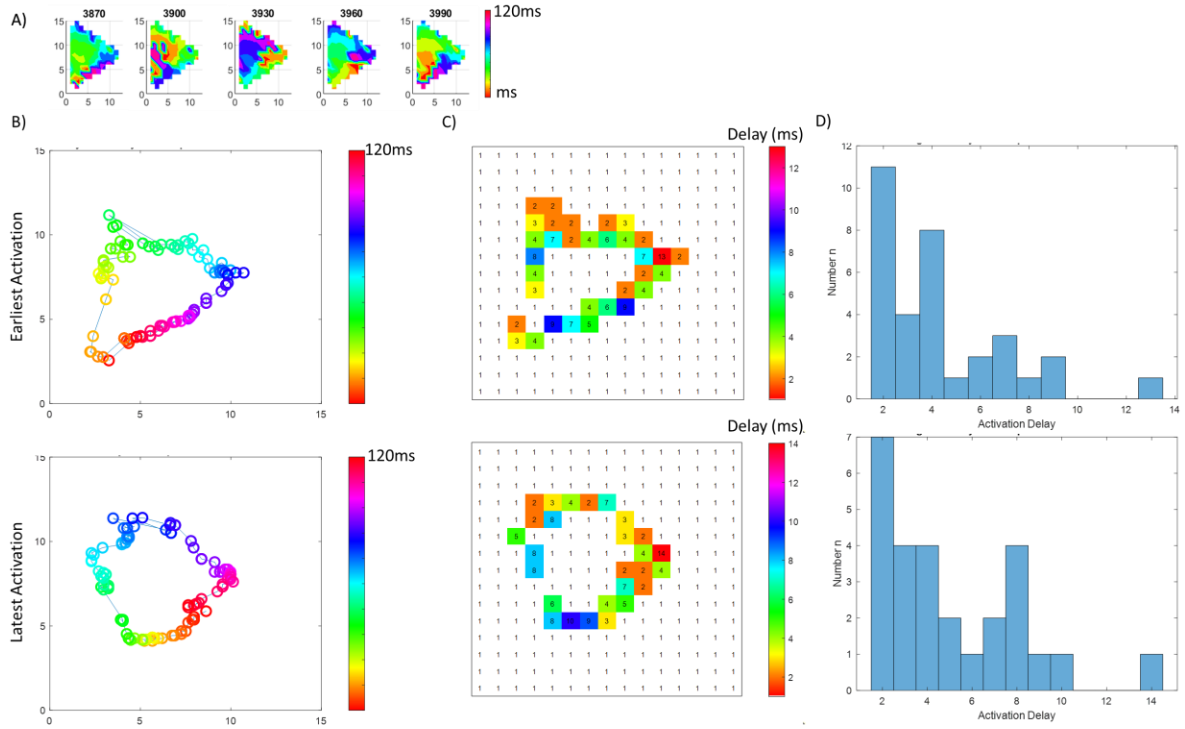


Heart #7, Region RAA: A) Stable Rotational Activity in LAT map. B) Trajectory of loop based on earliest activation (upper figure panel), latest activation (lower figure panel) within series of activation time maps in 1 msec steps. C) Activation delay along reentrant trajectory. D) Histogram of activation delay.

Supplemental Figure S31

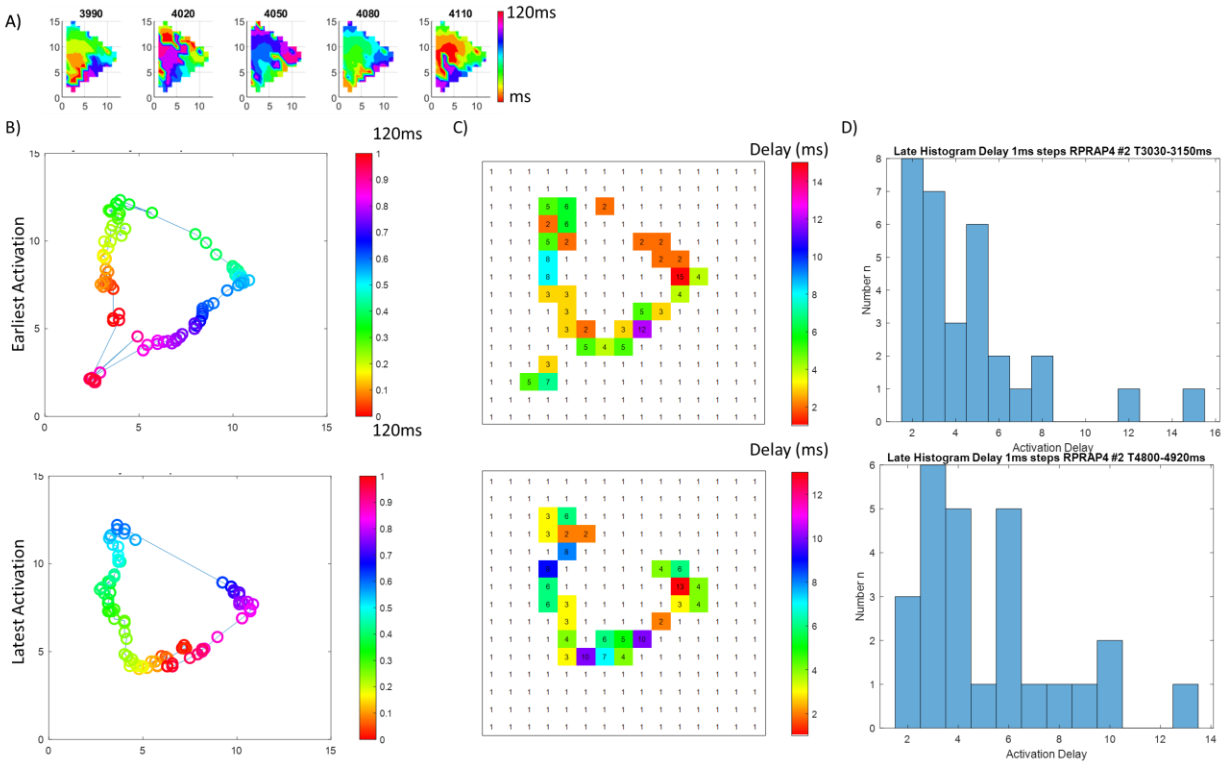


Heart #7, Region RAA: A) Stable Rotational Activity in LAT map. B) Trajectory of loop based on earliest activation (upper figure panel), latest activation (lower figure panel) within series of activation time maps in 1 msec steps. C) Activation delay along reentrant trajectory. D) Histogram of activation delay.

Supplemental Figure S32

Supplemental Figure S33

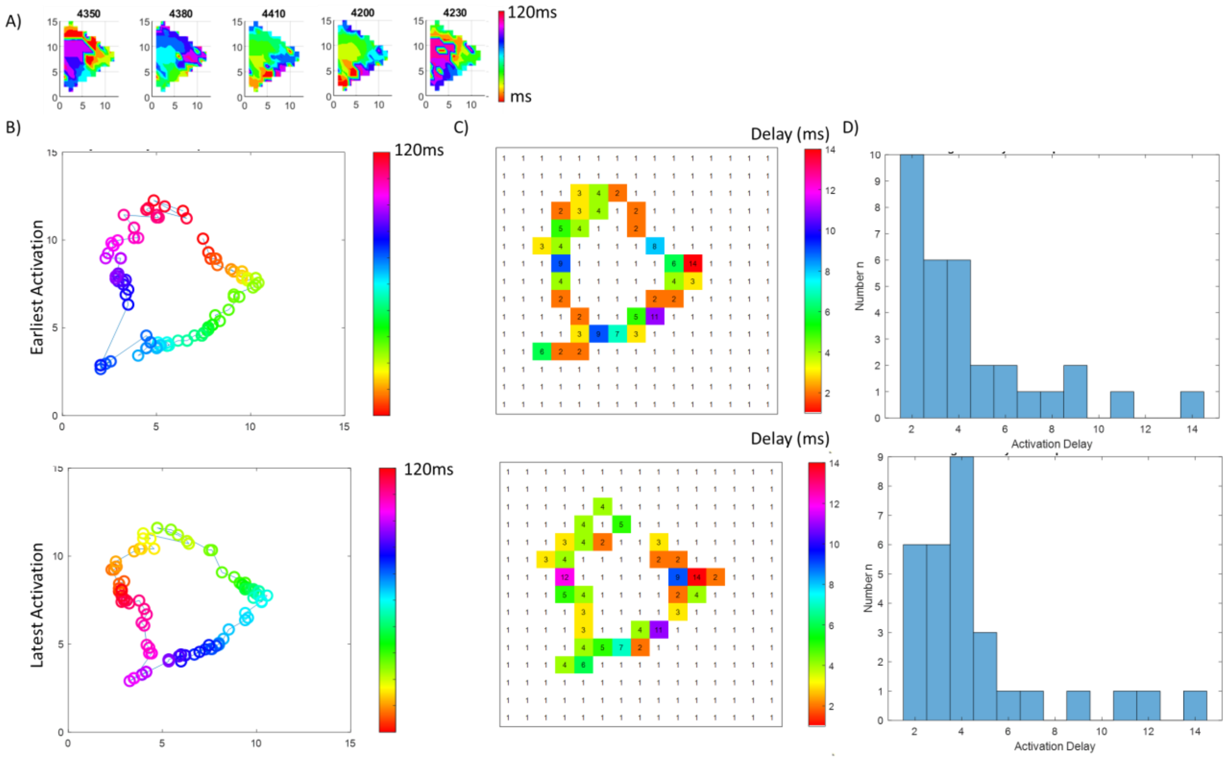


Heart #7, Region RAA: A) Stable Rotational Activity in LAT map. B) Trajectory of loop based on earliest activation (upper figure panel), latest activation (lower figure panel) within series of activation time maps in 1 msec steps. C) Activation delay along reentrant trajectory. D) Histogram of activation delay.

Supplemental Figure S34

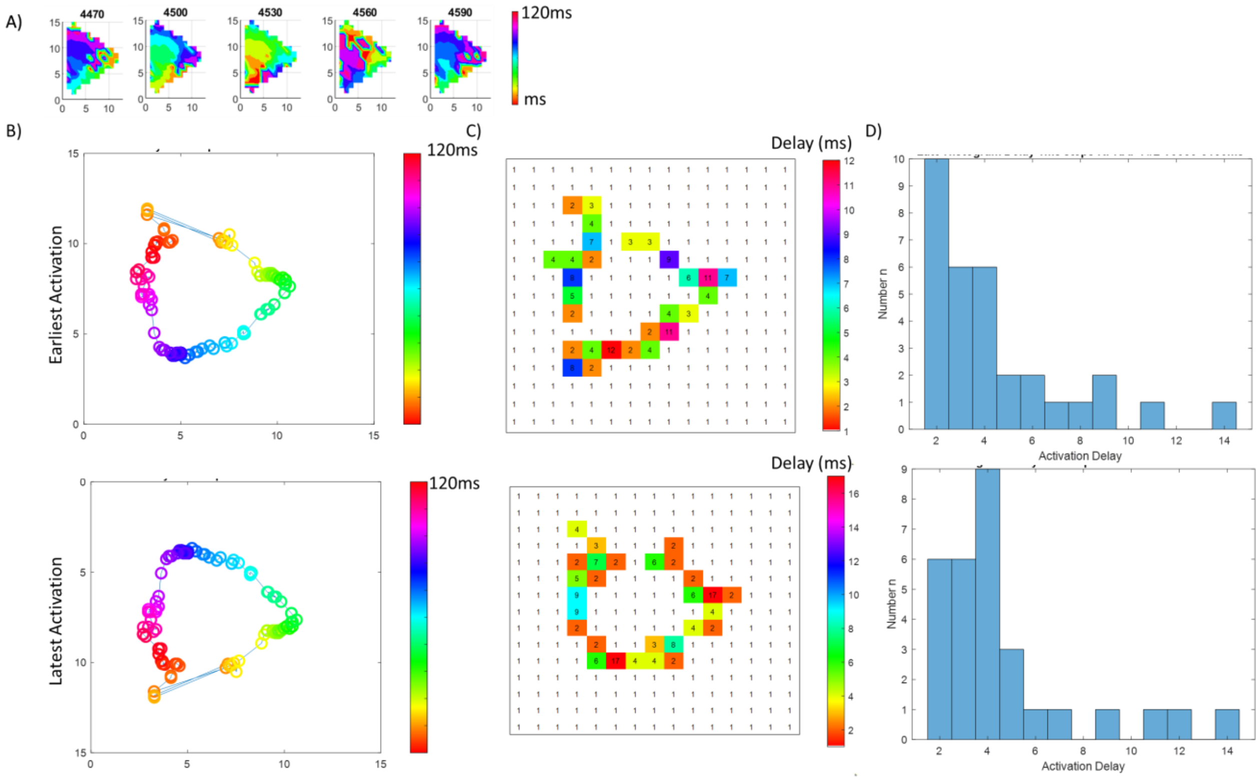


Heart #7, Region RAA: A) Stable Rotational Activity in LAT map. B) Trajectory of loop based on earliest activation (upper figure panel), latest activation (lower figure panel) within series of activation time maps in 1 msec steps. C) Activation delay along reentrant trajectory. D) Histogram of activation delay.

Supplemental Figure S35

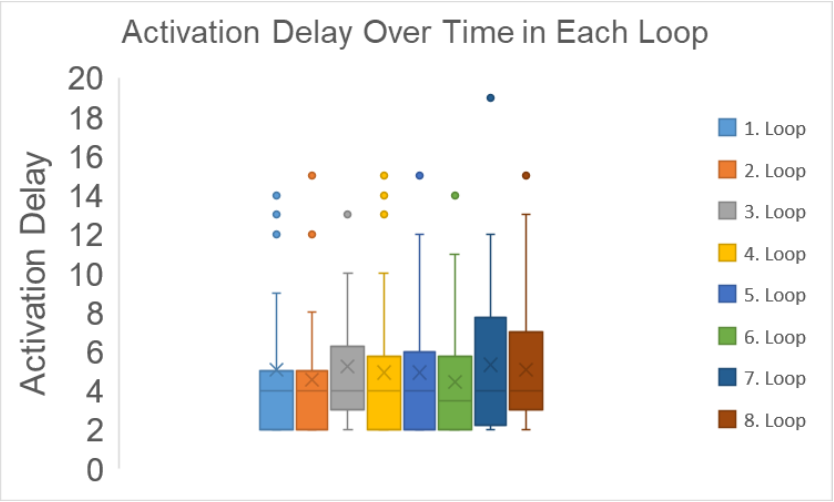


Heart #7, Activation delay along reentrant trajectory in 8 following reentrant loops.

Supplemental Figure S36

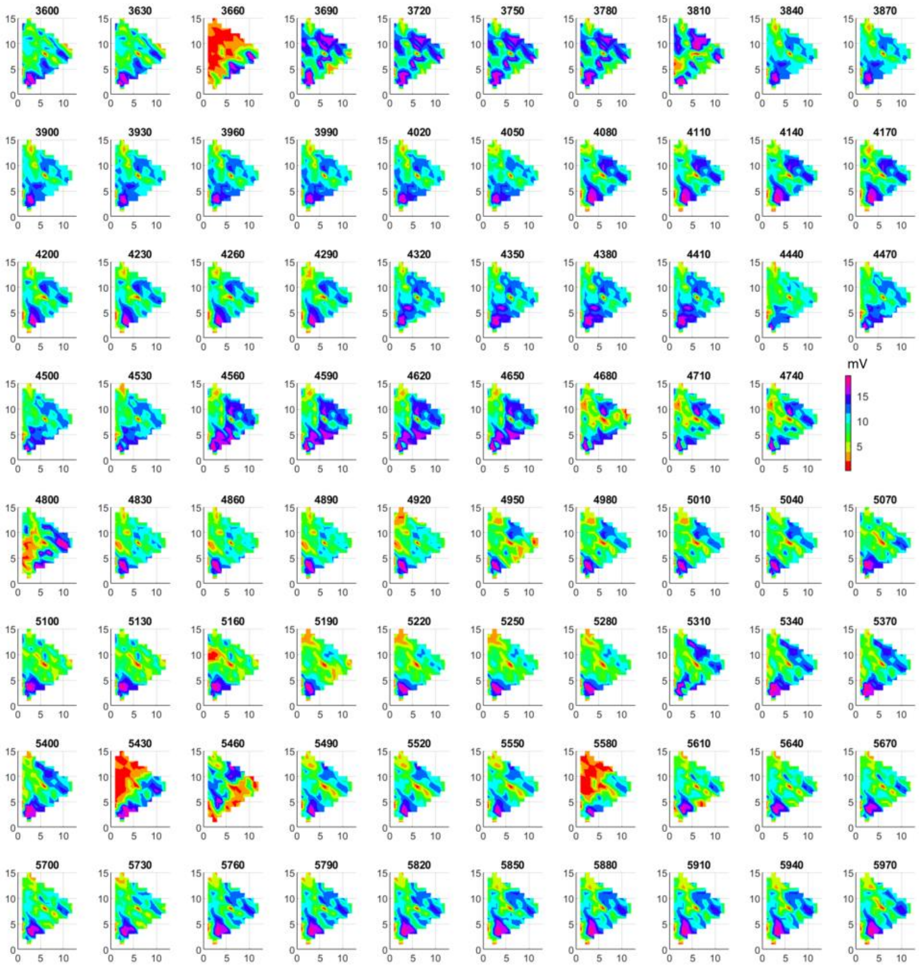

Heart #7, Region RAA: Bipolar voltage maps during rotational activity over time.

Supplemental Figure S37

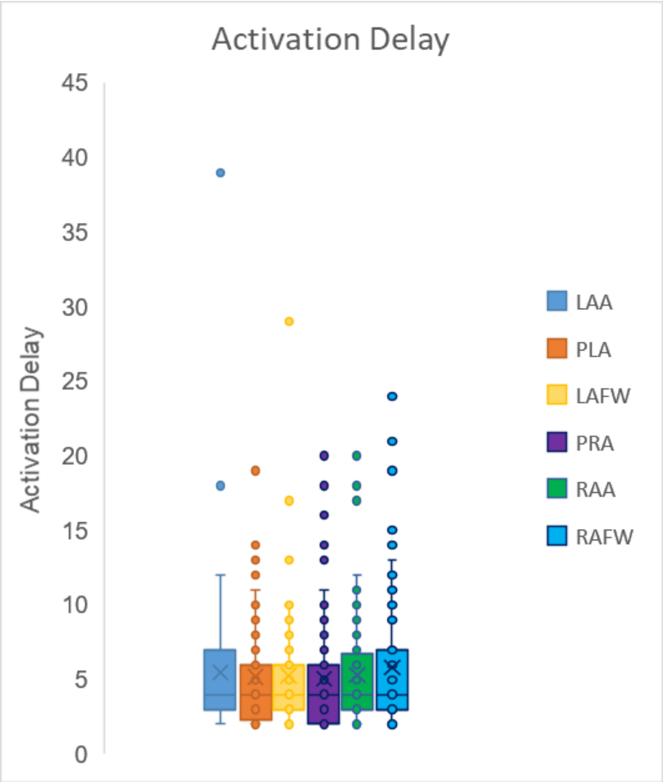


Heart #7, Activation delay along reentrant trajectory in all regions

Supplemental Figure S38

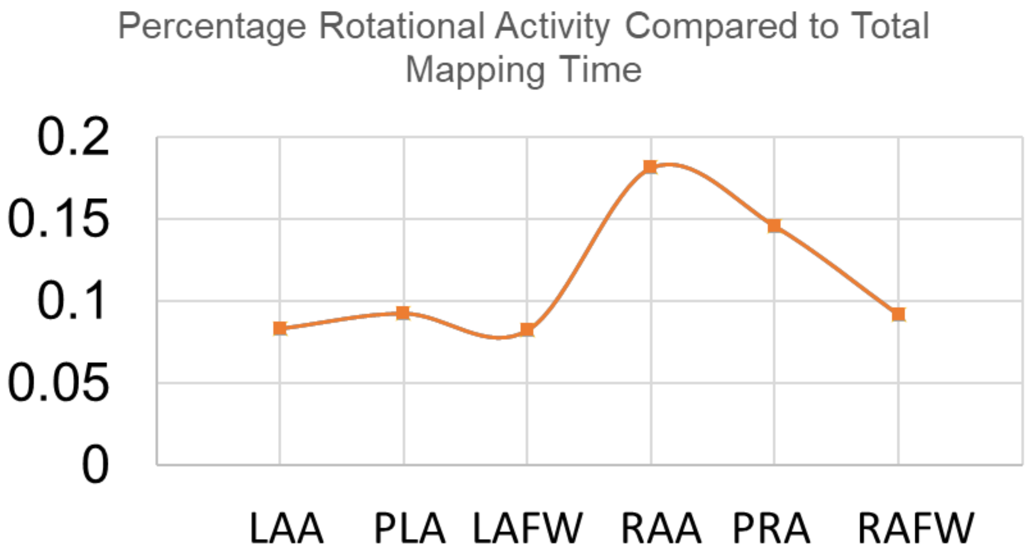


Percentage of rotational activity compared to total mapping time in all regions.

Supplemental Figure S39

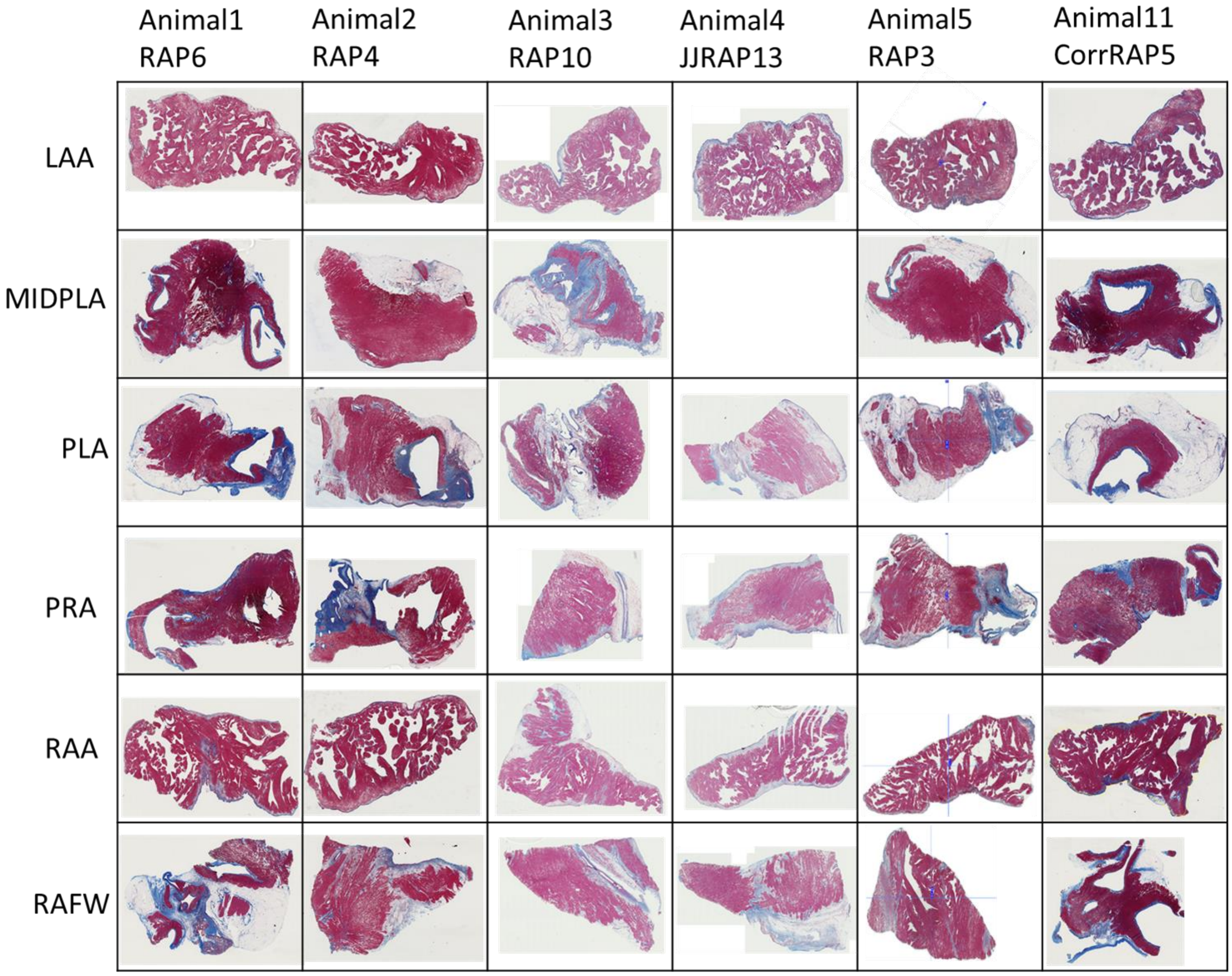

Supplemental Figure S40

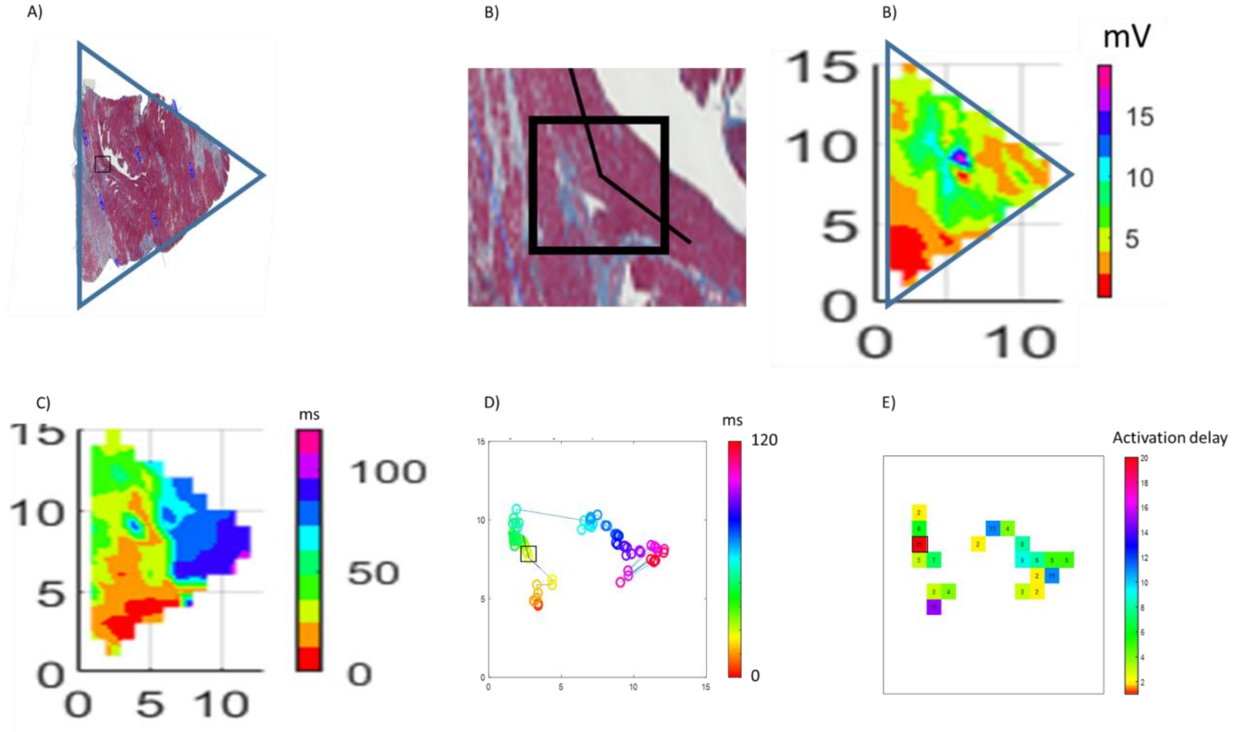


A) Masson's trichrome stained tissue section of heart 7 in the RAFW. Blue indicates fibrosis, red normal tissue. B) Zoomed tissue section of highest activation delay (black rectangle compare panel A, D, E) at overlapping fiber crossing (138 degree between fiber directions) and dense fibrosis. C) Corresponding bipolar voltage map. Low voltage (red) overlaps with larger fibrosis regions. D) Activation time map. E) Trajectory of rotational activity. F) Activation delay at dense fibrosis region. Highest activation delay at overlapping fiber crossing and dense fibrosis.

Supplemental Figure S41

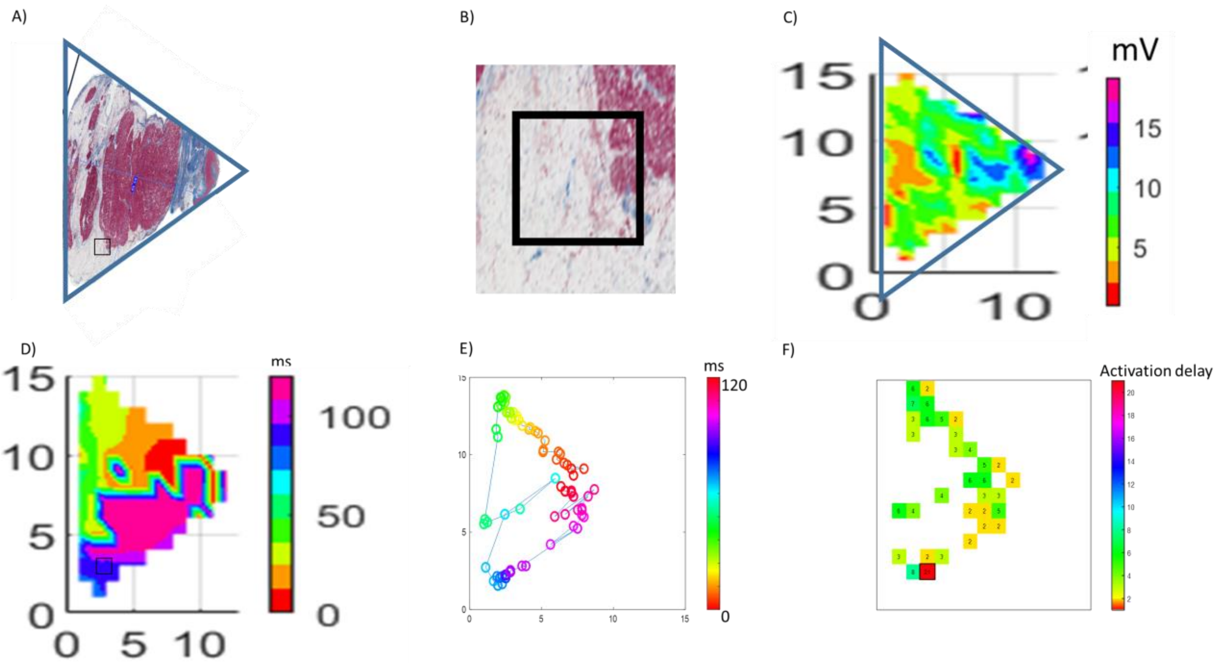


A) Masson's trichrome stained tissue section of heart 7 in the LAFW. Blue indicates fibrosis, red normal tissue. B) Zoomed tissue section of highest activation delay (black rectangle compare panel A, D, E) at dense fat tissue. C) Corresponding bipolar voltage map. Low voltage (red) overlaps partially with fat and fibrosis regions. D) Activation time map of rotational activity. E) Trajectory of rotational activity. E) Activation delay at trajectory of rotational activity. F) Highest activation delay at most dense fat tissue.

Supplemental Figure S42

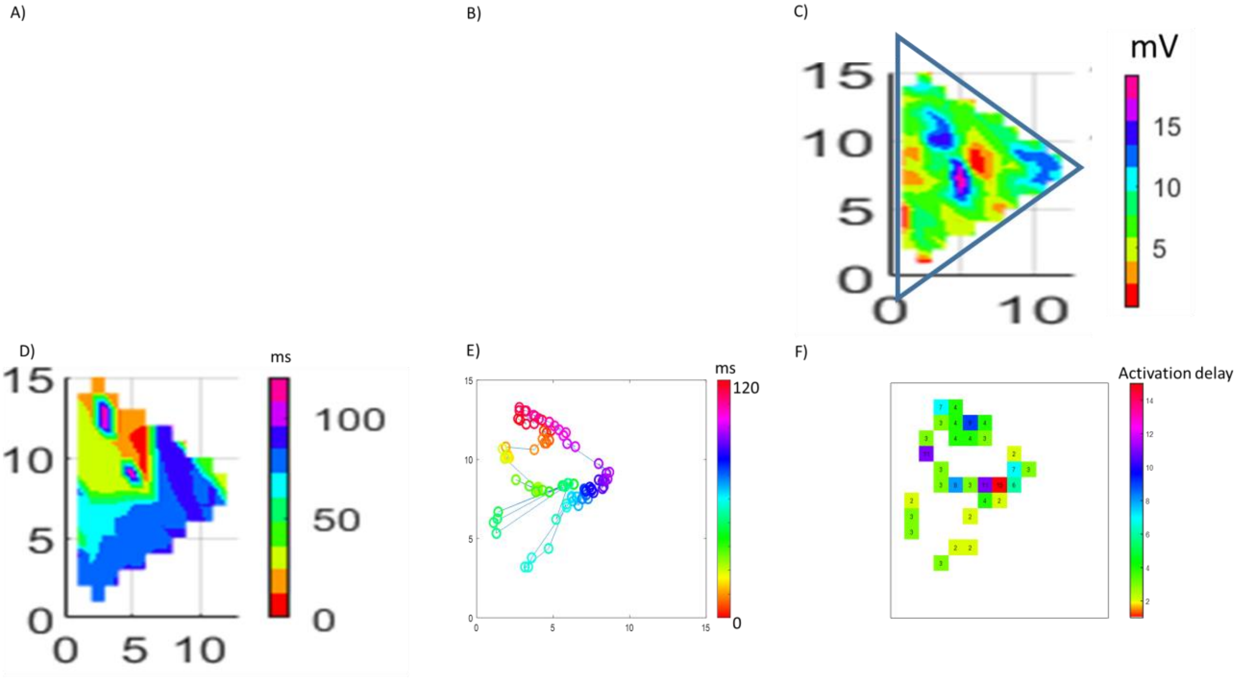


A) Masson's trichrome stained tissue section of heart 7 in the PLA. Blue indicates fibrosis, red normal tissue. B) Zoomed tissue section of highest activation delay (black rectangle compare panel A, D, E) at dense fat tissue. C) Corresponding bipolar voltage map. Low voltage (red) overlaps partially with fat and fibrosis regions. D) Activation time map of rotational activity. E) Trajectory of rotational activity. E) Activation delay at trajectory of rotational activity. F) Highest activation delay at most dense fat tissue.

Supplemental Figure S43

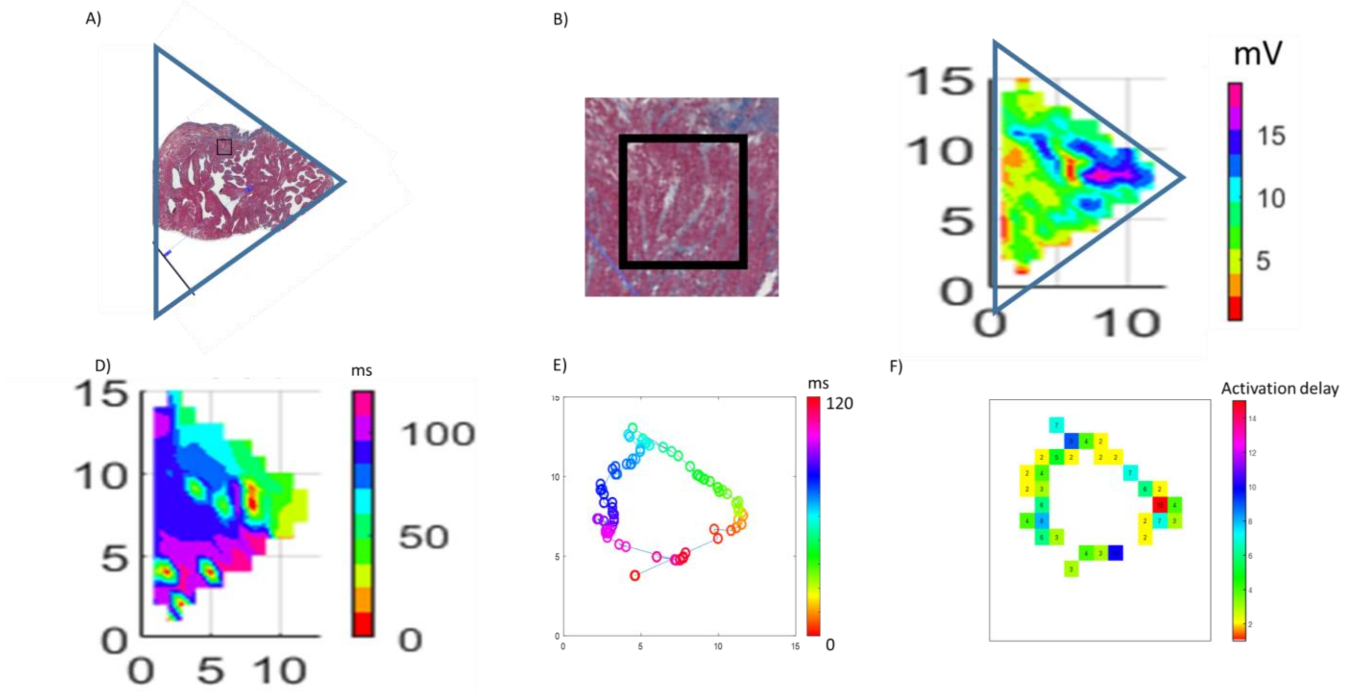


A) Masson's trichrome stained tissue section of heart 7 in the LAA. Blue indicates fibrosis, red normal tissue. B) Zoomed tissue section of highest activation delay (black rectangle compare panel A, D, E) at patchy fibrosis tissue. C) Corresponding bipolar voltage map. Low voltage (red) overlaps partially with fibrosis regions. D) Activation time map of rotational activity. E) Trajectory of rotational activity. E) Activation delay at trajectory of rotational activity. F) High activation delay at patchy fibrosis tissue.

Supplemental Figure S44

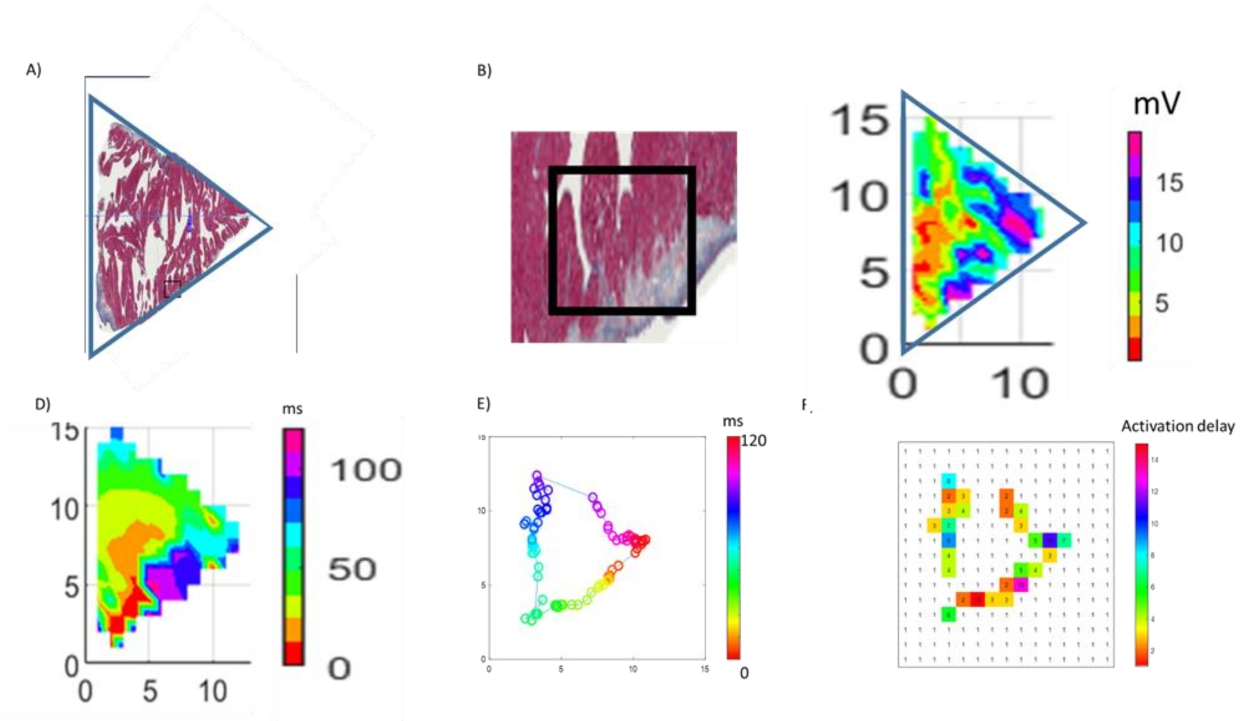


A) Masson’s trichrome stained tissue section of heart 7 in the RAA. Blue indicates fibrosis, red normal tissue. B) Zoomed tissue section of highest activation delay (black rectangle compare panel A, D, E) at dense fibrosis tissue. C) Corresponding bipolar voltage map. Low voltage (red) overlaps partially with fibrosis regions. D) Activation time map of rotational activity. E) Trajectory of rotational activity. E) Activation delay at trajectory of rotational activity. F) High activation delay at dense fibrosis tissue.

Supplemental Figure S45

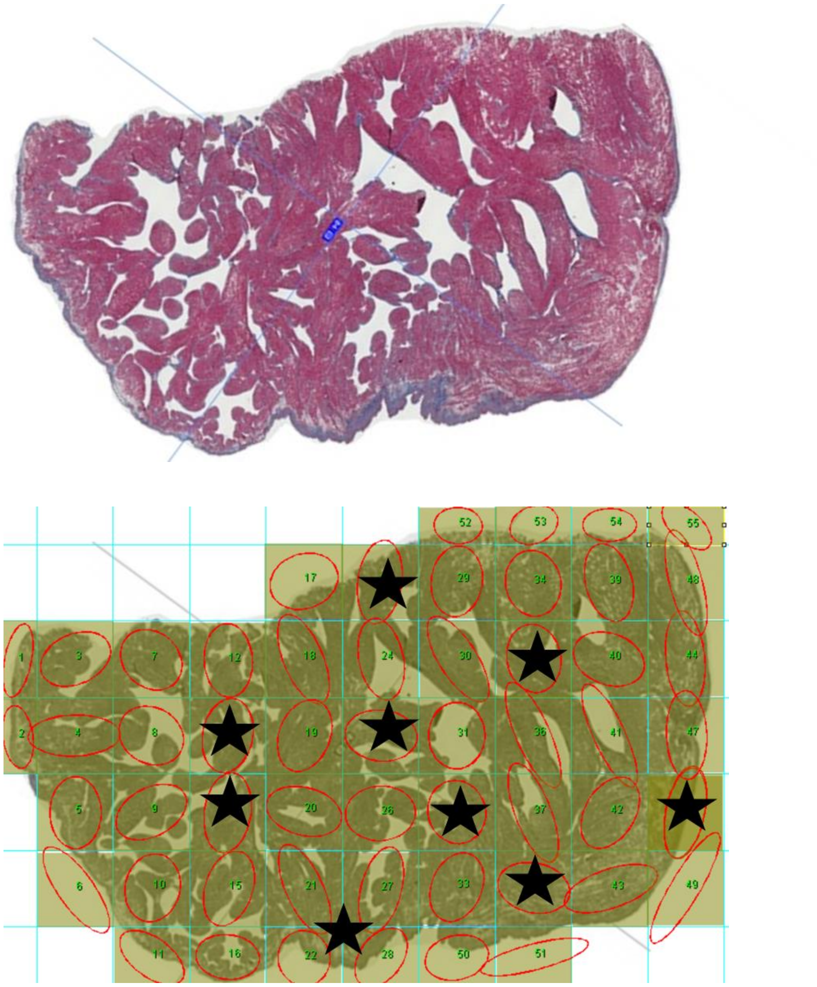

LAA, Upper panel:  Masson's trichrome stained tissue section; Lower panel: Red ovals indicate direction of fiber orientation in ROI's. Fiber crossings marked in black.

Supplemental Figure S46

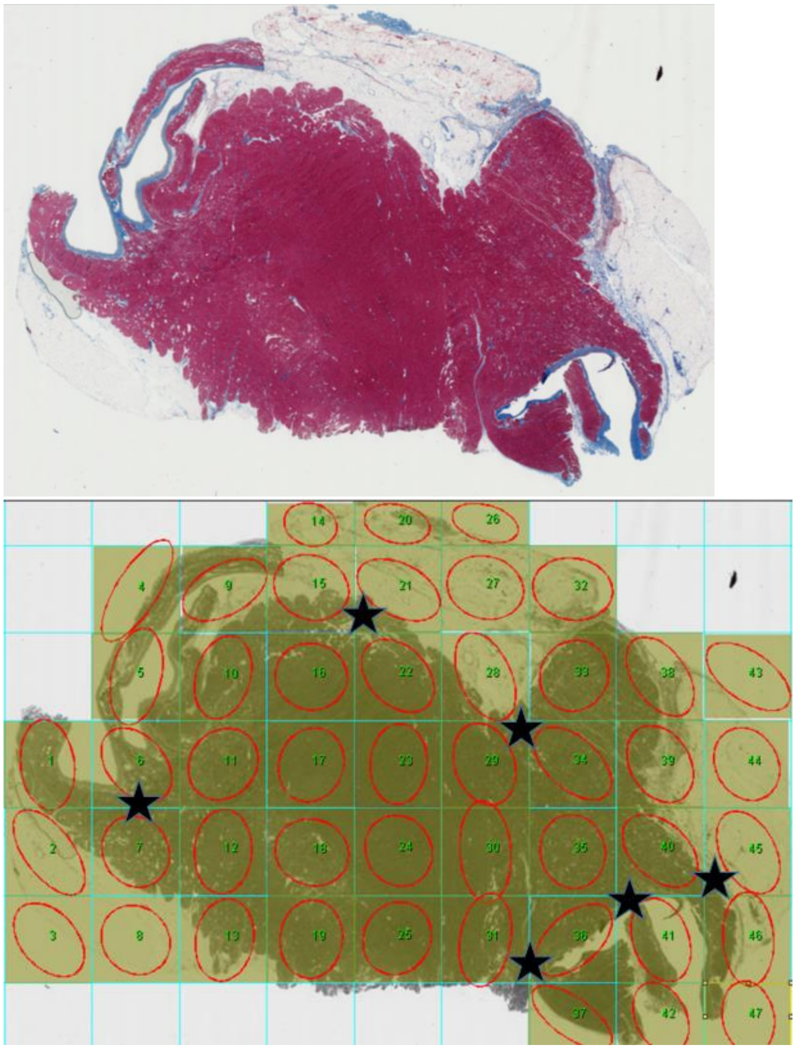

PLA, Upper panel: Masson’s trichrome stained tissue section; Lower panel: Red ovals indicate direction of fiber orientation in ROI’s. Fiber crossings marked in black.

Supplemental Figure S47

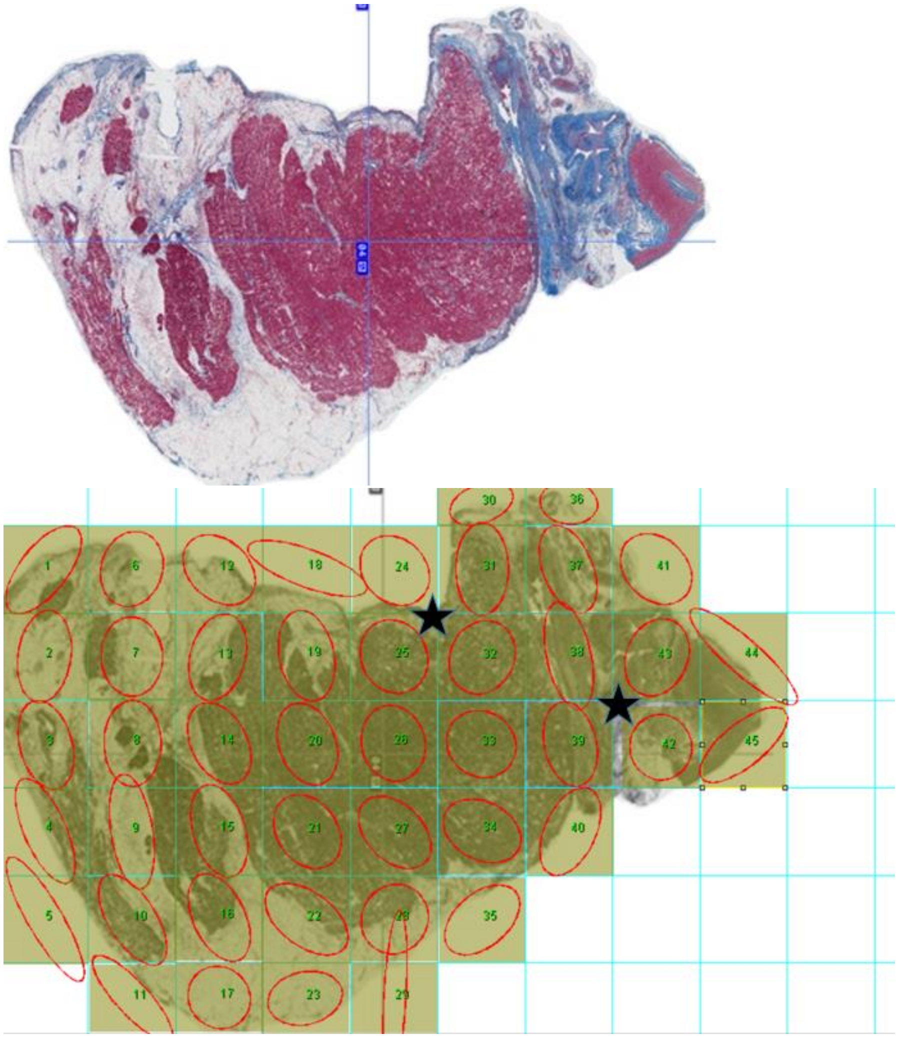

LAFW, Upper panel:  Masson's trichrome stained tissue section; Lower panel: Red ovals indicate direction of fiber orientation in ROI's. Fiber crossings marked in black.

Supplemental Figure S48

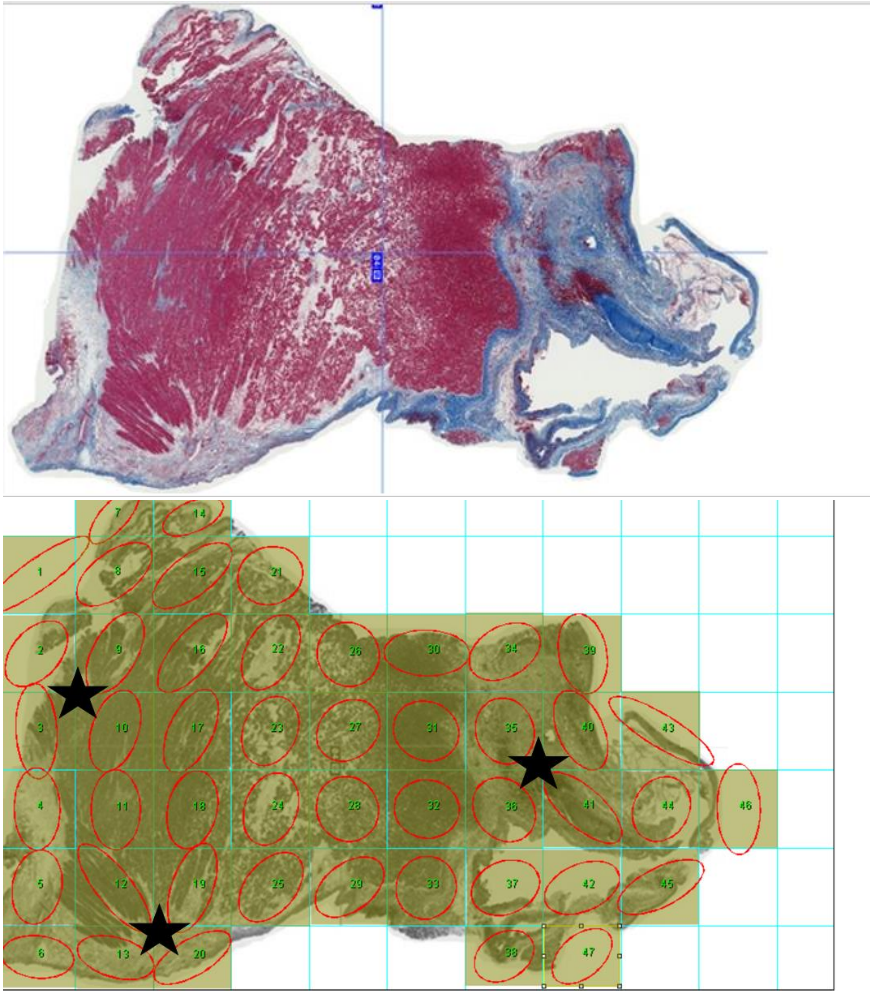

PRA, Upper panel:  Masson's trichrome stained tissue section; Lower panel: Red ovals indicate direction of fiber orientation in ROI's. Fiber crossings marked in black.

Supplemental Figure S49

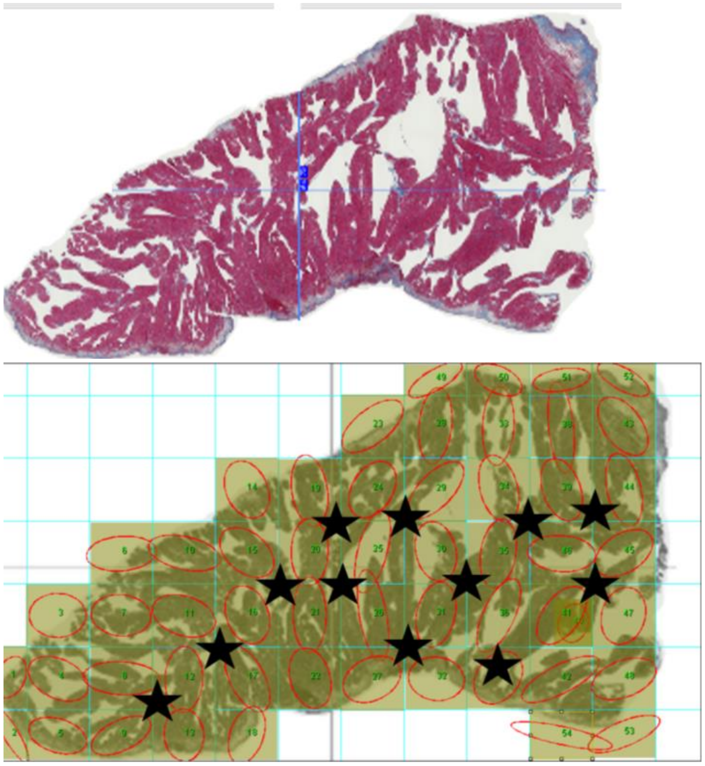

RAA, Upper panel:  Masson's trichrome stained tissue section; Lower panel: Red ovals indicate direction of fiber orientation in ROI's. Fiber crossings marked in black.

Supplemental Figure S50

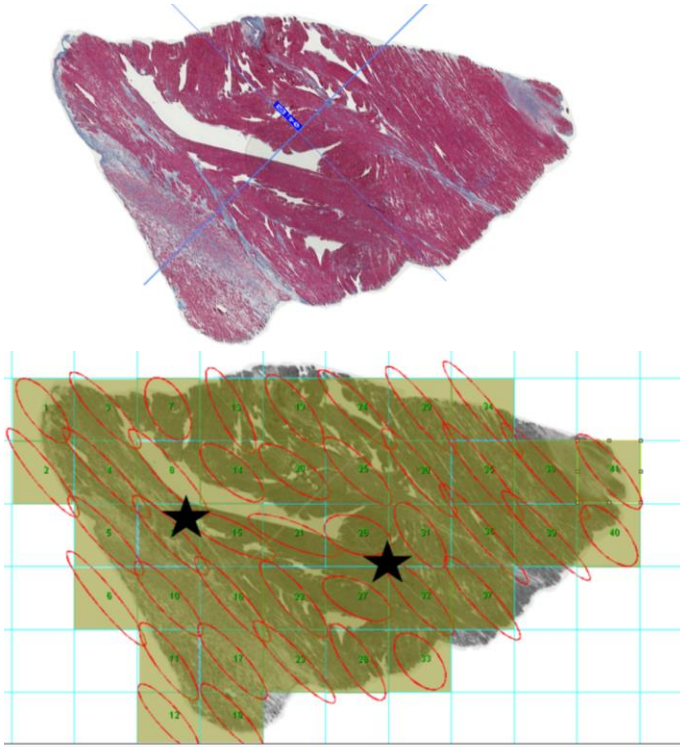

RAFW, Upper panel:  Masson's trichrome stained tissue section; Lower panel: Red ovals indicate direction of fiber orientation in ROI's. Fiber crossings marked in black.

Supplemental Figure S51

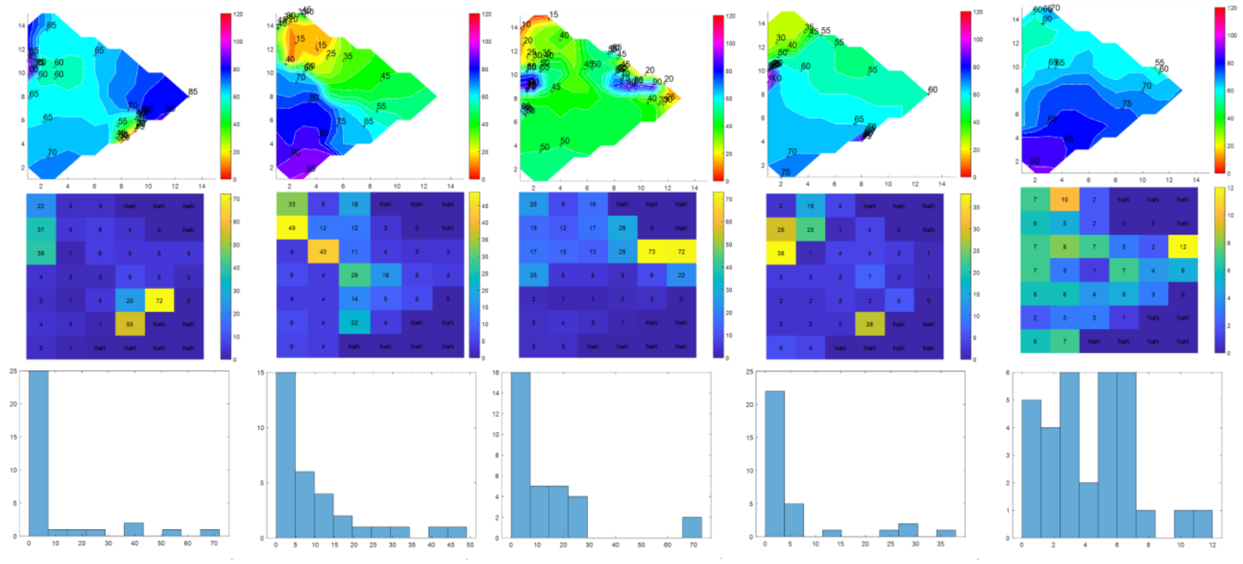

A) Activation delay in AF (left) and sinus rhythm/ paced rhythm showing similar located slow conduction zones.

Supplemental Figure S52

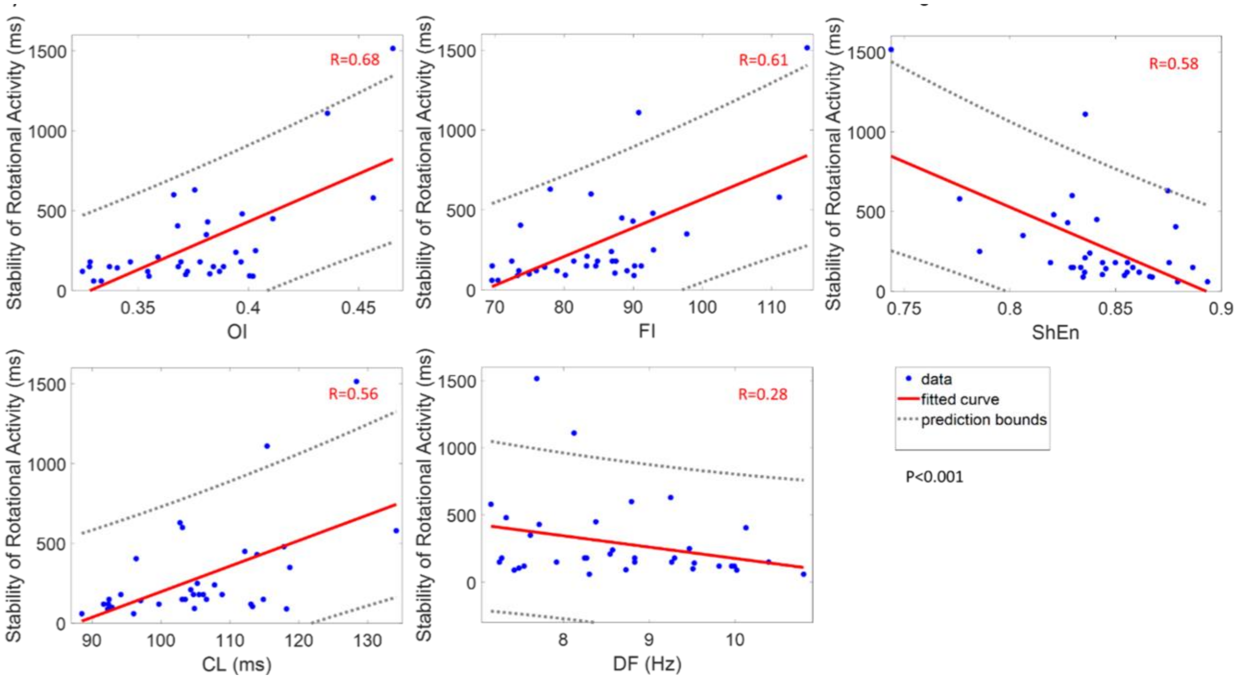


Correlation of electrogram measures with stability of rotational activity in the order of the highest correlation to less correlation. Moderately correlation of stability of rotational activity with electrogram measures OI (R=0.68), FI (R=0.61), ShEn (R=0.58) and CL (R=0.56), but not with DF (R=0.28). P < 0.001 for all comparisons.

Supplemental Figure S53

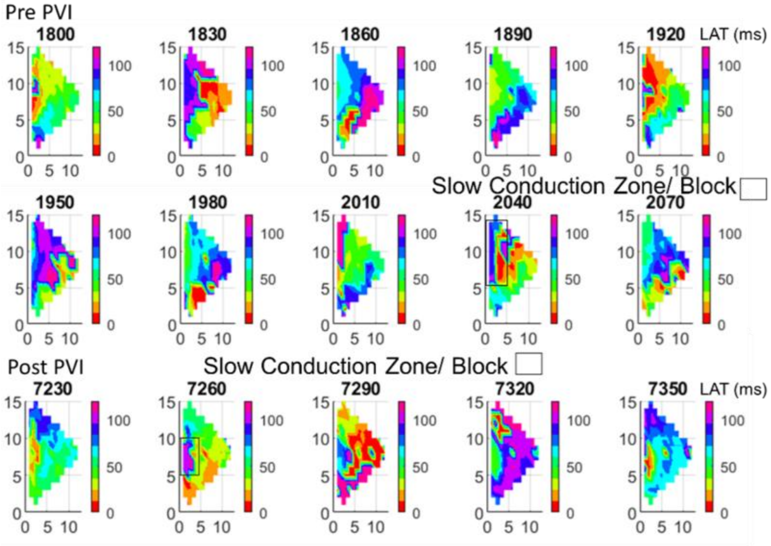


Slow conduction zones were similar located pre and post PVI. Reduced slow conduction zone/ block (marked in black) after PVI.
